# A Case for Automated Segmentation of MRI Data in Milder Neurodegenerative Diseases

**DOI:** 10.1101/2025.02.18.25322304

**Authors:** Connor J. Lewis, Jean M. Johnston, Precilla D’Souza, Josephine Kolstad, Christopher Zoppo, Zeynep Vardar, Anna Luisa Kühn, Ahmet Peker, Zubir S. Rentiya, William A. Gahl, Mohammed Salman Shazeeb, Cynthia J. Tifft, Maria T. Acosta

## Abstract

**Background:** Volumetric analysis and segmentation of magnetic resonance imaging (MRI) data is an important tool for evaluating neurological disease progression and neurodevelopment. Fully automated segmentation pipelines offer faster and more reproducible results. However, since these analysis pipelines were trained on or run based on atlases consisting of neurotypical controls, it is important to evaluate how accurate these methods are for neurodegenerative diseases. In this study, we compared 5 fully automated segmentation pipelines including FSL, Freesurfer, volBrain, SPM12, and SimNIBS with a manual segmentation process in GM1 gangliosidosis patients and neurotypical controls.

**Methods:** We analyzed 45 MRI scans from 16 juvenile GM1 gangliosidosis patients, 11 MRI scans from 8 late-infantile GM1 gangliosidosis patients, and 19 MRI scans from 11 neurotypical controls. We compared results for 7 brain structures including volumes of the total brain, bilateral thalamus, ventricles, bilateral caudate nucleus, bilateral lentiform nucleus, corpus callosum, and cerebellum.

**Results:** We found volBrain’s *vol2Brain* pipeline to have the strongest correlations with the manual segmentation process for the whole brain, ventricles, and thalamus. We also found Freesurfer’s *recon-all* pipeline to have the strongest correlations with the manual segmentation process for the caudate nucleus. For the cerebellum, we found a combination of volBrain’s *vol2Brain* and SimNIBS’ *headreco* to have the strongest correlations depending on the cohort. For the lentiform nucleus, we found a combination of *recon-all* and FSL’s *FIRST* to give the strongest correlations depending on the cohort. Lastly, we found segmentation of the corpus callosum to be highly variable.

**Conclusion:** Previous studies have considered automated segmentation techniques to be unreliable, particularly in neurodegenerative diseases. However, in our study we produced results comparable to those obtained with a manual segmentation process. While manual segmentation processes conducted by neuroradiologists remain the gold standard, we present evidence to the capabilities and advantages of using an automated process including the ability to segment white matter throughout the brain or analyze large datasets, which pose feasibility issues to fully manual processes. Future investigations should consider the use of artificial intelligence-based segmentation pipelines to determine their accuracy in GM1 gangliosidosis, lysosomal storage disorders, and other neurodegenerative diseases.

## Introduction

Volumetric analysis of magnetic resonance imaging (MRI) has proven useful in diagnosing and monitoring the progression of various neurological disorders.^1,2^ The capability to evaluate brain growth and development has provided important insights in brain development and pathogenesis.^3^ Numerous techniques for performing these volumetric calculations exist, including both manual and automated methods.^4,5^

Manual segmentation processing of MRI data involves a clinician manually tracing the structure of interest slice by slice. While these approaches are accurate, they are also time consuming and unrealistic for analyzing larger datasets.^6^ Inter-rater reliability issues are also noteworthy for manual approaches when results are compared between clinicians.^7,8^ Automated segmentation processes have received increased attention with the capability of analyzing large datasets systematically without fatigue.^9^ Automated segmentation of MRI data typically falls into one of two categories: either atlas-based or artificial intelligence (AI)-based methods.^10^ Atlas-based techniques utilize either a hand-labeled or statistical atlas, which is registered to the scan being analyzed to identify neuroanatomical structures.^11^ AI-based segmentation processes are trained to analyze neuroanatomical structures based on labeled training data and are then deployed to analyze MRI scans.^12^ AI techniques are more flexible based on their training data and once trained may reduce the computational cost.^13^ However, AI approaches have not had widespread adoption at the time of this study, and currently approaches are more specialized.^14–18^

Similarly, atlas-based segmentation processes have their own challenges. Firstly, the atlases utilized in the most common MRI segmentation pipelines may not be representative of the population being analyzed. For instance, Freesurfer utilizes the Desikan-Killiany dataset with 40 neurotypical control participants between the ages of 19 and 86 years, which may limit interpretation of subjects who are outside of this age range or who might be impaired.^19^ Similarly, with the pediatric brain specifically, challenges arise including increased noise, reduced contrast between tissues, and ongoing myelination.^20^ Furthermore, some of the most utilized automated segmentation methods ignore smaller substructures like the hypothalamus,^21^ pons,^22^ pituitary gland,^23^ and optic nerve.^24^ Ultimately, it is important to investigate which atlas-based technique is suitable for each analysis. In this study, we aim to investigate some of the most frequently utilized atlas-based techniques including Freesurfer, FSL, volBrain, and SPM in GM1 gangliosidosis brains, to understand which pipeline provides the highest accuracy of volumetric measurements compared to a manual approach.

GM1 gangliosidosis is an inherited ultra-rare neurodegenerative lysosomal storage disorder caused by variants in the *GLB1* gene encoding β-galactosidase.^25^ Three clinical subtypes for GM1 exist based on the age of symptom onset and residual enzyme activity.^26^ Type I (infantile) GM1 is the most severe form of the disease with symptoms beginning in the first year of life and death often before the age of 3.^27^ Type II GM1 gangliosidosis can be further classified into two forms (late-infantile and juvenile) with the late-infantile form leading to symptom onset between age 1 and 2 and survival into the second decade. Juvenile GM1 patients have symptom onset between 3 and 5 years old with survival into the fourth decade.^28^ Type III (adult) GM1 gangliosidosis patients typically have manifesting symptoms in the second decade, slower disease progression, and the longest survival. Currently, there are no approved therapies for GM1, and the disease is uniformly fatal. However, investigations into adeno-associated virus (AAV) mediated gene therapy have shown promise in animal models and one clinical trial in Type I and II GM1 gangliosidosis patients is currently underway (ClinicalTrials.gov Identifier NCT03952637).^29,30^

Previous investigations utilizing brain MRI in Type II GM1 gangliosidosis patients have demonstrated numerous findings.^31,32^ Atrophy of the cerebral cortex, corpus callosum, caudate nucleus, cerebellum, cerebellar white matter, basal ganglia, and associated ventricular enlargement have all been described.^31,32^ In this study, we first investigated the performance of 5 different fully automated MRI segmentation pipelines compared to a manual process in identifying MRI pathogenesis in GM1 gangliosidosis patients (Figure 1). Second, we evaluated which automated pipeline gave the most accurate results in terms of volumetric analysis of the different brain structures.

**Figure 1.**
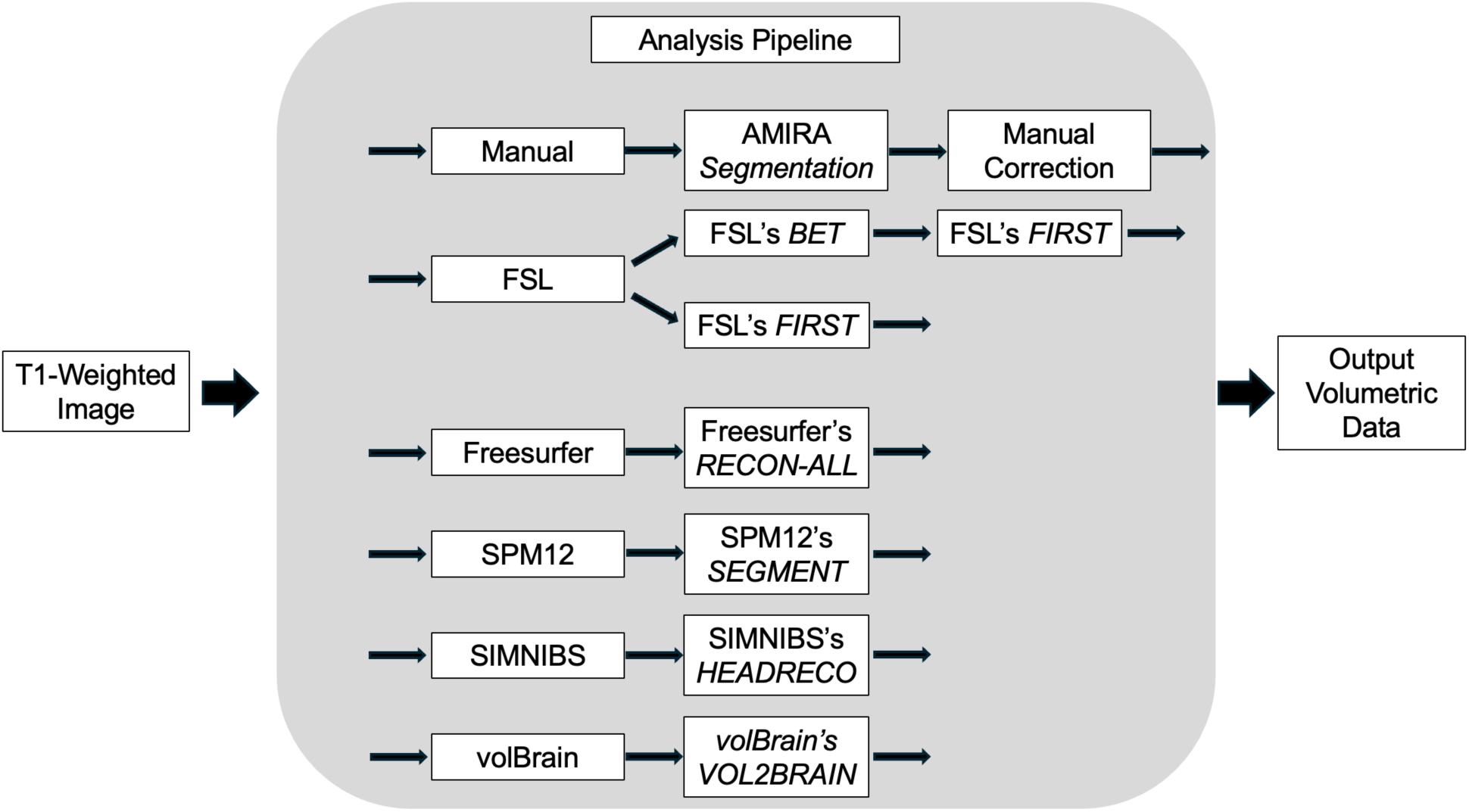
Segmentation Analysis Pipeline Overview. T1-Weighted MRI scans were processed through 5 fully automated segmentation pipelines in addition to the manual segmentation process. FSL required two different sub-pipelines. FSL’s *FIRST* was used to analyze bilateral volumes of the thalamus, caudate nucleus, and lentiform nucleus and FSL’s *FAST* was used to analyze whole brain volumes.

## Methods

### Type 2 GM1 Gangliosidosis Participants

Twenty-four GM1 patients from the “Natural History of Glycosphingolipid Storage Disorders and Glycoprotein Disorders” (ClinicalTrials.gov ID: NCT00029965) were included in this study.^33^ As described in D’Souza et al, a GM1 diagnosis was made through β-galactosidase enzyme deficiency or by biallelic variants in *GLB1*.^25^ Forty-five MRI scans from 16 juvenile (baseline age: 11.8 ± 4.9 years) patients were included in this study alongside 11 MRI scans from 8 late-infantile patients (Figure 2, baseline age: 5.5 ± 1.8 years).

**Figure 2.**
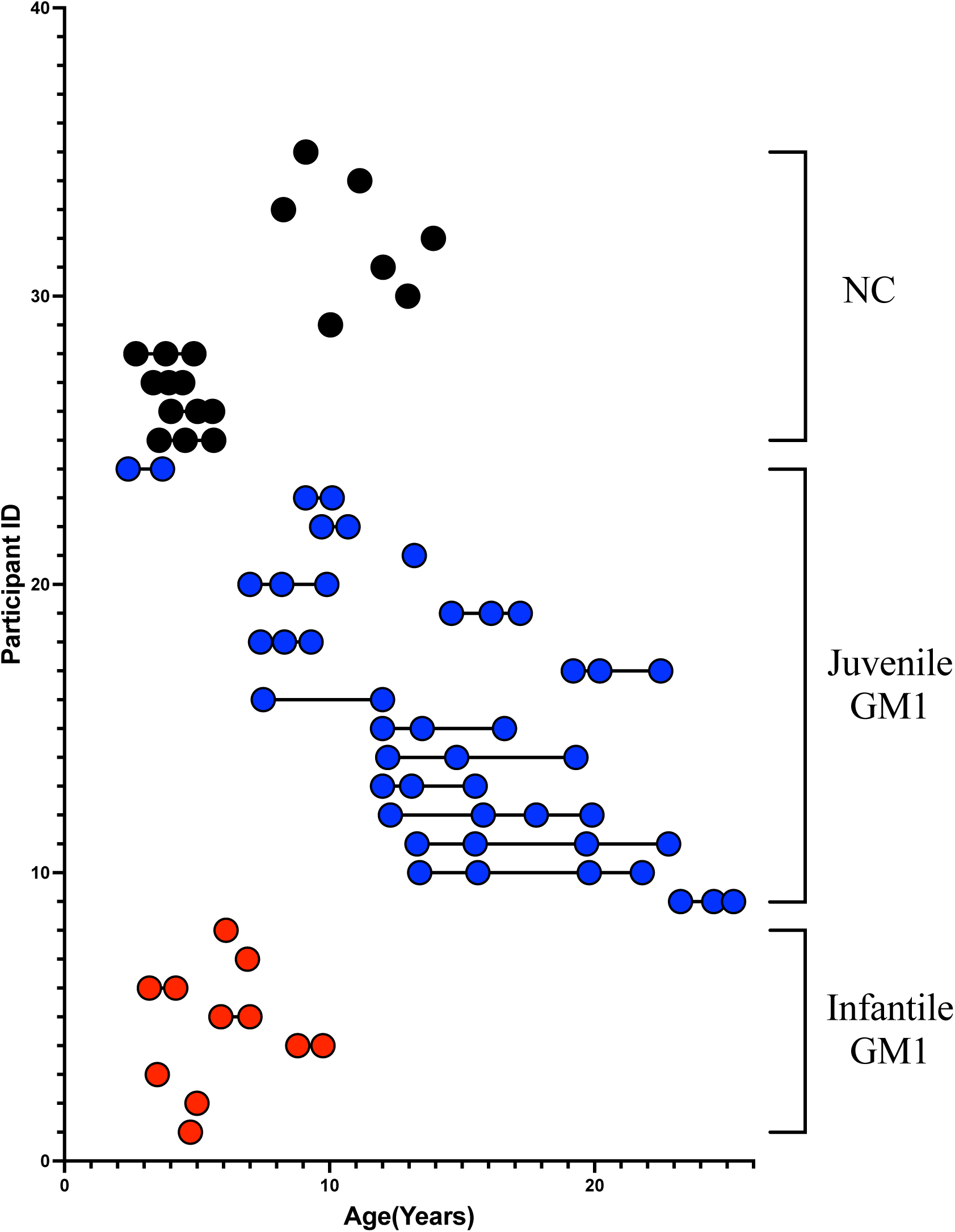
Age stratification of each participant at T1-weighted MRI scan. Infantile GM1 patients are shown in Red. Juvenile GM1 patients are shown in blue. Neurotypical controls are shown in black. Connecting lines indicate participant had repeated MRI scans.

### Neurotypical Controls

Neurotypical early childhood control MRI scans were gathered from Open Science Framework and included participants from the “Calgary Preschool magnetic resonance imaging (MRI) dataset” and consisted of participants between 2 and 8 years of age from the University of Calgary.^34^ Late childhood and adolescent neurotypical control MRI were gathered from Figshare and included participants from the “Detailing neuroanatomical development in late childhood and early adolescence using NODDI” study and consisted of participants between 8 and 13 years of age also from the University of Calgary.^35^ In total 19 MRI scans were obtained, from 11 neurotypical controls (Figure 2, baseline age: 8.3 ± 4.2 years).

### T1-Weighted MRI Acquisition

MRI scans were performed on a Philips 3T system (Achieva, Philips Healthcare, Best, The Netherlands) for all GM1 patients under sedation at all time-points with an 8-channel SENSE head coil. Images were acquired using a 3D T1-weighted protocol with slice thickness of 1 mm, repetition time (TR) of 11 ms, and an echo time (TE) of 7 ms. Unprocessed digital imaging and communications in medicine (DICOM) images were converted to NIfTI using *dcm2niix*.^36^ Calgary preschool and adolescent neurotypical controls were scanned without sedation on a General Electric 3T system (MR750w, GE Healthcare, Chicago, IL, USA) using a 32-channel head coil.^34,35^ T1-weighted imaging from preschool neurotypical controls was acquired with TR/TE = 8.23/3.76 ms and resolution of 0.9 mm × 0.9 mm × 0.9 mm (resampled to 0.45 mm × 0.45 mm × 0.9 mm).^34^ T1-weighted imaging from adolescent Calgary neurotypical controls was acquired with TR/TE = 8.21/3.16 ms and 0.8 mm^3^ isotropic resolution.^35^

### Manual MRI Volumetric Segmentation

Volumetric segmentations were performed on the 3D T1-weighted images, which provided sufficient tissue contrast to visualize the structures of interest. A semi-automated approach was used with manual corrections for the larger structures and a manual process was implemented for the smaller structures as previously described Zoppo et al.^8^ The following structures were segmented on all the scans: whole brain (without ventricles), cerebellum, ventricles, corpus callosum, caudate, lentiform nucleus, and thalamus. In brief, a team of researchers and trained neuroradiologists used the AMIRA analysis software (Amira, Thermo Fischer Scientific, Waltham, MA) in the native image space to define the regions of interest (ROIs) around the boundaries of the structures using a combination of signal thresholding and/or manual demarcation on a slice-by-slice basis. The method for determining the segmentation boundaries of each structure was carried out as described in prior work.^8^ The ROIs from all slices were rendered into a 3D volume for each structure to estimate the structure volume.

### Freesurfer Volumetric Segmentation

Automated segmentation of MRI images was first performed using Freesurfer’s (v7.4.1) *recon-all* reconstruction pipeline to calculate volumes for the structures of interest.^37–45^ Volumes of the whole brain (sum of gray matter and white matter), ventricles, cerebellum, caudate, thalamus, lentiform nucleus (sum of the globus pallidus and putamen), and corpus callosum were calculated, corresponding to the manual segmentation process. Automated segmentation was performed post-hoc of the manual segmentation process.

### FSL Volumetric Segmentation

MRI data was also segmented using tools from the FMRIB Software Library (FSL).^46–48^ T1-weighted imaging was first sent through FSL’s *bet* for brain extraction. Brain extracted images were then analyzed using FSL’s *fast* to create partial volume maps of the cerebrospinal fluid, gray matter, and white matter. T1-weighted imaging was also sent through FSL’s *run_first_all* pipeline to segment subcortical structures including the bilateral thalamus, bilateral putamen, bilateral globus pallidus, and bilateral caudate nucleus. FSL’s *fslstats* was then used to calculate volumes from the partial volume maps and segmented images.^46–48^

### volBrain Volumetric Segmentation

Automated segmentation of MRI images was also performed using volBrain’s updated segmentation algorithm, vol2Brain.^49^ vol2Brain uses the preprocessing algorithm from the original volBrain pipeline, followed by segmentation.^50^ T1-weighted images in this study were uploaded to the volBrain server, and volumetric results were obtained for the intracranial volume, bilateral gray matter, bilateral white matter, cerebellum, bilateral thalamus, bilateral globus pallidus, bilateral putamen, and bilateral caudate nucleus.

### SPM Volumetric Segmentation

Automated segmentation of T1-weighted images was also performed using Statistical Parametric Mapping (SPM) 12 housed within MATLAB R2023a (The MathWorks Inc., Natick, MA, USA).^51^ Tissue probability maps of the cerebrospinal fluid, gray matter, and white matter were created using the Segment tool. FSL’s *fslstats* was then used to calculate volumes from the tissue probability maps.

### Headreco Volumetric Segmentation

T1-weighted imaging in this study was also sent through SimNIBS’ (v3.2.6) *headreco* segmentation pipeline.^52^ *Headreco* utilizes SPM12 followed by the Computational Anatomy Toolbox (CAT12) to improve segmentation by SPM12.^51–54^ Gray matter, white matter, and cerebrospinal fluid volumes were calculated from the final mask generated by *headreco* using *fslstats*. Ventricle volume was also calculated using *fslstats* from the ventricle mask generated by *headreco.* Masks for the bilateral thalamus, corpus callosum, and cerebellum were generated from CAT12 after deformation-based morphometry (DBM), registration to the IXI-database^55^ of 555 healthy participants, and the Hammers atlas^56^ was used to select ROIs. Volumes of the bilateral thalamus, corpus callosum, and cerebellum were calculated from each respective mask using *fslstats*.

### Statistical Analysis

Cross-sectional data was analyzed in GraphPad Prism (v10.1.0, GraphPad Software, Boston, MA, USA) using Welch’s 2 sample t-test to demonstrate differences between neurotypical controls and both juvenile and late-infantile GM1 patients. Neurotypical controls were selected to age-match the late-infantile (Supplement B) and juvenile (Supplement C) GM1 patients separately since the juvenile patients were older than the late-infantile patients. Linear regression modeling was performed in GraphPad Prism to calculate correlation coefficients (R^2^) and F-statistics; *p*-values from the F-statistics were compared between each automated segmentation process and the manual segmentation process for the 7 structures of interest.

## Results

### Whole Brain Volume

Manual whole brain volume segmentation demonstrated higher whole brain volume in neurotypical controls when compared to both late-infantile (*t*(12) = 3.47, *p* = 0.0047, Figure 3) and juvenile (*t*(19) = 2.87, *p* = 0.0101, Figure 4) GM1 patients. volBrain, SPM, and Headreco also demonstrated higher whole brain volume in neurotypical controls compared to late-infantile patients; however, Freesurfer and FSL did not find a significant difference between late-infantile patients and neurotypical controls. Freesurfer, volBrain, and SPM demonstrated higher whole brain volume in neurotypical controls compared to juvenile patients; however, FSL and Headreco did not find a significant difference between the two cohorts (Figure 4).

**Figure 3.**
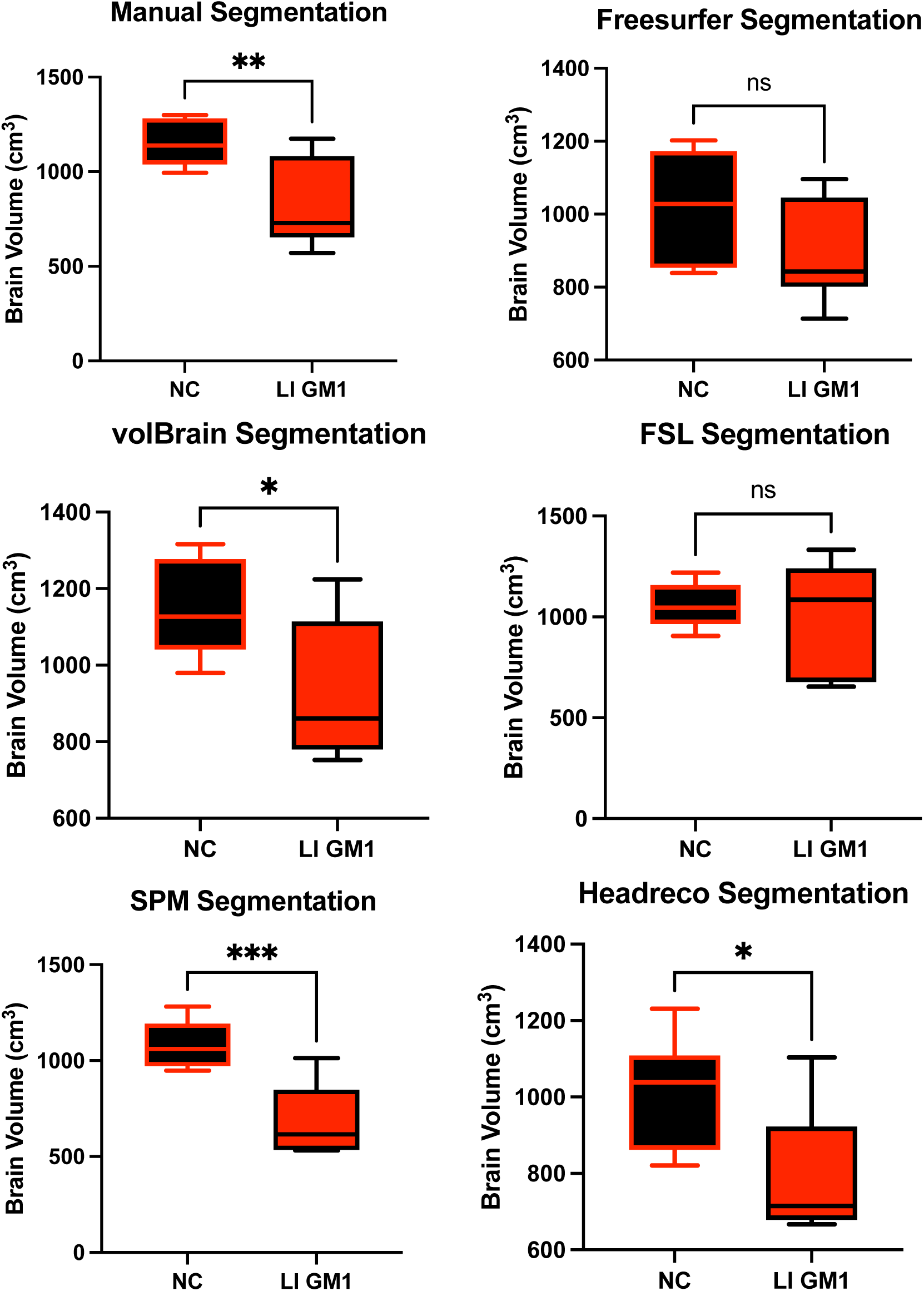
Cross-sectional evaluation of the 5 automated segmentation algorithms to demonstrate cohort differences in total brain volume. Late-infantile (LI) GM1 patients are shown in red. Neurotypical controls (NC) are shown in black. *P*-values were calculated from the *t*-statistic. * *P* < 0.05, ** *P* < 0.01, *** *P* < 0.001, **** *P* < 0.0001.

**Figure 4.**
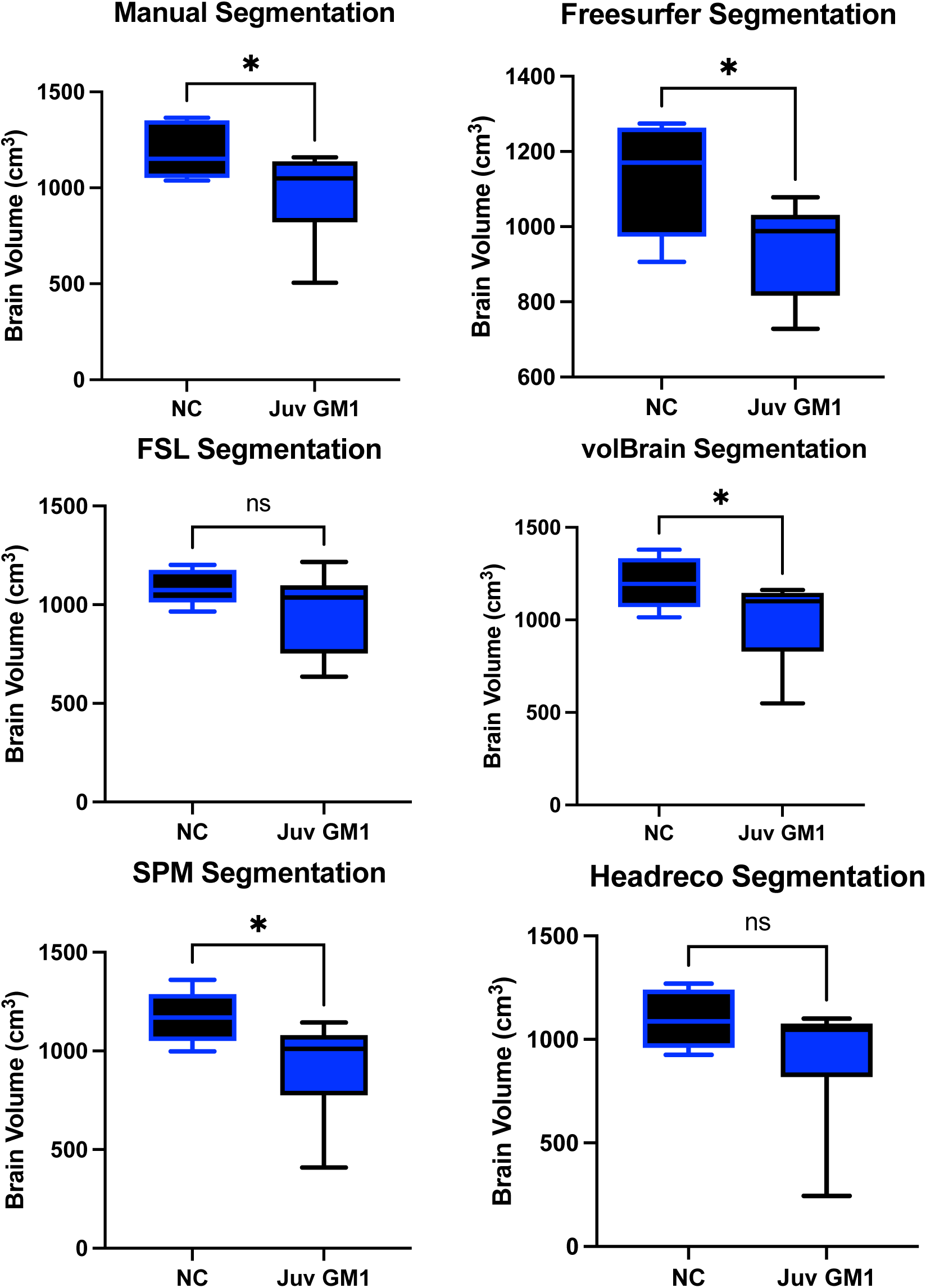
Cross-sectional evaluation of 5 automated segmentation algorithms ability to demonstrate cohort differences in total brain volume. Juvenile (Juv) GM1 patients are shown in blue. Neurotypical controls (NC) are shown in black. *P*-values were calculated from the *t*-statistic. * *P* < 0.05, ** *P* < 0.01, *** *P* < 0.001, **** *P* < 0.0001.

Correlations between the 5 fully automated segmentation processes and our manual process showed volBrain (R^2^ = 0.9852) and FSL (R^2^ = 0.9058) with R^2^ values above 0.90 in neurotypical controls (Figure 5). In juvenile GM1 patients, volBrain (R^2^ = 0.9583), Freesurfer (R^2^ = 0.9196), and SPM (R^2^ = 0.9642) all had R^2^ values above 0.90 (Figure 8). In late-infantile GM1 patients, volBrain (R^2^ = 0.9169) was the only fully automated segmentation process with an R^2^ above 0.90 when compared to the manual process (Figure 9).

**Figure 5.**
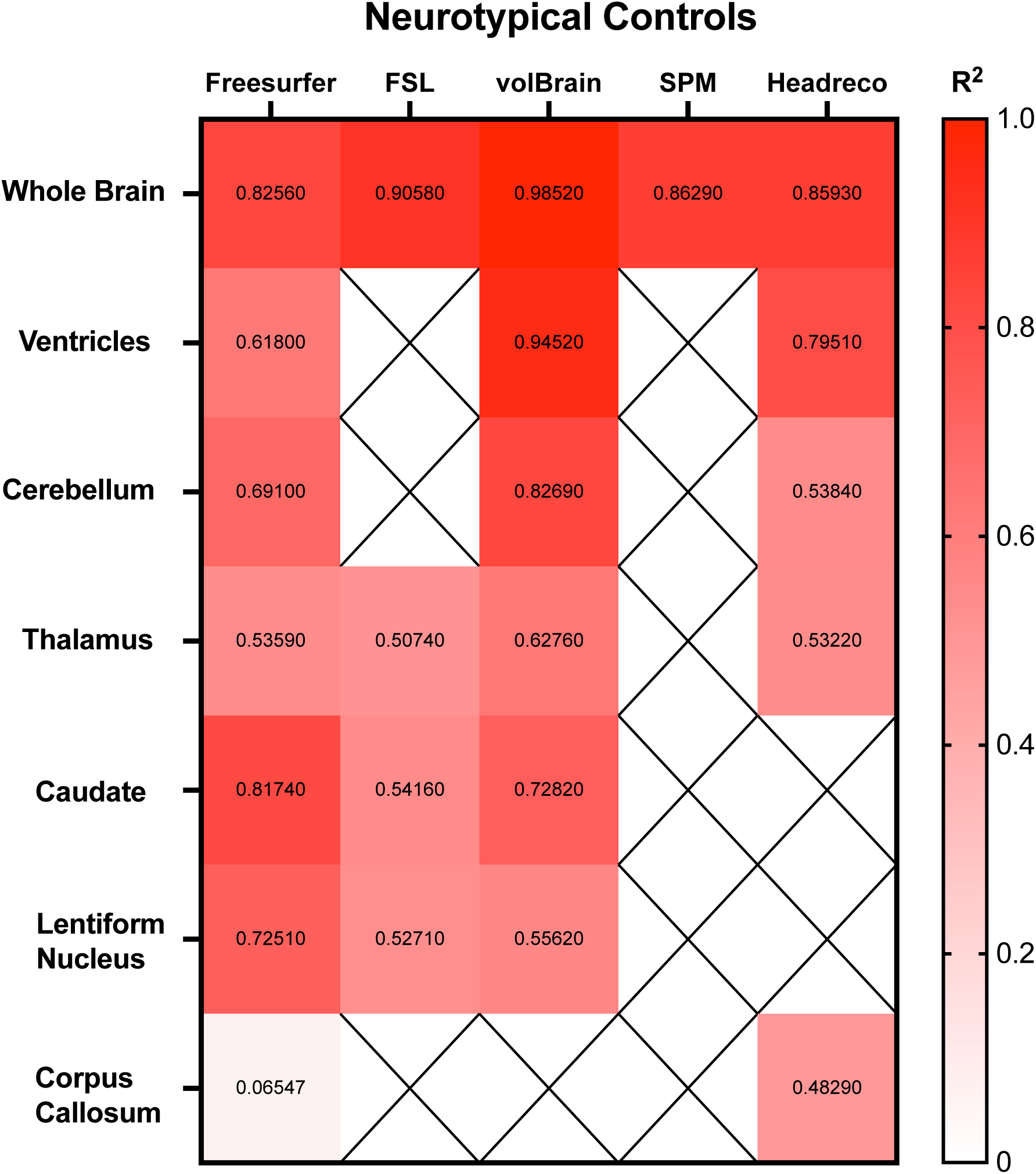
Heatmap of correlations strengths (R^2^) between the manual segmentation process and the 5 fully automated pipelines for the 7 structures of interest in neurotypical controls. N/A are designations where the region was not calculated using the specified segmentation algorithm.

### Ventricle Volume

Manual ventricle volumetric segmentation demonstrated higher ventricle volume in late-infantile GM1 patients when compared to neurotypical controls (*t*(12) = 2.98, *p* = 0.0115, Figure 8). Freesurfer, volBrain, and Headreco were also able to demonstrate this difference finding enlargement of the ventricles in late-infantile GM1 patients (Figure 8). In juvenile GM1 patients, no statistical difference was found in ventricle volume when compared to neurotypical controls using manual segmentation (*t*(19) = 2.98, *p* = 0.0115, Supplement Figure F1). Freesurfer, volBrain, and Headreco also did not find a statistical difference in ventricle volume between juvenile GM1 patients and neurotypical controls (Supplement Figure F1).

Correlations between the three fully automated ventricle segmentation processes and our manual process showed volBrain (R^2^ = 0.9452) as the only pipeline with a R^2^ value above 0.90 in neurotypical controls (Figure 5). In juvenile GM1 patients, volBrain (R^2^ = 0.9782) and Freesurfer (R^2^ = 0.9765) both had R^2^ values above 0.90 (Figure 6). In late-infantile GM1 patients, volBrain (R^2^ = 0.9974) and Headreco (R^2^ = 0.9950) both had R^2^ above 0.90 when compared to the manual process (Figure 7).

**Figure 6.**
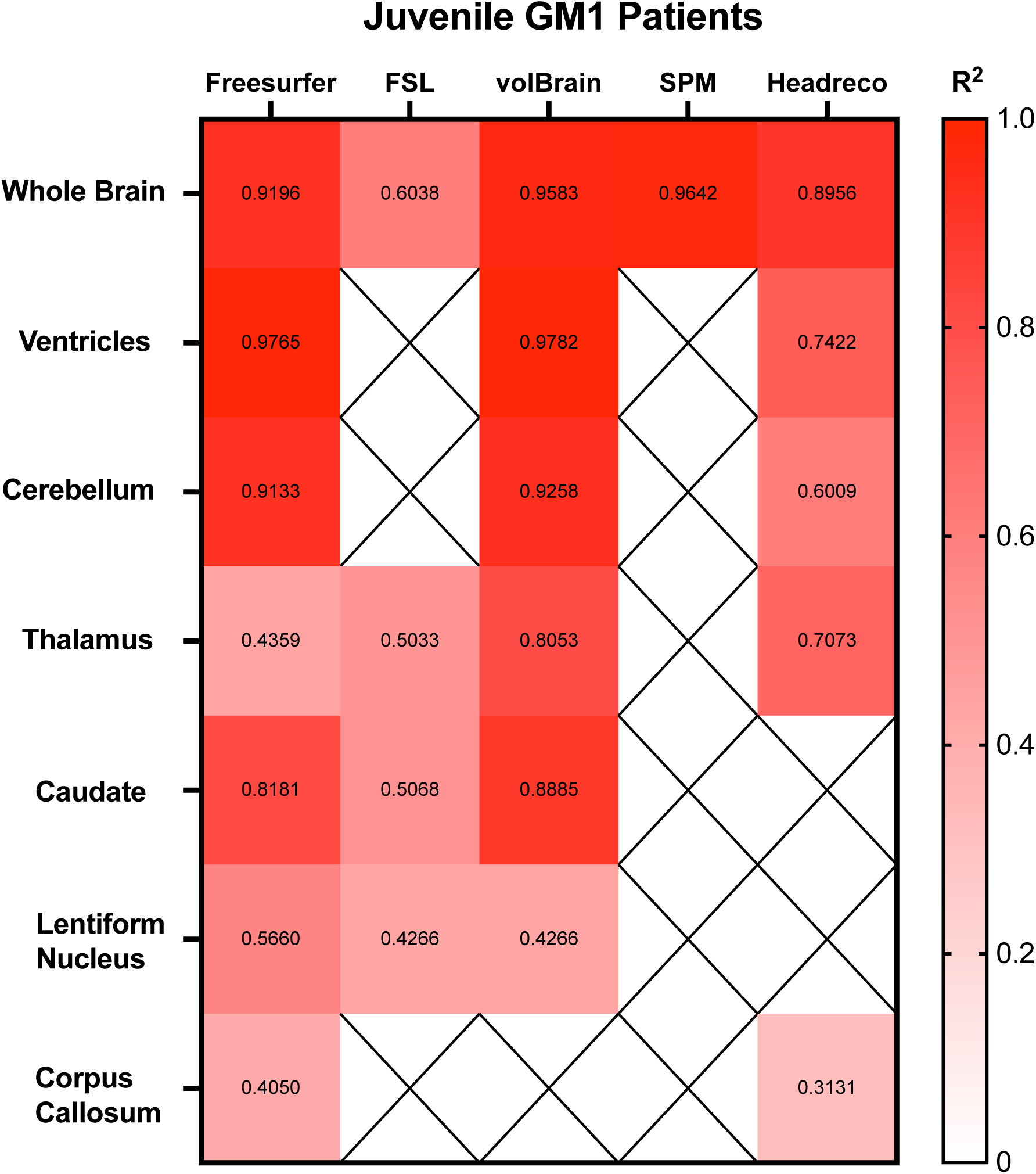
Heatmap of correlations strengths (R^2^) between the manual segmentation process and the 5 fully automated pipelines for the 7 structures of interest in juvenile (Juv) GM1 patients. N/A are designations where the region was not calculated using the specified segmentation algorithm.

**Figure 7.**
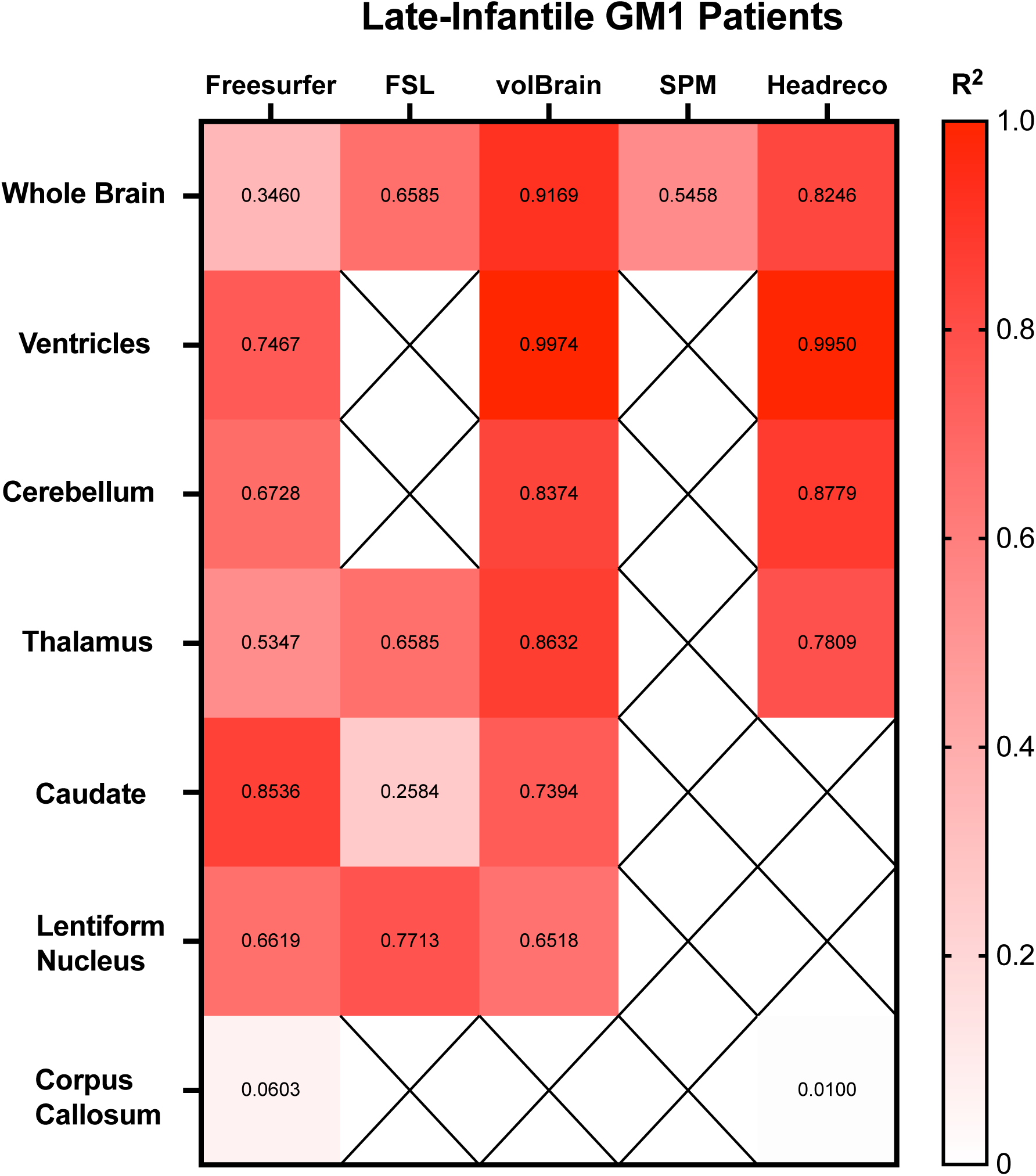
Heatmap of correlations strengths (R^2^) between the manual segmentation process and the 5 fully automated pipelines for the 7 structures of interest in late-infantile (LI) GM1 patients. N/A are designations where the region was not calculated using the specified segmentation algorithm.

### Cerebellar Volume

Manual cerebellar volume segmentation demonstrated higher cerebellar volume in neurotypical controls when compared to both late-infantile (t(12) = 3.32, *p* = 0.0061, Supplement Figure E1) and juvenile (*t*(19) = 2.69, *p* = 0.0146, Supplement Figure F2) GM1 patients. In late-infantile GM1 patients, Freesurfer, volBrain, and Headreco also demonstrated this result, finding larger cerebellar volume in neurotypical controls (Supplement Figure E1). In juvenile GM1 patients, Freesurfer and volBrain also demonstrated this result finding larger cerebellar volume in neurotypical controls, however Headreco did not find a statistical difference between juvenile GM1 patients and neurotypical controls (Supplement Figure F2).

Correlations between the three fully automated cerebellar segmentation processes and our manual process showed volBrain (R^2^ = 0.8259) as the only pipeline with a R^2^ value above 0.80 in neurotypical controls (Figure 5). In juvenile GM1 patients, volBrain (R^2^ = 0.9258) and Freesurfer (R^2^ = 0.9133) both had R^2^ values above 0.90 (Figure 6). In late-infantile GM1 patients, volBrain (R^2^ = 0.8374) and Headreco (R^2^ = 0.8779) both had R^2^ above 0.80 when compared to the manual process (Figure 7).

### Thalamic Volume

Manual thalamic volume segmentation demonstrated higher thalamic volume in neurotypical controls when compared to both late-infantile (t(12) = 6.14, *p* < 0.0001, Supplement Figure E2) and juvenile (t(19) = 3.51, *p* = 0.0023, Figure 9) GM1 patients. In late-infantile GM1 patients, Freesurfer, FSL, volBrain, and Headreco also demonstrated this result, finding larger thalamic volume in neurotypical controls (Supplement Figure E2). In juvenile GM1 patients, Freesurfer, volBrain, and Headreco also demonstrated this result finding larger thalamic volume in neurotypical controls, however FSL did not find a statistical difference between juvenile GM1 patients and neurotypical controls (Figure 9).

**Figure 8.**
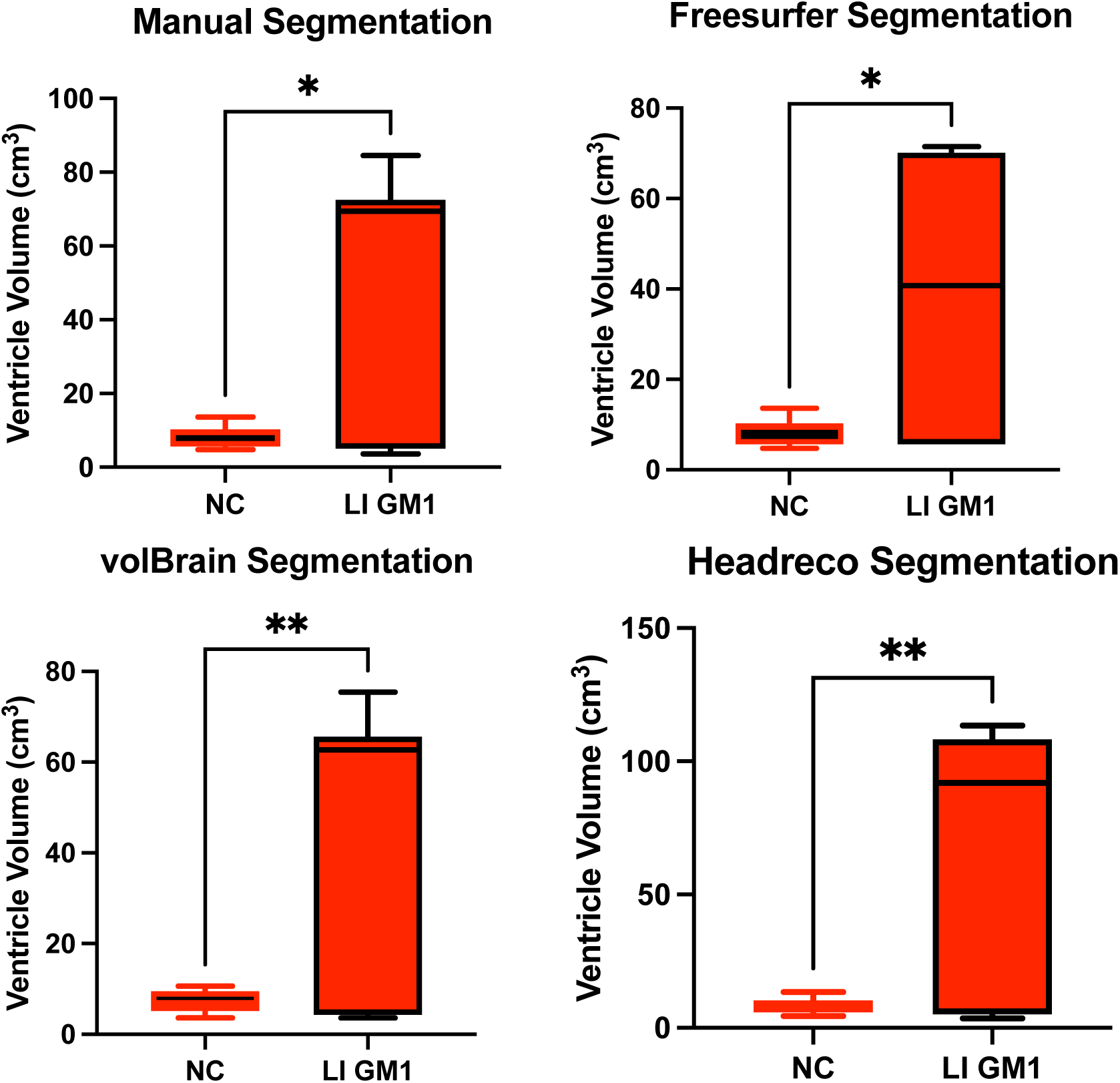
Cross-sectional evaluation of 3 automated segmentation algorithms to demonstrate cohort differences in ventricle volume. Late-infantile (LI) GM1 patients are shown in red. Neurotypical controls (NC) are shown in black. *P*-values were calculated from the *t*-statistic. * *P* < 0.05, ** *P* < 0.01, *** *P* < 0.001, **** *P* < 0.0001.

**Figure 9.**
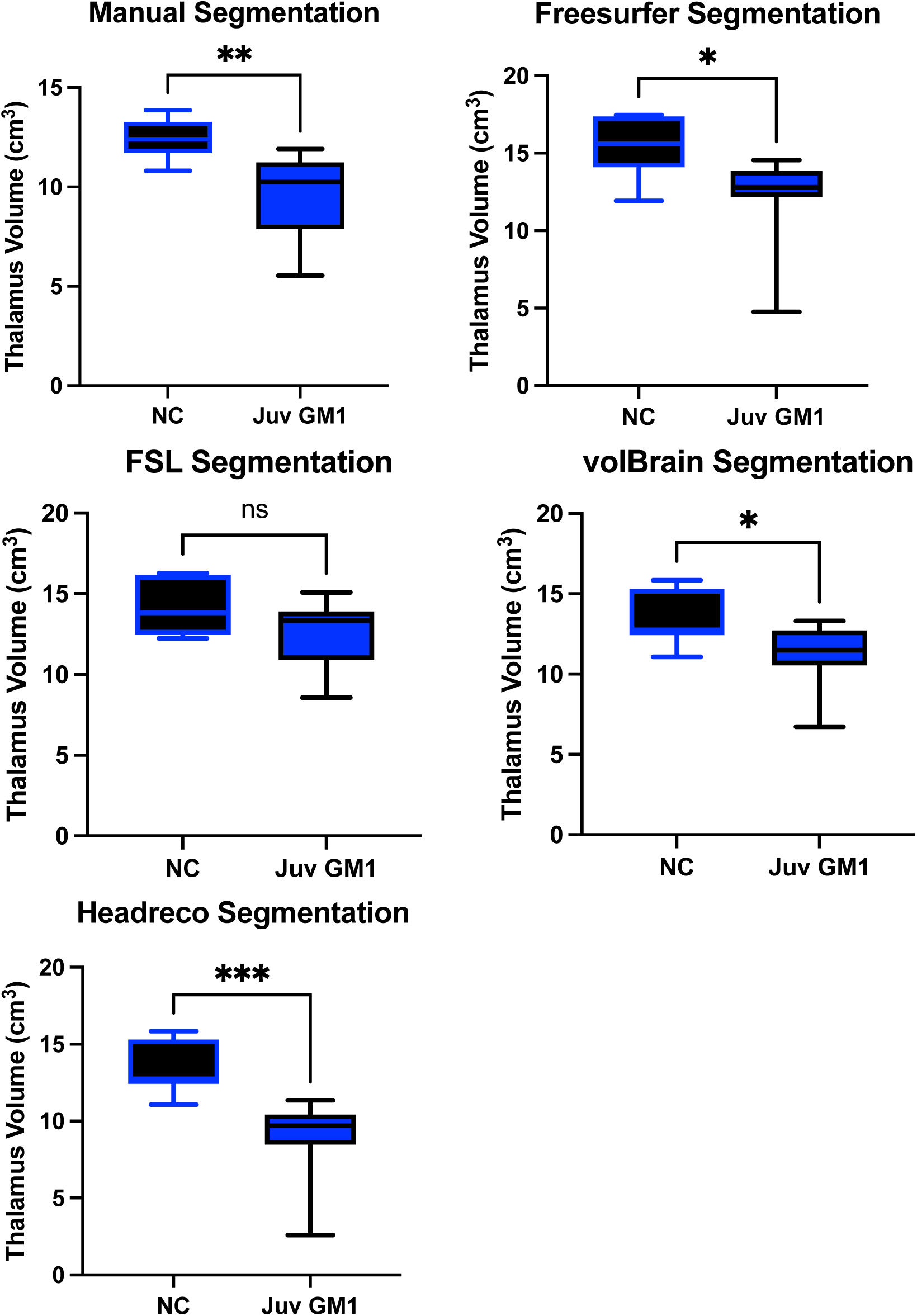
Cross-sectional evaluation of 4 automated segmentation algorithms ability to demonstrate cohort differences in thalamic volume. Juvenile (Juv) GM1 patients are shown in blue. Neurotypical controls (NC) are shown in black. *P*-values were calculated from the *t*-statistic. * *P* < 0.05, ** *P* < 0.01, *** *P* < 0.001, **** *P* < 0.0001.

Correlations between the four fully automated thalamic segmentation processes and our manual process showed volBrain (R^2^ = 0.6276) with the highest R^2^ value neurotypical controls (Figure 5). In juvenile GM1 patients, volBrain (R^2^ = 0.8053) again had the highest R^2^ value (Figure 6). In late-infantile GM1 patients, volBrain (R^2^ = 0.8632) again had the highest R^2^ value when compared to the manual process (Figure 7).

### Caudate Volume

Manual caudate volume segmentation demonstrated higher thalamic volume in neurotypical controls when compared to both late-infantile (t(12) = 2.72, *p* = 0.0185, Supplement Figure E3) and juvenile (t(19) = 6.00, *p* < 0.0001, Supplement Figure F3) GM1 patients. In late-infantile GM1 patients, Freesurfer, FSL, and volBrain also demonstrated this result, finding larger caudate volume in neurotypical controls (Supplement Figure E3). In juvenile GM1 patients, Freesurfer also demonstrated this result finding larger caudate volume in neurotypical controls, however FSL and volBrain did not find a statistical difference between juvenile GM1 patients and neurotypical controls (Supplement Figure F3).

Correlations between the three fully automated caudate nucleus segmentation processes and our manual process showed Freesurfer (R^2^ = 0.8174) with the highest R^2^ value in neurotypical controls (Figure 5). In juvenile GM1 patients, volBrain (R^2^ = 0.8885) had the highest R^2^ value (Figure 6). In late-infantile GM1 patients, Freesurfer (R^2^ = 0.8536) again had the highest R^2^ value when compared to the manual process (Figure 7).

### Lentiform Nucleus Volume

Manual caudate volume segmentation demonstrated higher thalamic volume in neurotypical controls when compared to both late-infantile (t(12) = 4.46, *p* = 0.0008, Supplement Figure E4) and juvenile (t(19) = 7.51, *p* < 0.0001, Supplement Figure F4) GM1 patients. In late-infantile GM1 patients, Freesurfer, FSL, and volBrain also demonstrated this result, finding larger lentiform nucleus volume in neurotypical controls (Supplement Figure E4). In juvenile GM1 patients, Freesurfer, FSL, and volBrain also demonstrated this result finding larger lentiform nucleus volume in neurotypical controls (Supplement Figure F4).

Correlations between the three fully automated lentiform nucleus segmentation processes and our manual process showed Freesurfer (R^2^ = 0.7251) with the highest R^2^ value in neurotypical controls (Figure 5). In juvenile GM1 patients, volBrain (R^2^ = 0.6605) had the highest R^2^ value (Figure 6). In late-infantile GM1 patients (Figure 7), Freesurfer (R^2^ = 0.6619) again had the highest R^2^ value when compared to the manual process. In late-infantile GM1 patients, volBrain had a similar correlation coefficient strength to Freesurfer (0.6518).

### Corpus Callosum Volume

Manual corpus callosum volume segmentation demonstrated higher corpus callosum volume in neurotypical controls when compared to both late-infantile (t(12) = 4.46, *p* = 0.0008, Supplement Figure E5) and juvenile (t(19) = 2.49, *p* = 0.0222, Supplement Figure F5) GM1 patients. In late-infantile GM1 patients, Freesurfer also demonstrated this result, finding larger corpus callosum volume in neurotypical controls (Supplement Figure E5), however Headreco did not demonstrate this difference. In juvenile GM1 patients, Headreco also demonstrated this result finding larger corpus callosum volume in neurotypical controls (Supplement Figure F5), however Freesurfer did not demonstrate this difference.

Correlations between the two fully automated corpus callosum segmentation processes and our manual process showed SimNIBS’ Headreco (R^2^ = 0.4829) with the highest R^2^ value neurotypical controls (Figure 5). In juvenile GM1 patients, Freesurfer (R^2^ = 0.4050) again had the highest R^2^ value (Figure 6). In late-infantile GM1 patients, both Freesurfer and Headreco had weak correlations (R^2^ < 0.1) with the manual process (Figure 7).

### Extended Results

Individual figures for the comparisons between late-infantile and juvenile with neurotypical controls for the remaining analyzed structures are included in sections E and F of the Supplementary materials, respectively. Individual figures for the correlations between each of the 5 automated pipelines with the manual process for all analyzed structures are included in sections H-L of the Supplementary Materials. Slope estimates and y-intercept values with standard errors for the correlations between each of the 5 automated pipelines with the manual process are included in section M of the Supplementary Materials.

## Discussion

This study assessed the capabilities of 5 fully automated brain MRI segmentation pipelines compared to a manual approach in juvenile and infantile GM1 gangliosidosis patients and neurotypical controls. We aimed to demonstrate the accuracy of the fully automated techniques, which have significant time saving advantages and can enable the analysis of large-scale datasets and larger complex brain structures including the cerebral white matter. We found our automated results to closely reflect those found by the manual approach suggesting the utility of fully automated processes in neurodegenerative diseases.

We first compared each automated segmentation pipeline’s ability to differentiate between GM1 patients and neurotypical controls using the 7 brain structures of interest. The manual segmentation process found statistically significant differences in 6 out of the 7 brain structures between juvenile GM1 patients and neurotypical controls. For the specific segmentations that were performed by each of the fully automated pipelines, statistically significant differences in brain structure volume were observed in 5 out of 6 for Freesurfer, 4 out of 5 for volBrain, 1 out of 4 for FSL, 1 out of 1 for SPM12, and 2 out of 4 for Headreco. In late-infantile patients, the manual segmentation process found statistically significant differences in all 7 brain structures. Freesurfer, volBrain, FSL, SPM12, and Headreco identified statistically significant differences in 6 out of 7, 6 out of 6, 3 out of 4, 1 out of 1, and 4 out of 5 brain structure volumes, respectively.

Correlation measurements were then performed comparing the volumetric results from each of the fully automated pipelines with the results from the manual approach to understand how they relate to a clinician’s measurements. volBrain showed the strongest correlation with the manual pipeline for the whole brain, ventricles, and thalamus for all three cohorts (Figures 5, 6, and 7). volBrain also showed the strongest correlation in the cerebellum in both the neurotypical controls and juvenile GM1 patients, with Headreco demonstrating the strongest correlation in the late-infantile GM1 patients. Freesurfer showed the strongest correlation with the manual pipeline for the caudate nucleus in neurotypical controls and late-infantile GM1 patients, with volBrain demonstrating the strongest correlation in the juvenile GM1 patients. Freesurfer also showed the strongest correlation for the lentiform nucleus in the neurotypical controls, with FSL demonstrating the strongest correlation in the late-infantile GM1 patients, and volBrain demonstrating the strongest correlation in the juvenile GM1 patients. Overall, our results suggest that among the 5 automated pipelines, volBrain is the most accurate for analyzing ventricle size, total brain volume, and thalamic volume in GM1 patients while Freesurfer is the most accurate for analyzing the caudate nucleus.

Corpus callosum segmentation in this study resulted in varying success. On the one hand, Freesurfer was able to demonstrate the correct positive relationship in the late-infantile GM1 patients and Headreco in juvenile GM1 patients. However, correlations with the manual process were weak and highly variable for all three cohorts. Furthermore, variability in the measured size was also observed. Headreco appears to measure further into the bilateral hemispheres while Freesurfer appears to exclude relevant regions that is included in our manual process. This result is consistent with previous analysis which suggests that a dedicated corpus callosum segmentation process may be justified for future studies.^57–59^

Limitations of this study need to be considered before the results are used to guide clinical practice. First, this study is limited by a small sample size in neurotypical controls (n = 11, 19 MRI scans), juvenile GM1 patients (n = 16, 45 MRI scans), and late-infantile patients (n = 8, 11 MRI scans). However, this study represents the largest MRI dataset of Type II GM1 gangliosidosis patients.^60^ Second, this study is limited by the variations in scanners and protocols used between the GM1 patients and neurotypical controls. However, all 6 analysis techniques were given the same images to analyze, and the results of this study focus on the comparisons between these techniques. Ultimately, future investigations should utilize identical protocols when comparing MRI segmentation techniques. Dice similarity coefficients, Hausdorff distance, and the mean distance to agreement were also not considered in this study.^61–63^ Future studies investigating the accuracy of automated MRI segmentation methods should incorporate these metrics, however in this pilot study we focused on the capability of these methods for demonstrating GM1 associated neurodegeneration compared to neurotypical controls.

### Conclusion

In this study, we compared 5 fully automated segmentation pipelines with a manual pipeline in neurotypical controls and juvenile and late-infantile GM1 gangliosidosis patients. We analyzed 7 brain structures of interest including volumes of the total brain, cerebellum, ventricles, bilateral thalami, bilateral lentiform nucleus, bilateral caudate nucleus, and corpus callosum. We found that a combination of pipelines led to the strongest correlations with the manual pipeline with volBrain’s *vol2Brain* demonstrating the strongest correlations for the whole brain, ventricles, and thalamus, Freesurfer’s *recon-all* demonstrating the strongest correlations for the caudate nucleus, and a combination of pipelines demonstrating the strongest correlations for the cerebellum and lentiform nucleus. Ultimately, we found our fully automated results to be consistent with our manual process, however further studies are needed to investigate other neurodegenerative and lysosomal storage disorders.

## Data Availability

The data described in this manuscript are available from the corresponding author upon reasonable request. Neuroimaging data for the early childhood neurotypical control group are publicly available here: https://osf.io/axz5r/.^33^ Neuroimaging data for the adolescent neurotypical control group are publicly available here: https://doi.org/10.6084/m9.figshare.6002273.v2.34

## Acknowledgements

We thank the participants and their families for the generosity of their time and efforts. Volumetric MRI analysis in this work utilized the computational resources of the Biowulf Linux cluster at the National Institutes of Health (http://hpc.nih.gov).

## Authors’ Contribution

CJL, WAG, MSS, CJT, and MTA contributed to the study conception and design. Data collection was performed by CJL, JMJ, PD, WAG, CJT, and MTA. Figure layout was performed by CJL, WAG, MSS, CJT, and MTA. Bioinformatic and imaging analysis was performed by CJL, JK, CZ, ZV, ALK, ZSR, MSS, CJT, and MTA. The first draft of the manuscript was written by CJL, MTA, and CJT, and all authors commented on previous versions of the manuscript.

## Ethics Declaration

The NIH Institutional Review Board approved this protocol (02-HG-0107). Informed consent was completed with parents or legal guardians of the patients. All participants were assessed for their ability to provide assent; none were deemed capable.

## Consent for Publication

Not applicable.

## Supplementary Methods

### Supplement A: Natural History Study Participant Characteristics^1^

**Table A1.**
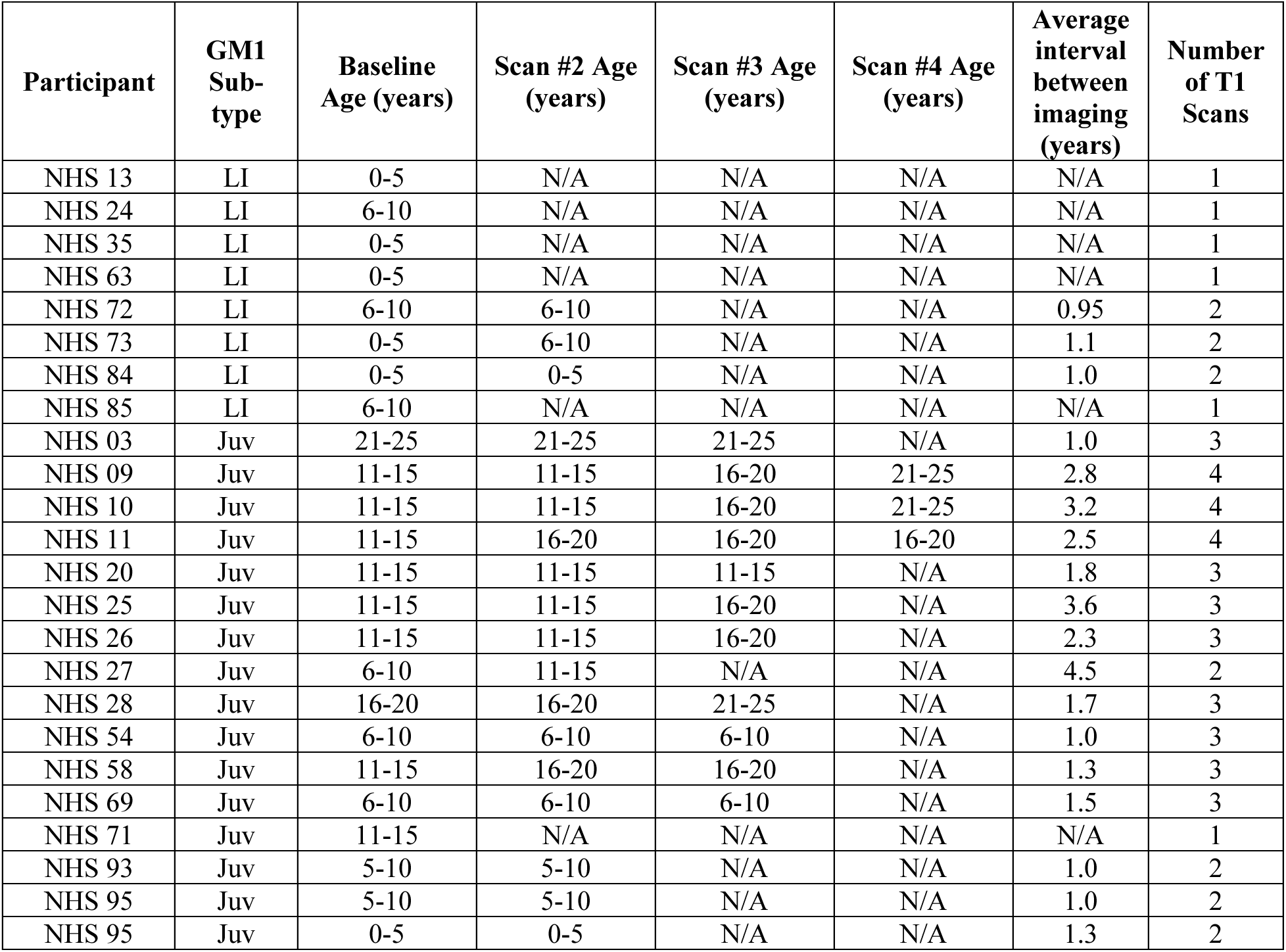
Natural History Study T1-Weighted MRI Cohort (n = 24), specific ages redacted per MedArXiv requirements

| Participant | GM1 Sub-type | Baseline Age (years) | Scan #2 Age (years) | Scan #3 Age (years) | Scan #4 Age (years) | Average interval between imaging (years) | Number of T1 Scans |
| --- | --- | --- | --- | --- | --- | --- | --- |
| NHS 13 | LI | 0-5 | N/A | N/A | N/A | N/A | 1 |
| NHS 24 | LI | 6-10 | N/A | N/A | N/A | N/A | 1 |
| NHS 35 | LI | 0-5 | N/A | N/A | N/A | N/A | 1 |
| NHS 63 | LI | 0-5 | N/A | N/A | N/A | N/A | 1 |
| NHS 72 | LI | 6-10 | 6-10 | N/A | N/A | 0.95 | 2 |
| NHS 73 | LI | 0-5 | 6-10 | N/A | N/A | 1.1 | 2 |
| NHS 84 | LI | 0-5 | 0-5 | N/A | N/A | 1.0 | 2 |
| NHS 85 | LI | 6-10 | N/A | N/A | N/A | N/A | 1 |
| NHS 03 | Juv | 21-25 | 21-25 | 21-25 | N/A | 1.0 | 3 |
| NHS 09 | Juv | 11-15 | 11-15 | 16-20 | 21-25 | 2.8 | 4 |
| NHS 10 | Juv | 11-15 | 11-15 | 16-20 | 21-25 | 3.2 | 4 |
| NHS 11 | Juv | 11-15 | 16-20 | 16-20 | 16-20 | 2.5 | 4 |
| NHS 20 | Juv | 11-15 | 11-15 | 11-15 | N/A | 1.8 | 3 |
| NHS 25 | Juv | 11-15 | 11-15 | 16-20 | N/A | 3.6 | 3 |
| NHS 26 | Juv | 11-15 | 11-15 | 16-20 | N/A | 2.3 | 3 |
| NHS 27 | Juv | 6-10 | 11-15 | N/A | N/A | 4.5 | 2 |
| NHS 28 | Juv | 16-20 | 16-20 | 21-25 | N/A | 1.7 | 3 |
| NHS 54 | Juv | 6-10 | 6-10 | 6-10 | N/A | 1.0 | 3 |
| NHS 58 | Juv | 11-15 | 16-20 | 16-20 | N/A | 1.3 | 3 |
| NHS 69 | Juv | 6-10 | 6-10 | 6-10 | N/A | 1.5 | 3 |
| NHS 71 | Juv | 11-15 | N/A | N/A | N/A | N/A | 1 |
| NHS 93 | Juv | 5-10 | 5-10 | N/A | N/A | 1.0 | 2 |
| NHS 95 | Juv | 5-10 | 5-10 | N/A | N/A | 1.0 | 2 |
| NHS 95 | Juv | 0-5 | 0-5 | N/A | N/A | 1.3 | 2 |

### Supplement B: Age-Matched Methodology for Late-Infantile Patients

To evaluate the 5 automated segmentation pipelines’ ability to demonstrate volumetric differences between the late-infantile patients and neurotypical controls, we performed a cross-sectional analysis including age and sex-matched data (Table B1 and B2). Seven late-infantile patients’ (4 females, 3 males) baseline MRI scans and 7 neurotypical control (4 females, 3 males) scans from the Calgary Preschool^2^ and Adolescent^3^ databases were included in this analysis.

**Table B1.** Late-infantile Age Matched Cohort

| Participant | Sex | Age |
| --- | --- | --- |
| NHS 13 | F | 0-5 |
| NHS 35 | F | 0-5 |
| NHS 63 | M | 0-5 |
| NHS 72 | M | 6-10 |
| NHS 73 | M | 0-5 |
| NHS 84 | F | 0-5 |
| NHS 85 | F | 6-10 |
| | | $5.3 \pm 1.9$ |

**Table B2.** Late-infantile Age Matched Cohort of Neurotypical Controls

| Participant | Dataset | Sex | Age |
| --- | --- | --- | --- |
| Calgary 10014 | Preschool | M | 0-5 |
| Calgary 10027 | Preschool | F | 0-5 |
| Calgary 10044 | Preschool | M | 0-5 |
| Calgary 10064 | Preschool | F | 6-10 |
| Calgary 15003 | Adolescent | F | 6-10 |
| Calgary 15008 | Adolescent | F | 11-15 |
| Calgary 15010 | Adolescent | M | 6-10 |
| | | | $6.0 \pm 3.4$ |

**Figure B1.**
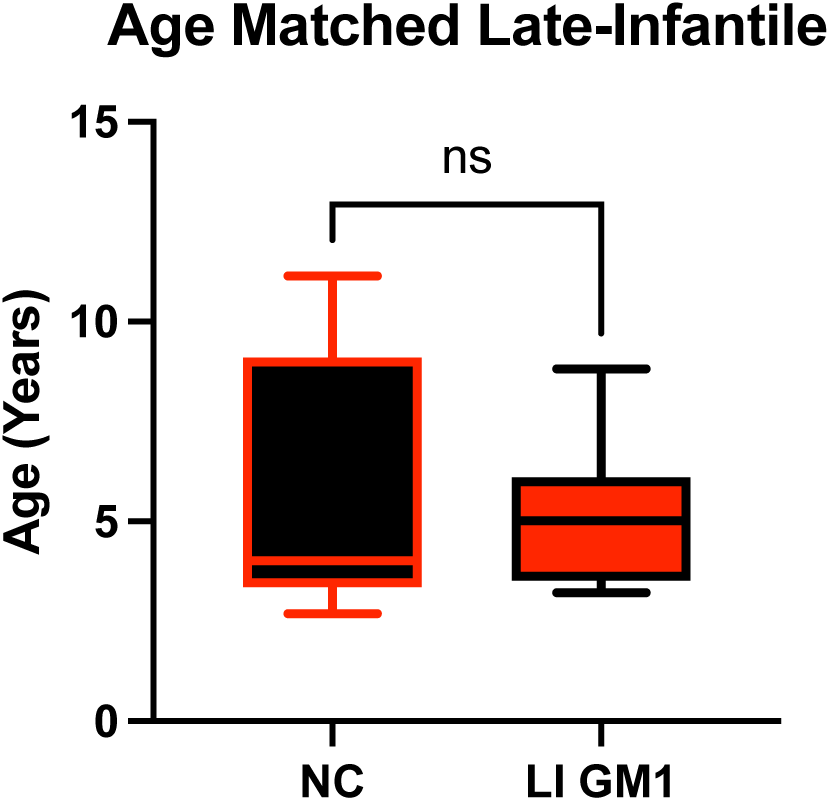
Age and sex-matched late-infantile GM1 gangliosidosis cohort evaluated for differences by a 2 tailed t-test (t=0.4771, df =12, *p* = 0.6418).

### Supplement C: Age-Matched Methodology for Juvenile Patients

To evaluate the 5 automated segmentation pipelines’ ability to demonstrate volumetric differences between the juvenile patients and neurotypical controls, we performed a cross-sectional analysis including age and sex-matched data (Table C1 and C2). Fourteen juvenile patients’ (8 females, 6 males) baseline MRI scans and 7 neurotypical control (4 females, 3 males) scans from the Calgary Preschool^2^ and Adolescent^3^ databases were included in this analysis.

**Table C1.** Juvenile Age Matched Cohort

| Participant | Sex | Age |
| --- | --- | --- |
| NHS 09 | M | 11-15 |
| NHS 10 | F | 11-15 |
| NHS 11 | M | 11-15 |
| NHS 20 | F | 11-15 |
| NHS 25 | F | 11-15 |
| NHS 26 | M | 11-15 |
| NHS 27 | M | 6-10 |
| NHS 28 | F | 16-20 |
| NHS 54 | F | 6-10 |
| NHS 58 | F | 11-15 |
| NHS 69 | F | 6-10 |
| NHS 93 | F | 6-10 |
| NHS 94 | M | 6-10 |
| NHS 95 | M | 0-5 |
|  |  | 10.9 ± 4.1 |

**Table C2.** Juvenile Age Matched Cohort of Neurotypical Controls

| Participant | Dataset | Sex | Age |
| --- | --- | --- | --- |
| Calgary 10027 | Preschool | F | 0-5 |
| Calgary 14006 | Preschool | M | 6-10 |
| Calgary 14008 | Preschool | M | 11-15 |
| Calgary 14011 | Preschool | M | 11-15 |
| Calgary 14013 | Adolescent | F | 11-15 |
| Calgary 15003 | Adolescent | F | 6-10 |
| Calgary 15008 | Adolescent | F | 11-15 |
|  |  |  | 10.6 ± 2.9 |

**Figure C1.**
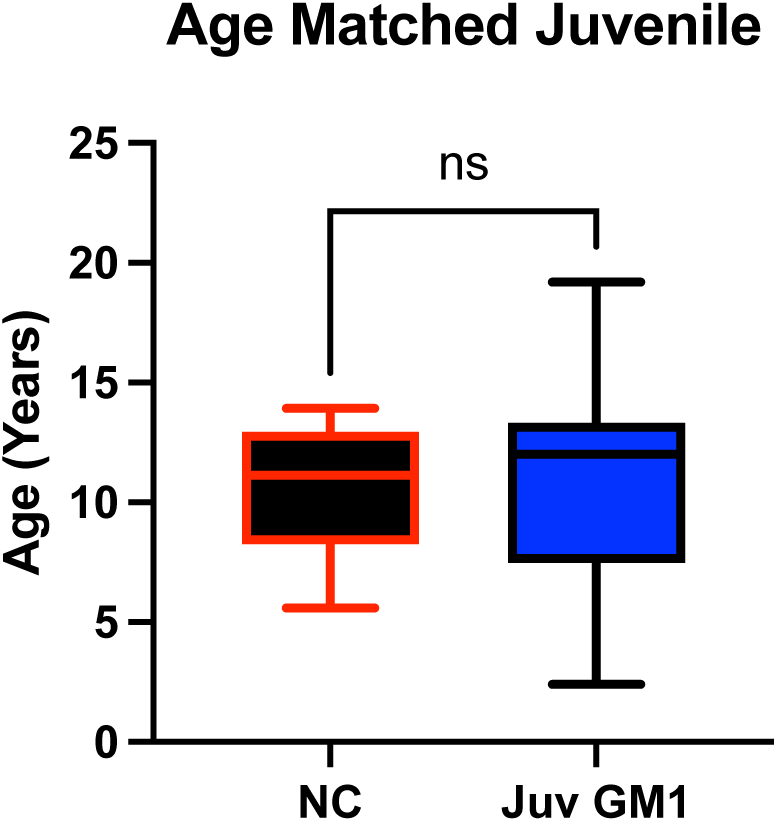
Age and sex-matched juvenile GM1 gangliosidosis cohort evaluated for differences by a 2 tailed *t*-test (*t*=0.1757, df =19, *p* = 0.8624).

### Supplement D: MRI Exclusion

#### Manual Segmentation Process

All 19 MRI scans from neurotypical controls, all 11 scans from 8 late-infantile GM1 gangliosidosis patients, and all 45 MRI scans from 16 juvenile GM1 gangliosidosis patients were analyzed using the manual segmentation method.

#### Freesurfer

All 19 MRI scans from 11 neurotypical controls were analyzed using the Freesurfer *recon-all* pipeline. 10 MRI scans from 7 late-infantile patients were analyzed using the Freesurfer *recon-all* pipeline, with one late-infantile patient MRI scan failing to process due to significant atrophy and demyelination (Figure D1). Thirty-nine MRI scans from 16 juvenile patients were analyzed using the Freesurfer *recon-all* pipeline with one 3 MRI scans failing to process due to the presence of bone marrow in the cerebrospinal fluid (Figure D2) and 3 MRI scans failing to process due to significant atrophy (Figure D3).

#### volBrain

All 19 MRI scans from 11 neurotypical controls were analyzed using volBrain’s *vol2Brain* pipeline. Ten MRI scans from 7 late-infantile patients were analyzed using the volBrain’s *vol2Brain* pipeline, with one late-infantile patient MRI scan failing to process due to significant atrophy and demyelination (Figure D1). All 45 MRI scans from 16 juvenile GM1 gangliosidosis patients were analyzed using the volBrain’s *vol2Brain* segmentation method.

#### FSL

##### FSL FAST

All 19 MRI scans from 11 neurotypical controls were analyzed using FSL’s *FAST* pipeline. Ten MRI scans from 7 late-infantile GM1 gangliosidosis patients were analyzed using FSL’s *FAST* pipeline with one late-infantile patient MRI scan failing to process due to significant atrophy and demyelination (Figure D1). All 45 MRI scans from 16 juvenile GM1 gangliosidosis patients were analyzed using the FSL’s *fast* segmentation method.

##### FSL FIRST

All 19 MRI scans from 11 neurotypical controls were analyzed using the FSL’s *FIRST* pipeline. 10 MRI scans from 7 late-infantile patients were analyzed using FSL’s *FIRST* with one late-infantile patient MRI scan failing to process due to significant atrophy (Figure D1). Forty-three MRI scans from 16 juvenile GM1 gangliosidosis patients were analyzed using the FSL *first* segmentation methods. Two MRI scans failed to process due to the presence of bone marrow in the cerebrospinal fluid (Figure D2) and significant atrophy (Figure D3, Scan3).

#### SPM12

All 19 MRI scans from 11 neurotypical controls were analyzed using the SPM12’s *Segment* pipeline. All 11 MRI scans from late-infantile GM1 gangliosidosis patients were analyzed using SPM12’s *Segment* pipeline. All 45 MRI scans from 16 juvenile GM1 gangliosidosis patients were analyzed using the SPM12’s *Segment* segmentation method.

#### SimNIBS’ Headreco

All 19 MRI scans from 11 neurotypical controls were analyzed using the SimNIBS’ *Headreco* pipeline. Ten MRI scans from 7 late-infantile patients were analyzed using the SimNIBS’ *Headreco* pipeline, with one late-infantile patient MRI scan failing to process due to significant atrophy and demyelination (Figure D1). Forty-three scans from 16 juvenile GM1 patients were analyzed using SimNIBS. Two scans from a juvenile participant of the GM1 gangliosidosis cohort were excluded from this analysis due to the presence of bone marrow in the cerebrospinal fluid (Figure D2).

**Figure D1.**
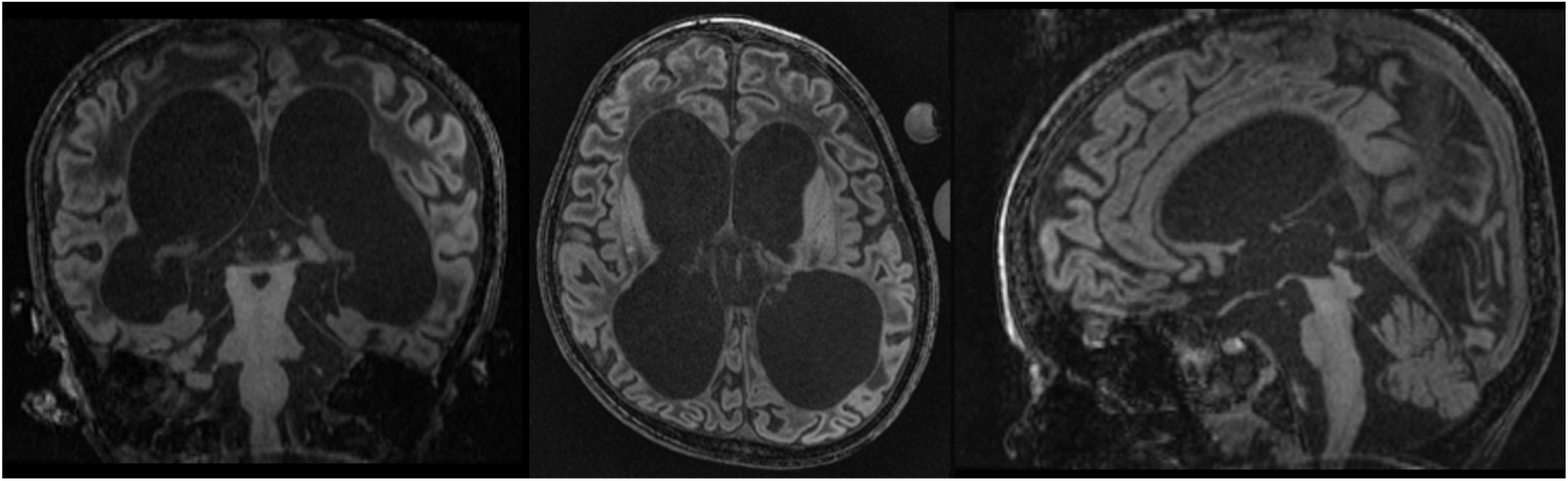
Coronal, Axial, and Sagittal T1-weighted MRI of a late-infantile GM1 patient with significant cerebral atrophy, ventricle enlargement, and hypomyelination.

**Figure D2.**
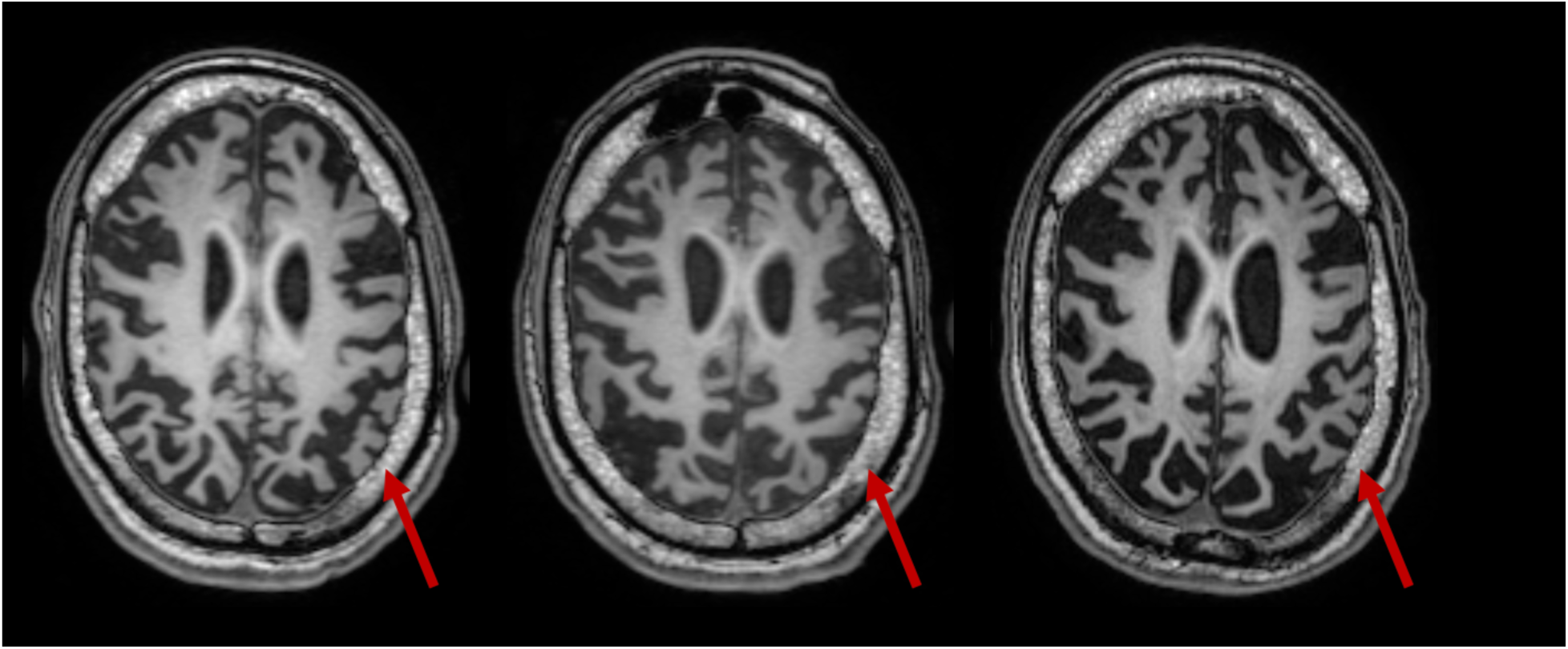
Serial T1 MRI of an excluded juvenile GM1 patient at age 11-15 (Scan 1), 16-20 (Scan 2), and 16-20 (Scan 3) years old with bone marrow (red arrow) in the cerebrospinal fluid.

**Figure D3.**
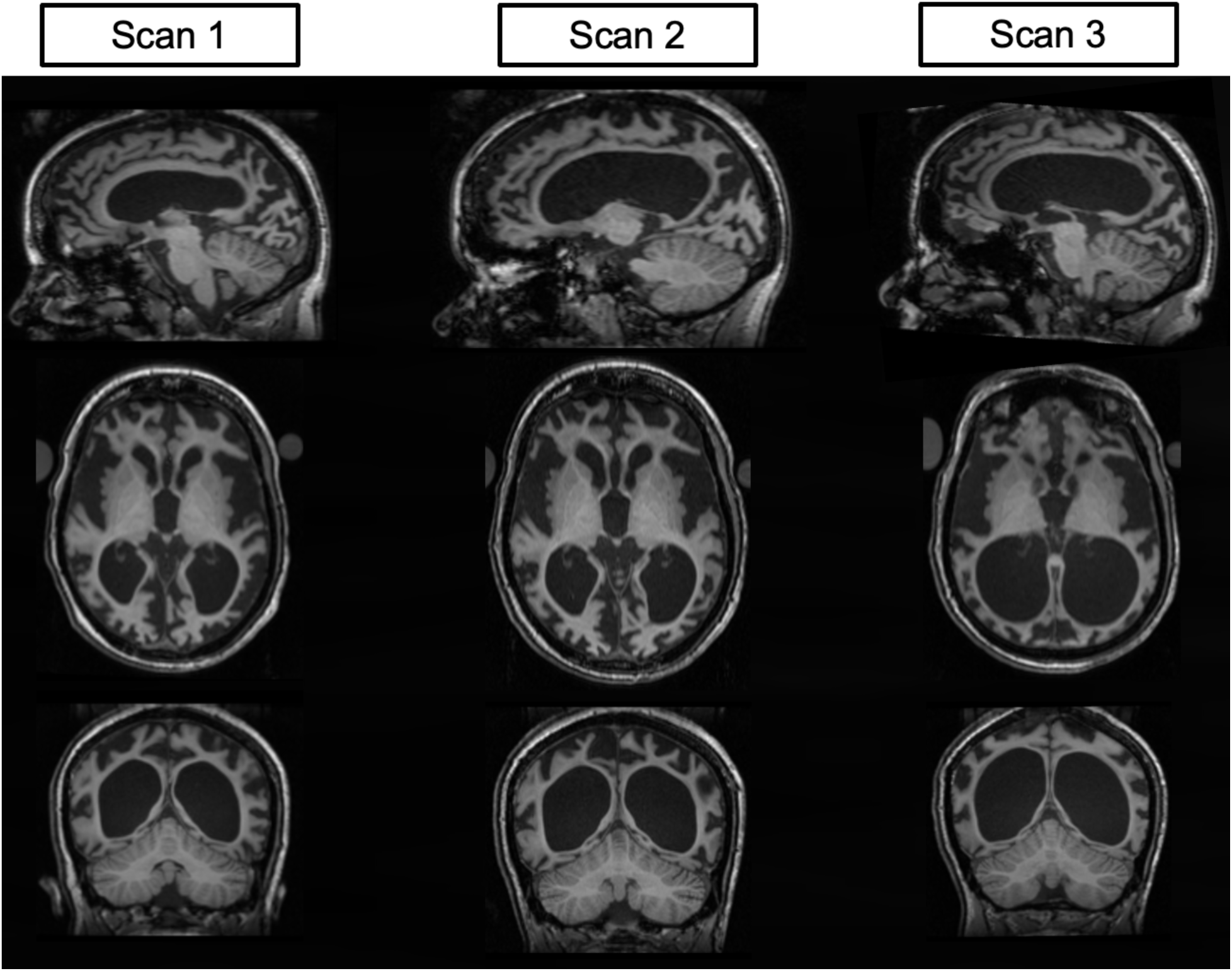
Serial T1 MRI imaging of a juvenile GM1 patient at age 16-20 (Scan 1), 16-20 (Scan 2), and 21-25 (Scan 3) years old with significant atrophy and ventricle enlargement across three evaluations.

## Supplementary Results

### Supplement E. Cross-sectional Analysis of Late-Infantile Patients

**Figure E1.**
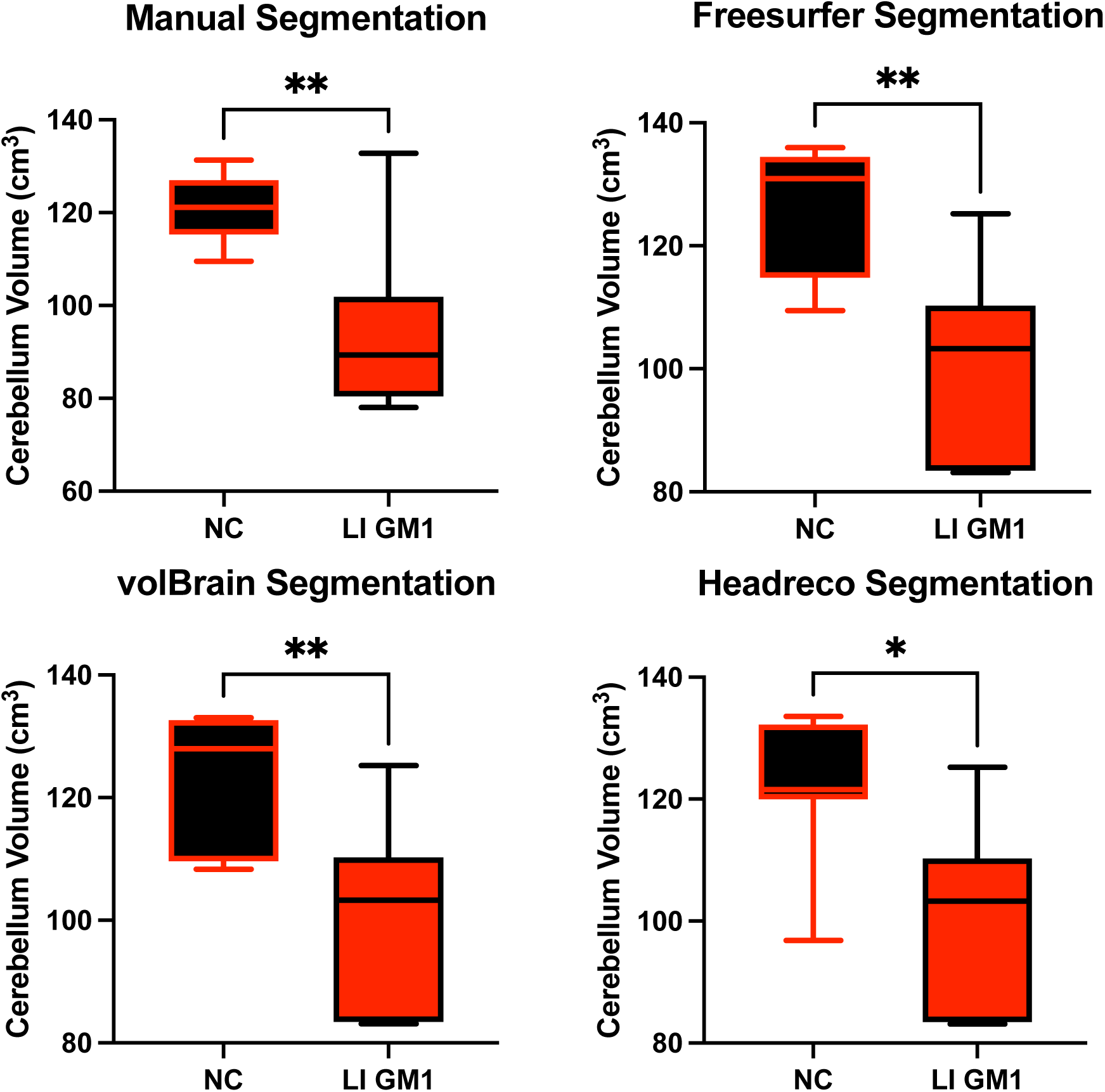
Cross-sectional evaluation of the 3 automated segmentation algorithms to demonstrate cohort differences in cerebellar volume. Late-infantile (LI) GM1 patients are shown in Red. Neurotypical controls (NC) are shown in black. *P*-values were calculated from the *t*-statistic. * *P* < 0.05, ** *P* < 0.01, *** *P* < 0.001, **** *P* < 0.0001.

**Figure E2.**
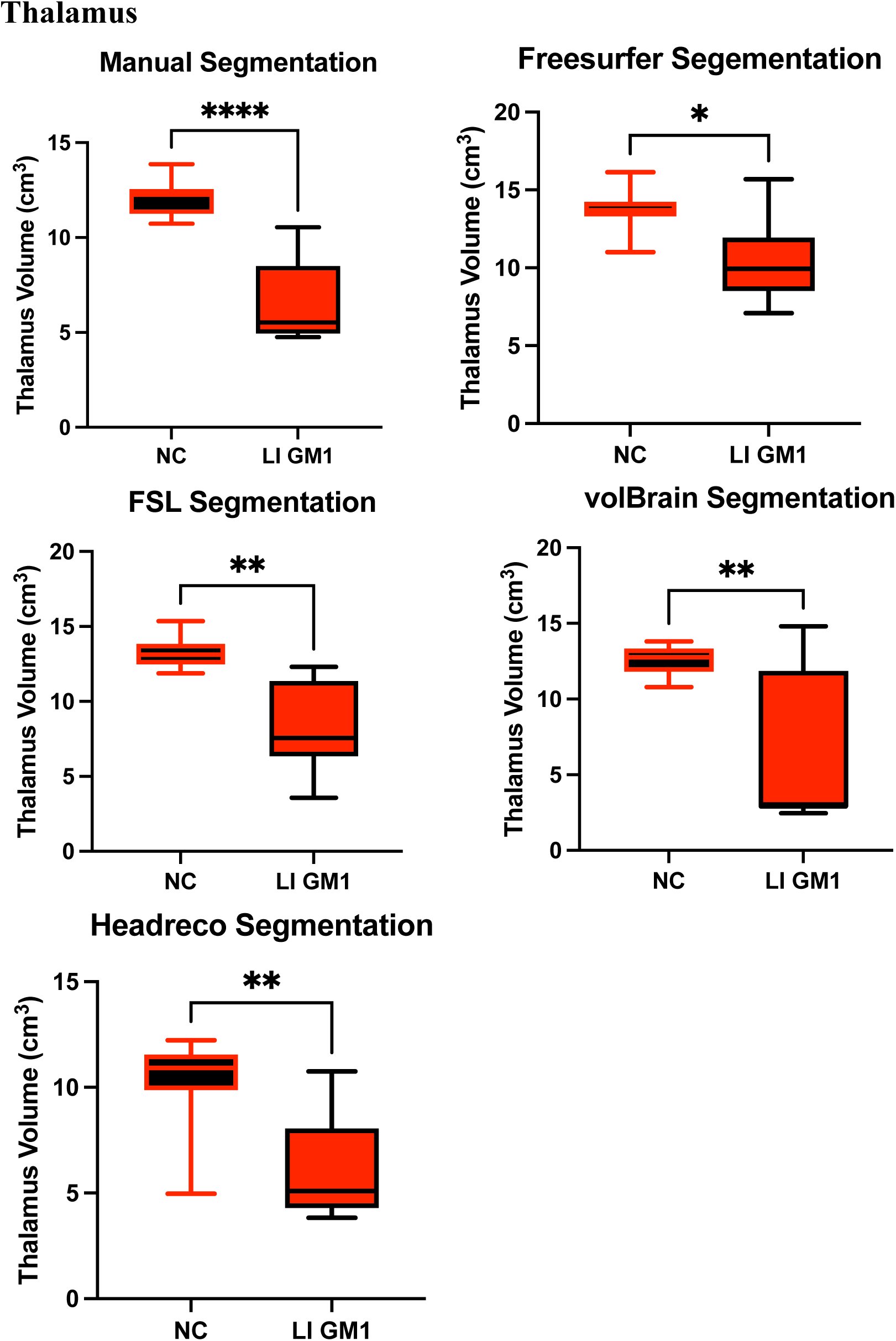
Cross-sectional evaluation of the 4 automated segmentation algorithms to demonstrate cohort differences in thalamic volume. Late-infantile (LI) GM1 patients are shown in Red. Neurotypical controls (NC) are shown in black. *P*-values were calculated from the *t*-statistic. * *P* < 0.05, ** *P* < 0.01, *** *P* < 0.001, **** *P* < 0.0001.

**Figure E3.**
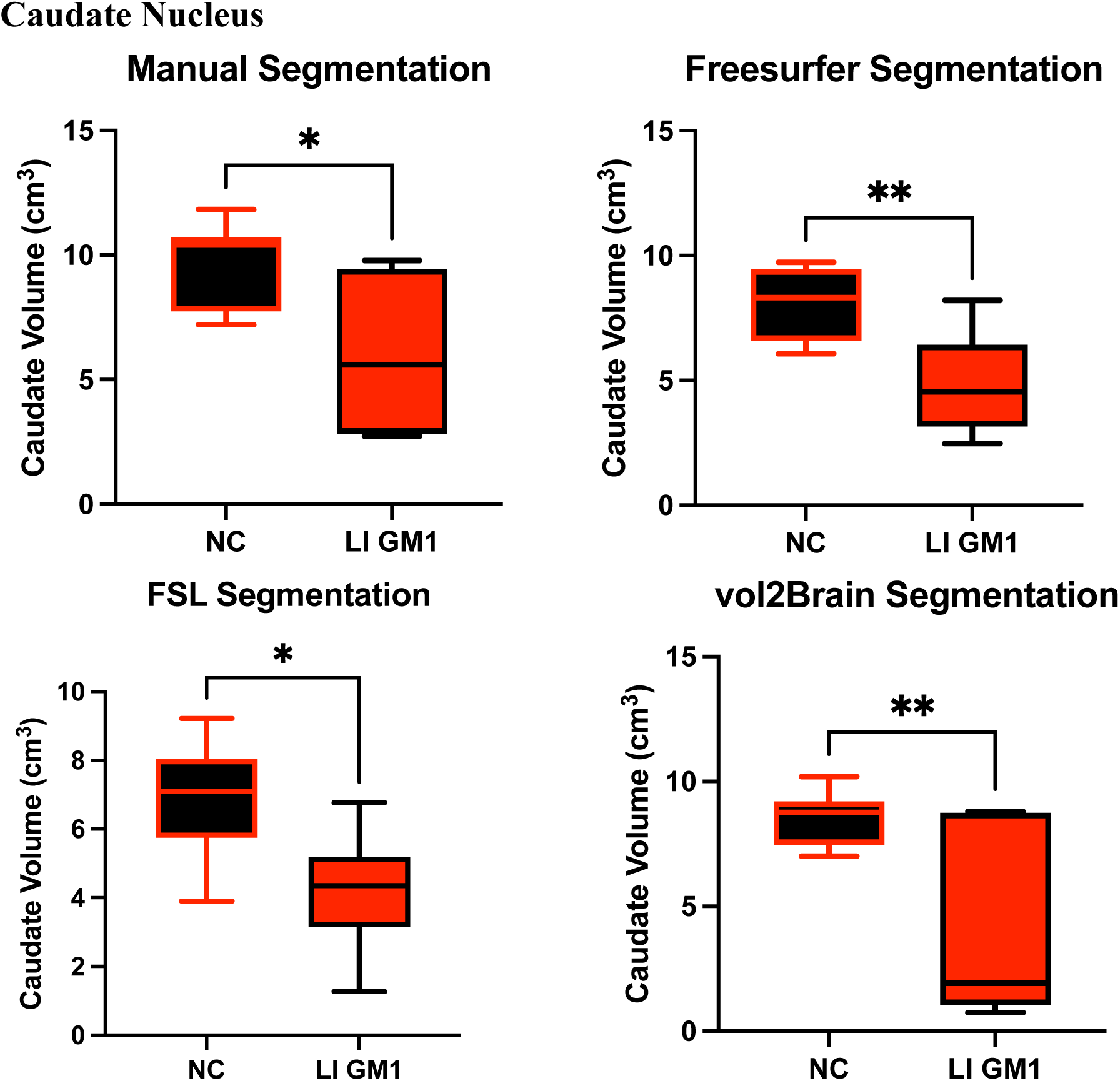
Cross-sectional evaluation of the 3 automated segmentation algorithms to demonstrate cohort differences in caudate volume. Late-infantile (LI) GM1 patients are shown in Red. Neurotypical controls (NC) are shown in black. *P*-values were calculated from the *t*-statistic. * *P* < 0.05, ** *P* < 0.01, *** *P* < 0.001, **** *P* < 0.0001.

**Figure E4.**
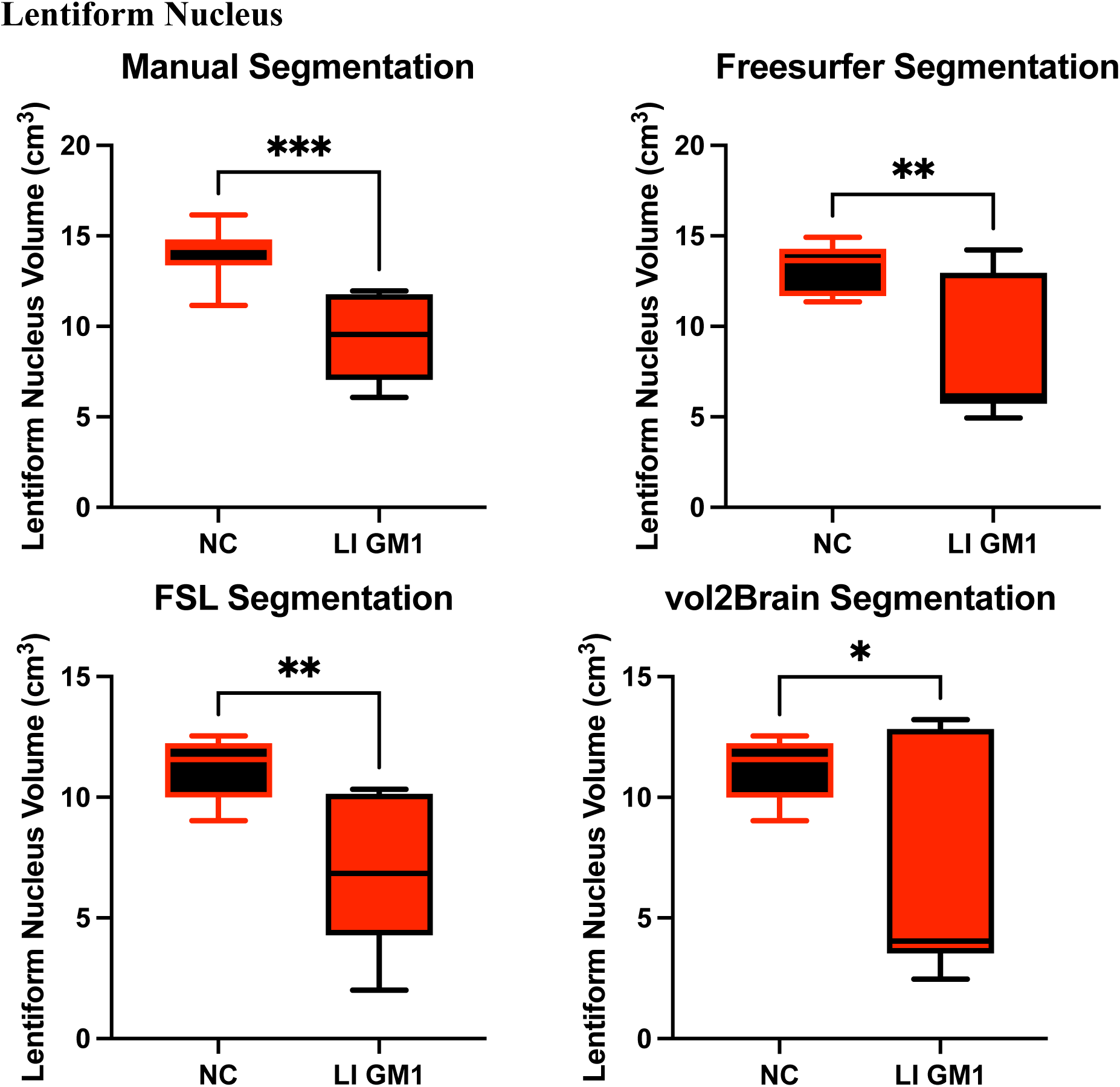
Cross-sectional evaluation of the 3 automated segmentation algorithms to demonstrate cohort differences in lentiform nucleus volume. Late-infantile (LI) GM1 patients are shown in Red. Neurotypical controls (NC) are shown in black. *P*-values were calculated from the *t*-statistic. * *P* < 0.05, ** *P* < 0.01, *** *P* < 0.001, **** *P* < 0.0001.

**Figure E5.**
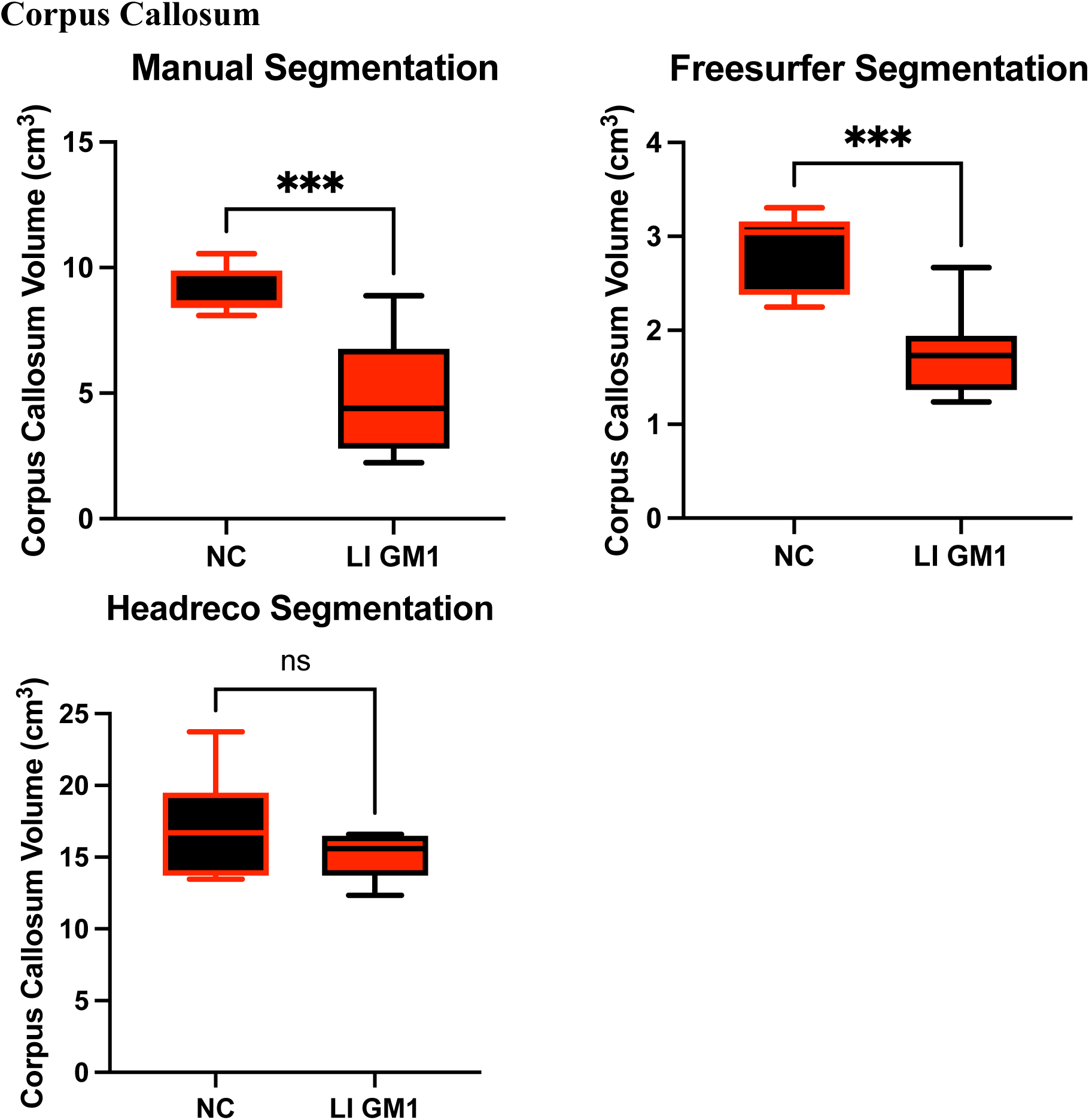
Cross-sectional evaluation of the 2 automated segmentation algorithms to demonstrate cohort differences in corpus callosum volume. Late-infantile (LI) GM1 patients are shown in Red. Neurotypical controls (NC) are shown in black. *P*-values were calculated from the *t*-statistic. * *P* < 0.05, ** *P* < 0.01, *** *P* < 0.001, **** *P* < 0.0001.

### Supplement F. Cross-sectional Analysis of Juvenile Patients

**Figure F1.**
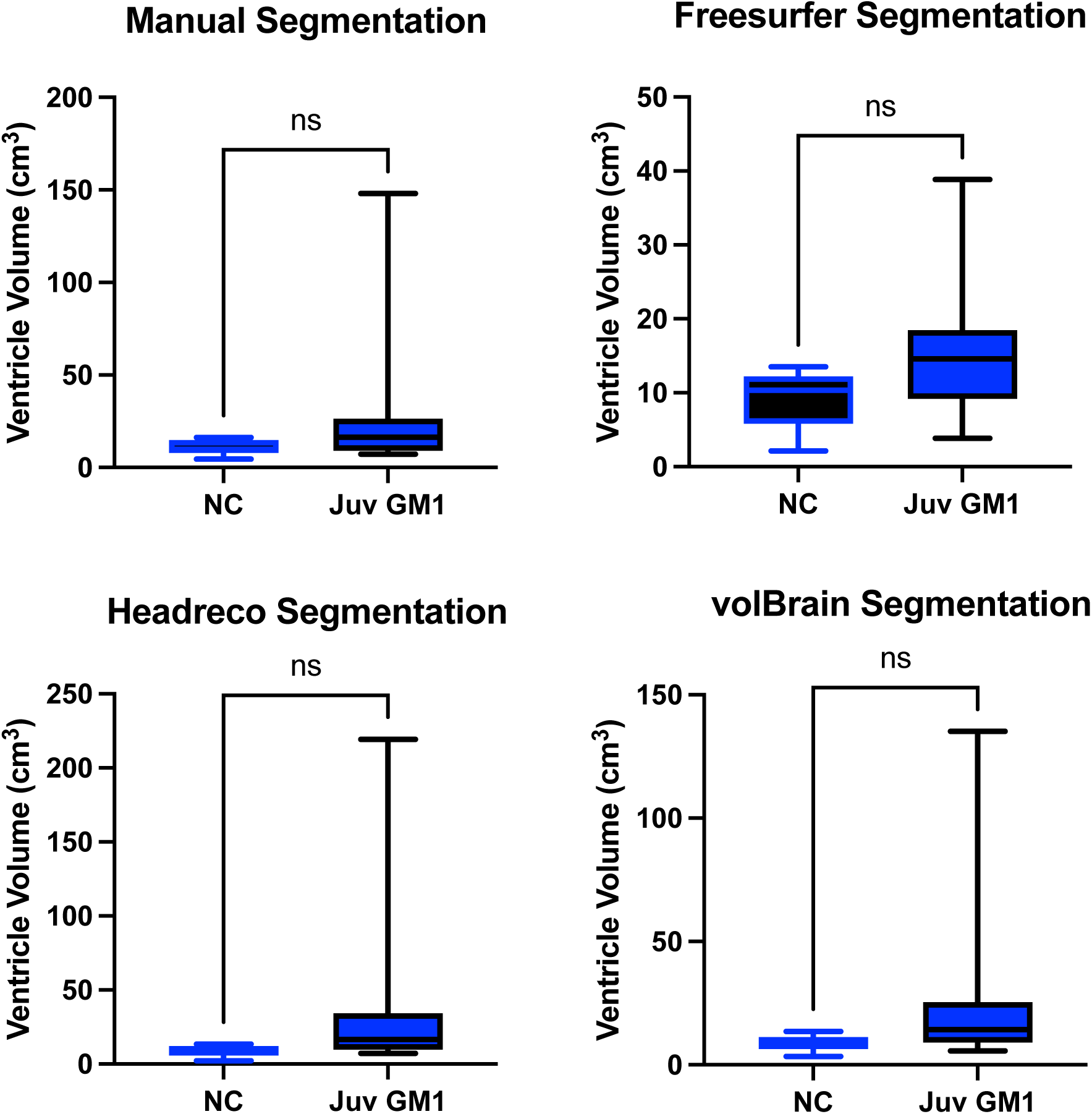
Cross-sectional evaluation of the 3 automated segmentation algorithms to demonstrate cohort differences in ventricle volume. Juvenile (Juv) GM1 patients are shown in blue. Neurotypical controls (NC) are shown in black. *P*-values were calculated from the *t*-statistic. * *P* < 0.05, ** *P* < 0.01, *** *P* < 0.001, **** *P* < 0.0001.

**Figure F2.**
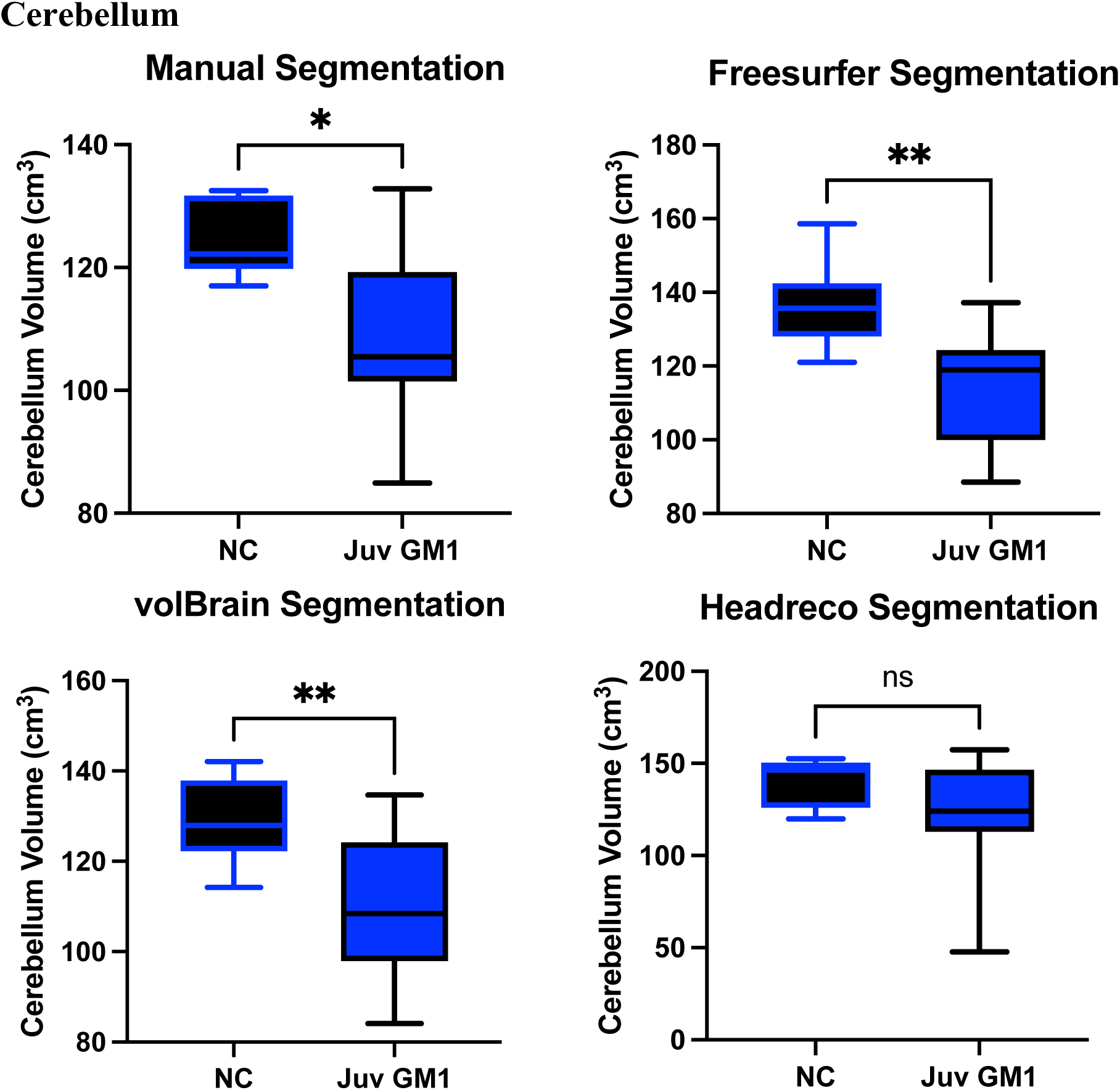
Cross-sectional evaluation of the 3 automated segmentation algorithms to demonstrate cohort differences in cerebellum volume. Juvenile (Juv) GM1 patients are shown in blue. Neurotypical controls (NC) are shown in black. *P*-values were calculated from the *t*-statistic. * *P* < 0.05, ** *P* < 0.01, *** *P* < 0.001, **** *P* < 0.0001.

**Figure F3.**
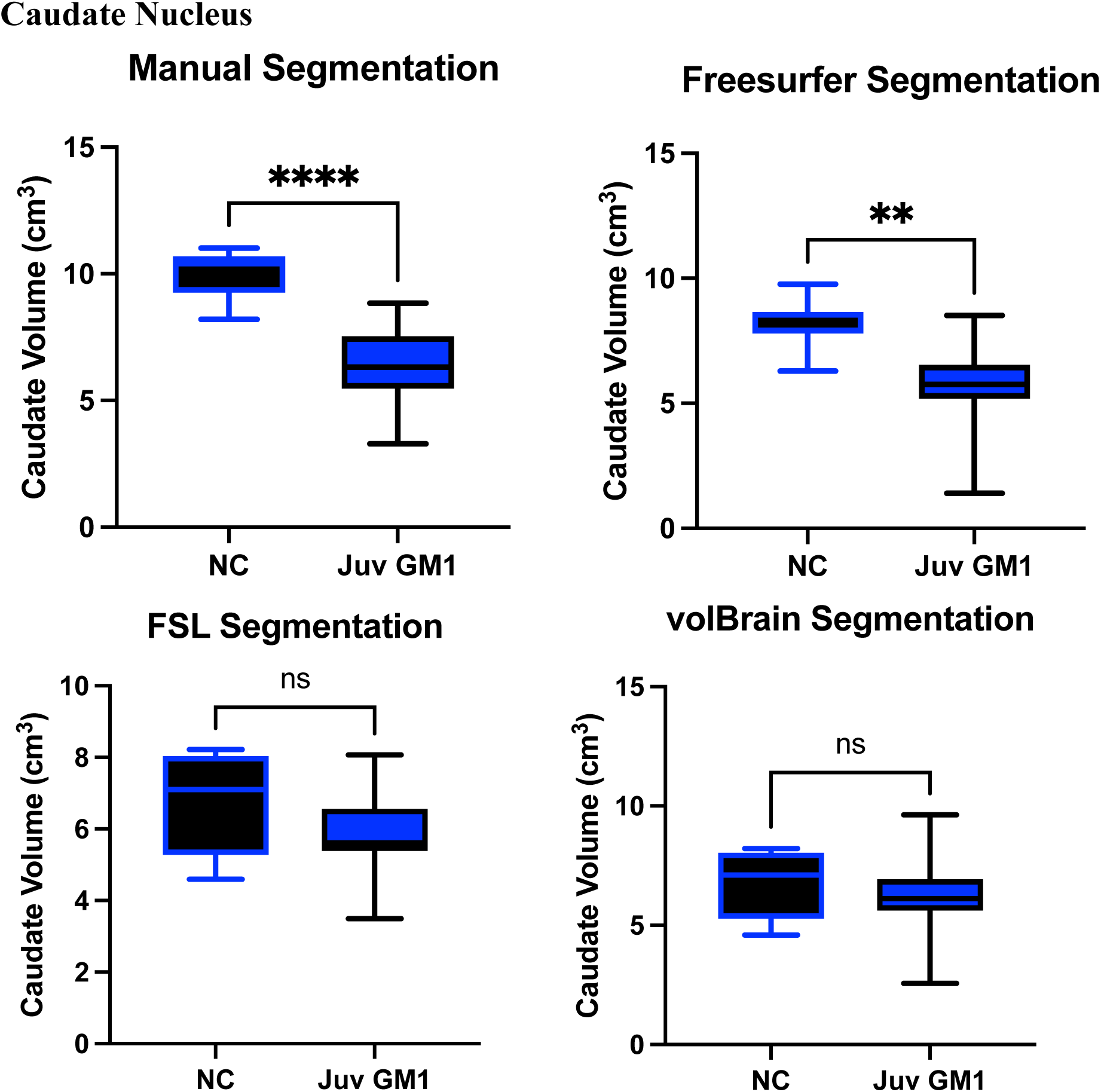
Cross-sectional evaluation of the 3 automated segmentation algorithms to demonstrate cohort differences in caudate volume. Juvenile (Juv) GM1 patients are shown in blue. Neurotypical controls (NC) are shown in black. *P*-values were calculated from the *t*-statistic. * *P* < 0.05, ** *P* < 0.01, *** *P* < 0.001, **** *P* < 0.0001.

**Figure F4.**
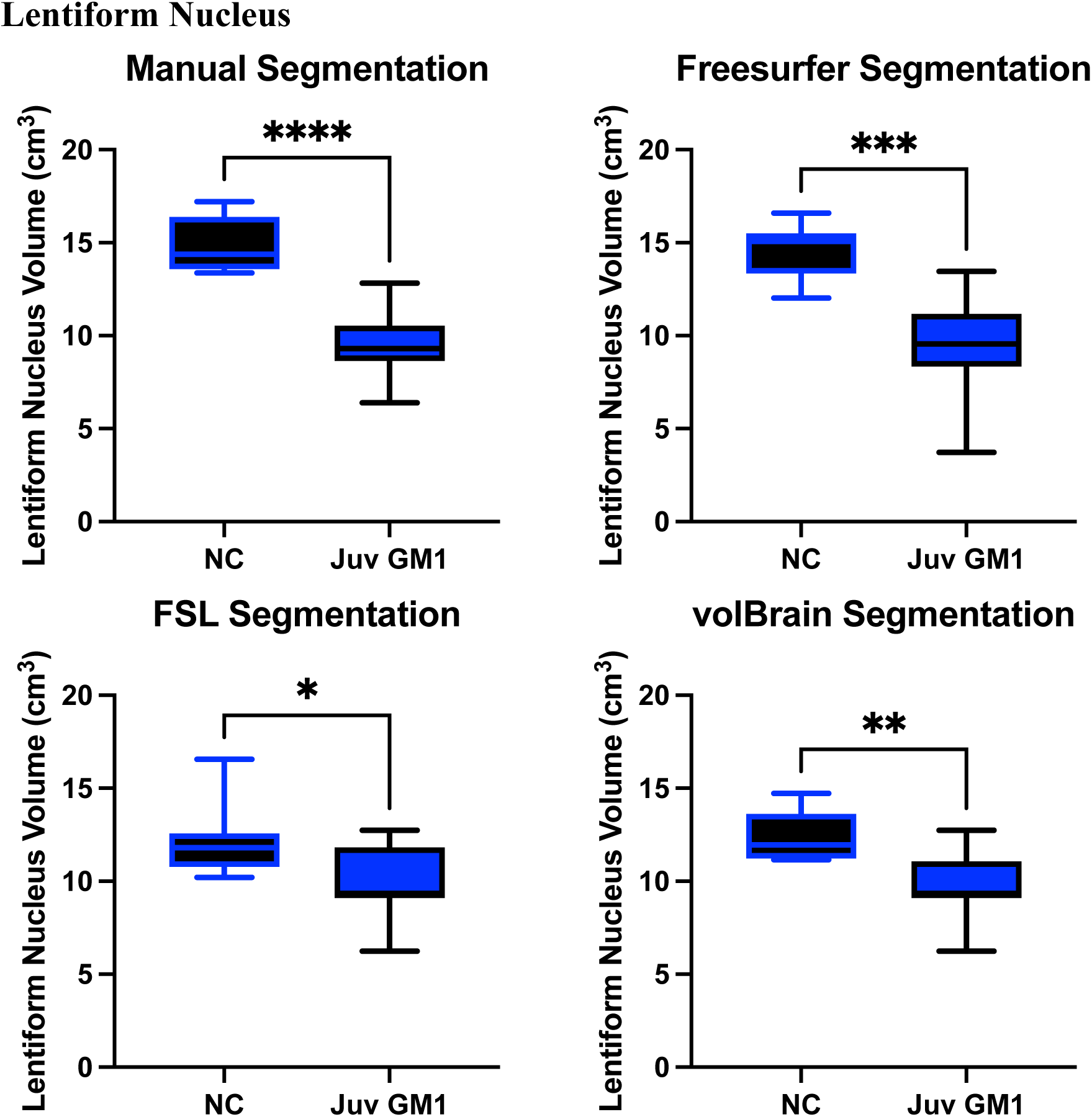
Cross-sectional evaluation of the 3 automated segmentation algorithms to demonstrate cohort differences in lentiform nucleus volume. Juvenile (Juv) GM1 patients are shown in blue. Neurotypical controls (NC) are shown in black. *P*-values were calculated from the *t*-statistic. * *P* < 0.05, ** *P* < 0.01, *** *P* < 0.001, **** *P* < 0.0001.

**Figure F5.**
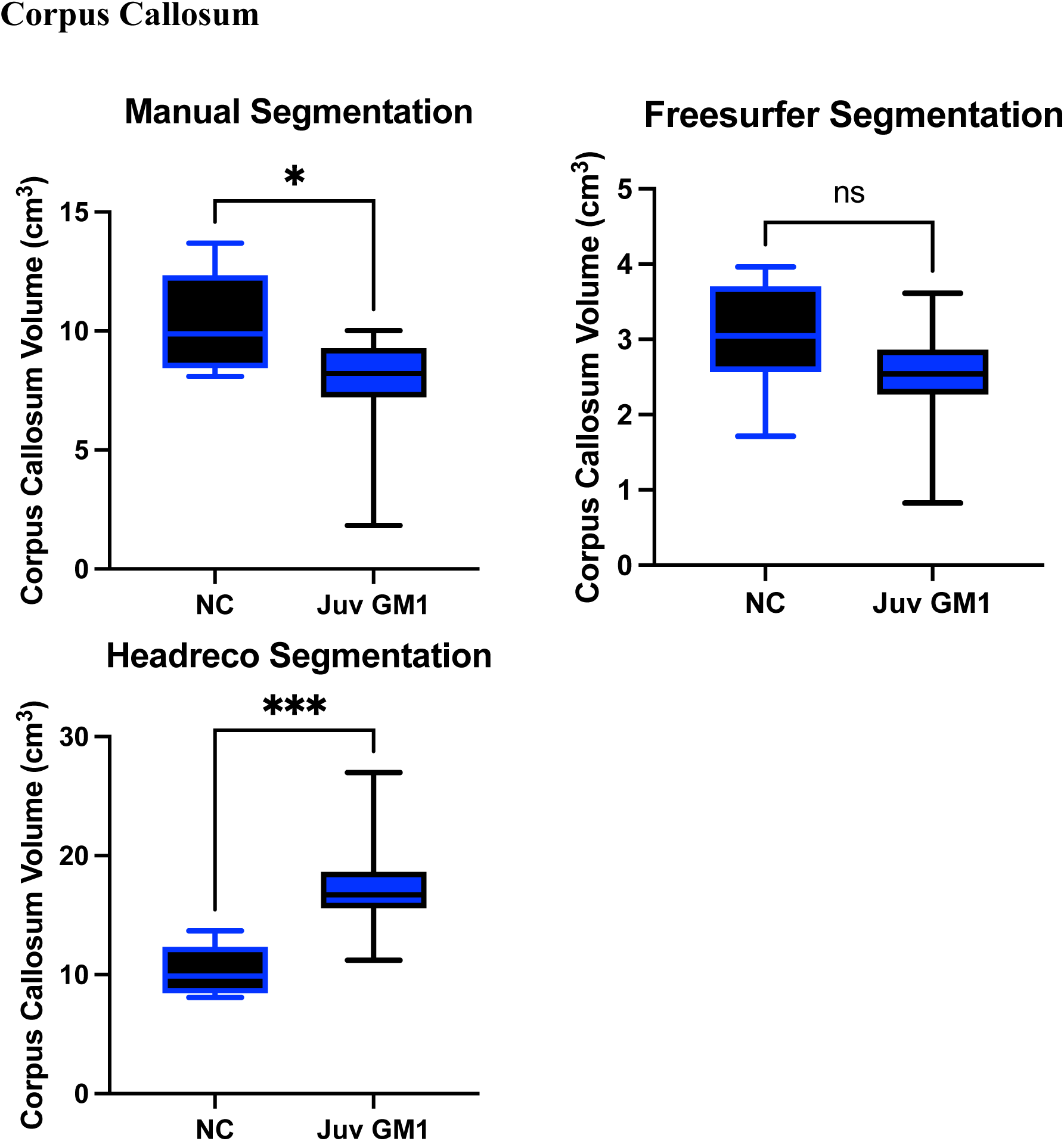
Cross-sectional evaluation of the 2 automated segmentation algorithms to demonstrate cohort differences in corpus callosum volume. Juvenile (Juv) GM1 patients are shown in blue. Neurotypical controls (NC) are shown in black. *P*-values were calculated from the *t*-statistic. * *P* < 0.05, ** *P* < 0.01, *** *P* < 0.001, **** *P* < 0.0001.

### Supplement G: Correlation Strength Tables between Manual and Automated Segmentation

**Table GI.** Comparison of correlation coefficients (R^2^, top row) between the 5 fully automated segmentation pipelines and the manual pipeline in the 7 different brain regions for the <u>neurotypical controls</u>. Normality was determined using the Shapiro-Wilk test. *P-*values (bottom row) were calculated from the Pearson product moment correlation coefficient when the data was normally distributed and from Spearman’s rank correlation coefficient when the data was not normally distributed and designated with a ^+^. N/A are designations where the region was not calculated using the specified segmentation algorithm.

| **Structure** | **Freesurfer** | **FSL** | **volBrain** | **SPM** | **SimNIBS**<br>**Headreco**<br>**(SPM+CAT)** |
| --- | --- | --- | --- | --- | --- |
| **Whole Brain** | 0.8256<br>$p < 0.0001$ | 0.9058<br>$p < 0.0001$ | 0.9852<br>$p < 0.0001$ | 0.8629<br>$p < 0.0001^+$ | 0.8593<br>$p < 0.0001$ |
| **Ventricles** | 0.6180<br>$p < 0.0001$ | N/A | 0.9452<br>$p < 0.0001$ | N/A | 0.7951<br>$p < 0.0001$ |
| **Cerebellum** | 0.6910<br>$p < 0.0001$ | N/A | 0.8269<br>$p < 0.0001$ | N/A | 0.5384<br>$p = 0.0003$ |
| **Thalamus** | 0.5359<br>$p = 0.0004$ | 0.5074<br>$p = 0.0006$ | 0.6276<br>$p < 0.0001$ | N/A | 0.5322<br>$p < 0.0001^+$ |
| **Caudate** | 0.8174<br>$p < 0.0001$ | 0.5416<br>$p = 0.0003$ | 0.7282<br>$p < 0.0001$ | N/A | N/A |
| **Lentiform**<br>**Nucleus** | 0.7251<br>$p < 0.0001$ | 0.5271<br>$p = 0.0004^+$ | 0.5562<br>$p = 0.0002$ | N/A | N/A |
| **Corpus**<br>**Callosum** | 0.06547<br>$p = 0.2904$ | N/A | N/A | N/A | 0.4829<br>$p = 0.0010$ |

**Table GII.** Comparison of correlation coefficients (R^2^, top row) between the 5 fully automated segmentation pipelines and the manual pipeline in the 7 different brain regions for the <u>juvenile</u> <u>GM1 patients</u>. Normality was determined using the Shapiro-Wilk test. *P-*values (bottom row) were calculated from the Pearson product moment correlation coefficient when the data was normally distributed and from Spearman’s rank correlation coefficient when the data was not normally distributed and designated with a ^+^. N/A are designations where the region was not calculated using the specified segmentation algorithm.

| **Structure** | **Freesurfer** | **FSL** | **volBrain** | **SPM** | **SimNIBS**<br>**Headreco**<br>**(SPM+CAT)** |
| --- | --- | --- | --- | --- | --- |
| **Whole Brain** | 0.9196<br>$p < 0.0001^+$ | 0.6038<br>$p < 0.0001$ | 0.9583<br>$p < 0.0001^+$ | 0.9642<br>$p < 0.0001^+$ | 0.8956<br>$p < 0.0001^+$ |
| **Ventricles** | 0.9765<br>$p < 0.0001^+$ | N/A | 0.9782<br>$p < 0.0001^+$ | N/A | 0.7422<br>$p < 0.0001^+$ |
| **Cerebellum** | 0.9133<br>$p < 0.0001$ | N/A | 0.9258<br>$p < 0.0001$ | N/A | 0.6009<br>$p < 0.0001^+$ |
| **Thalamus** | 0.4359<br>$p < 0.0001$ | 0.5033<br>$p < 0.0001^+$ | 0.8053<br>$p < 0.0001^+$ | N/A | 0.7073<br>$p < 0.0001^+$ |
| **Caudate** | 0.8181<br>$p < 0.0001^+$ | 0.5068<br>$p < 0.0001^+$ | 0.8885<br>$p < 0.0001^+$ | N/A | N/A |
| **Lentiform**<br>**Nucleus** | 0.5660<br>$p < 0.0001$ | 0.4266<br>$p < 0.0001$ | 0.6605<br>$p < 0.0001$ | N/A | N/A |
| **Corpus**<br>**Callosum** | 0.4050<br>$p = 0.0122^+$ | N/A | N/A | N/A | 0.3131<br>$p < 0.0001$ |

**Table GIII.** Comparison of correlation coefficients (R^2^, top row) between the 5 fully automated segmentation pipelines and the manual pipeline in the 7 different brain regions for the <u>late-</u> <u>infantile GM1 patients</u>. Normality was determined using the Shapiro-Wilk test. *P-*values (bottom row) were calculated from the Pearson product moment correlation coefficient when the data was normally distributed and from Spearman’s rank correlation coefficient when the data was not normally distributed and designated with a ^+^. N/A are designations where the region was not calculated using the specified segmentation algorithm.

| Structure | Freesurfer | FSL | volBrain | SPM | SimNIBS<br>Headreco<br>(SPM+CAT) |
| --- | --- | --- | --- | --- | --- |
| **Whole Brain** | 0.3460<br>$p = 0.0736$ | 0.6585<br>$p = 0.0044$ | 0.9169<br>$p = 0.0072^+$ | 0.5458<br>$p = 0.0094$ | 0.8246<br>$p = 0.0003$ |
| **Ventricles** | 0.7467<br>$p = 0.0013$ | N/A | 0.9974<br>$p < 0.0001$ | N/A | 0.9950<br>$p < 0.0001$ |
| **Cerebellum** | 0.6728<br>$p = 0.0037$ | N/A | 0.8374<br>$p < 0.0001$ | N/A | 0.8779<br>$p < 0.0001$ |
| **Thalamus** | 0.5347<br>$p = 0.0163$ | 0.6585<br>$p = 0.0044$ | 0.8632<br>$p < 0.0001^+$ | N/A | 0.7809<br>$p = 0.0003$ |
| **Caudate** | 0.8536<br>$p = 0.0001$ | 0.2584<br>$p = 0.1335$ | 0.7394<br>$p = 0.0027^+$ | N/A | N/A |
| **Lentiform**<br>**Nucleus** | 0.6619<br>$p = 0.0347^+$ | 0.7713<br>$p = 0.0008$ | 0.6518<br>$p = 0.0368^+$ | N/A | N/A |
| **Corpus**<br>**Callosum** | 0.06030<br>$p = 0.4941$ | N/A | N/A | N/A | $< 0.01$<br>$p = 0.9895^+$ |

### Supplement H: Correlations between Manual and Freesurfer Segmentation

**Figure H1.**
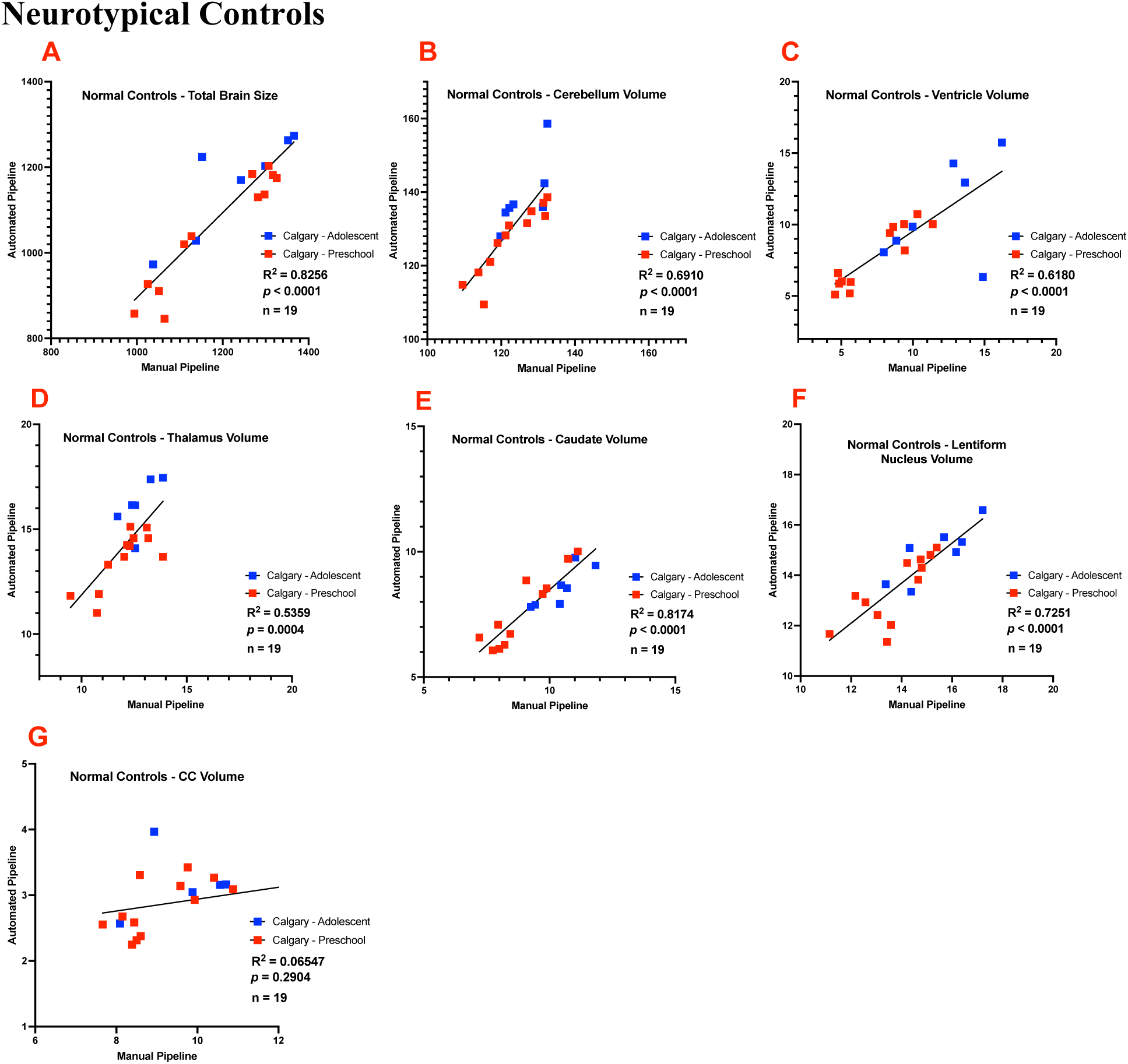
Correlations between Freesurfer’s automated and the manual approach in neurotypical controls for the A.) Total brain volume B.) Cerebellum volume C.) Ventricle volume D.) Thalamic volume E.) Caduate volume F.) Lentiform nucleus volume and G.) Corpus callosum volume. Participants from the Calgary preschool MRI data set are shown in red and participants from the adolescent Calgary dataset are shown in blue. All volumes are shown in cm^3^.

**Figure H2.**
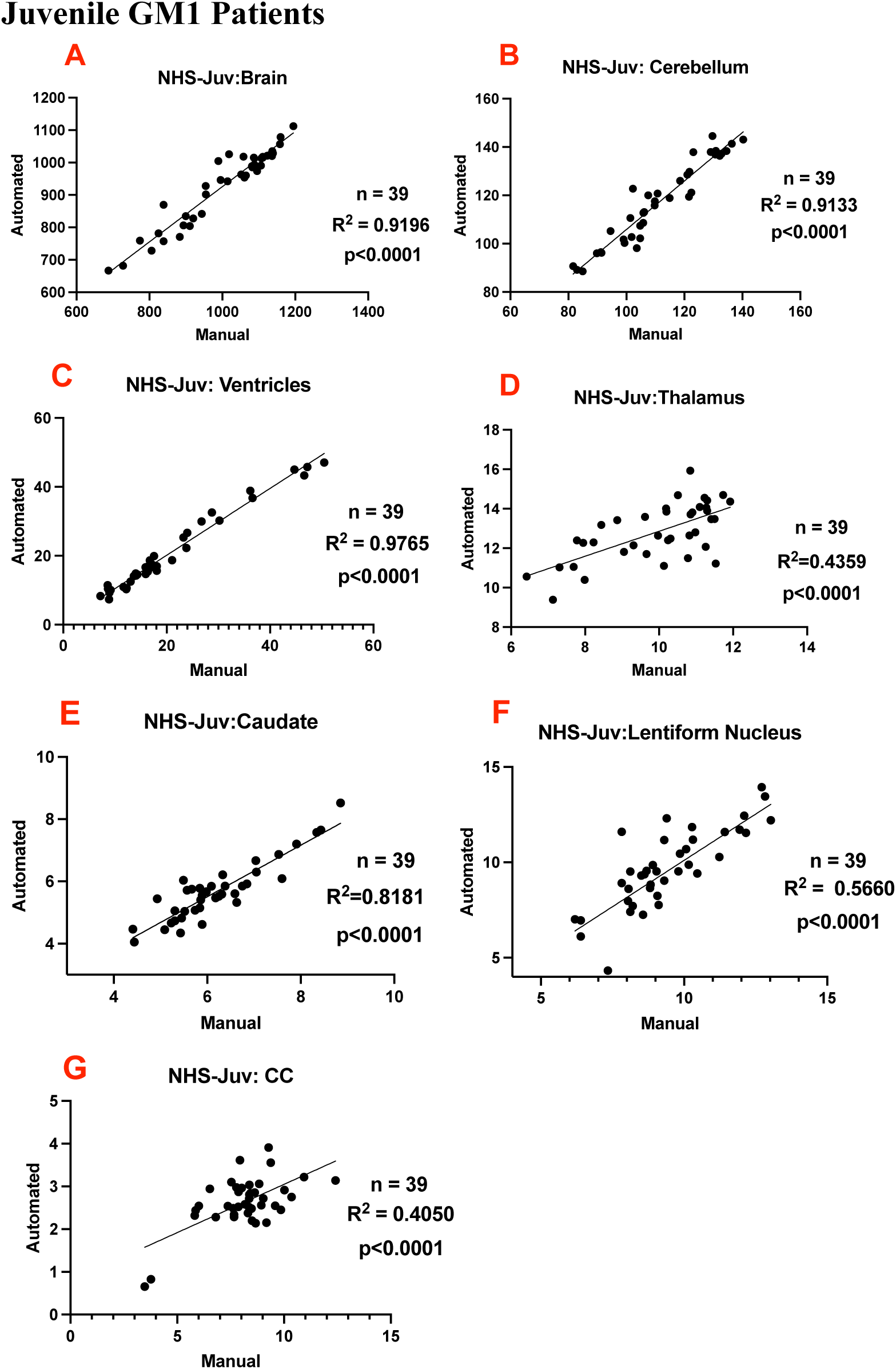
Correlations between Freesurfer’s automated and the manual approach in natural history study (NHS) juvenile GM1 gangliosidosis patients for the A.) Total brain volume B.) Cerebellum volume C.) Ventricle volume D.) Thalamic volume E.) Caduate volume F.) Lentiform nucleus volume and G.) Corpus callosum volume. All volumes are shown in cm^3^.

**Figure H3.**
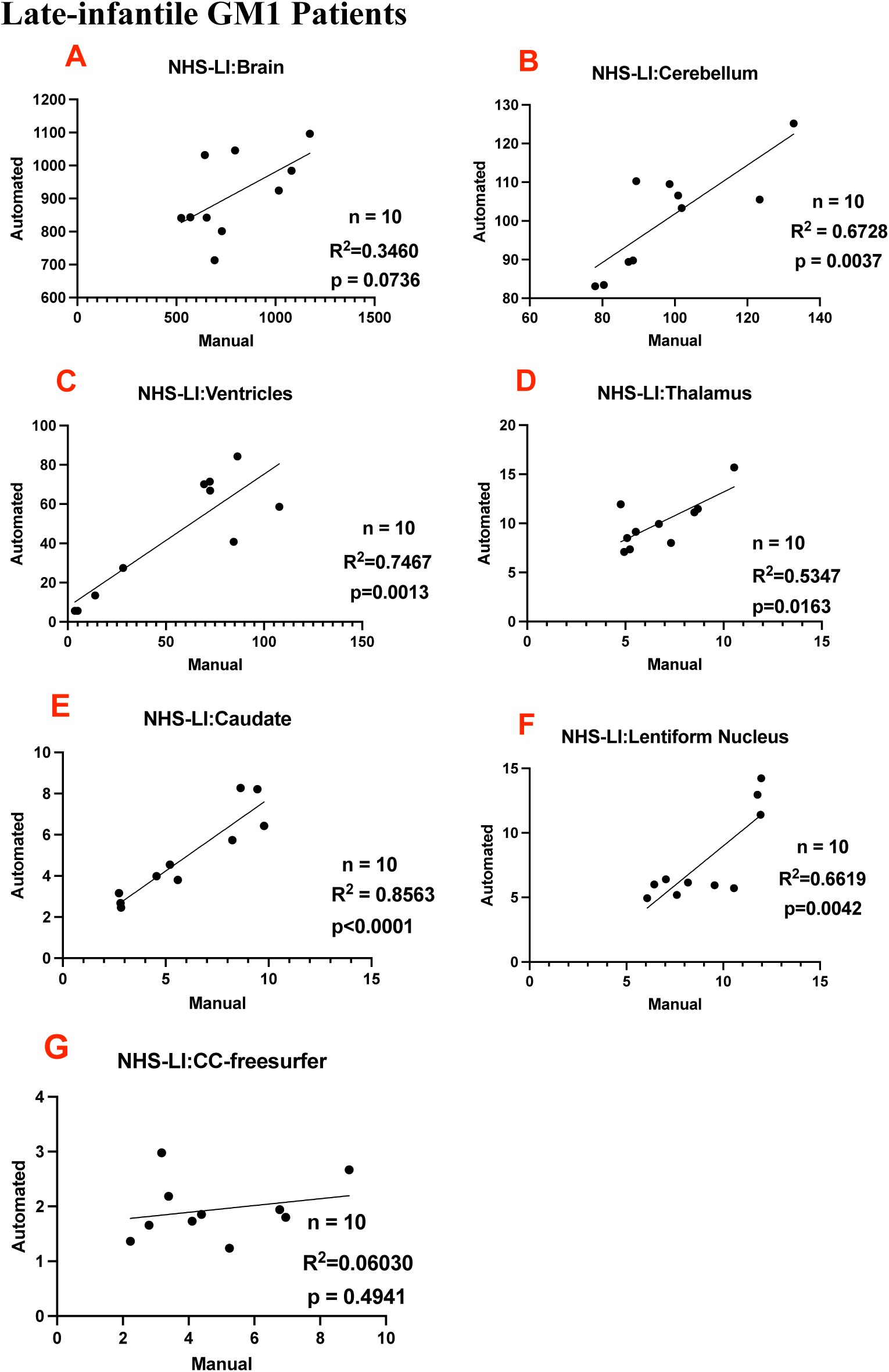
Correlations between Freesurfer’s automated and the manual approach in natural history study (NHS) late-infantile GM1 gangliosidosis patients for the A.) Total brain volume B.) Cerebellum volume C.) Ventricle volume D.) Thalamic volume E.) Caduate volume F.) Lentiform nucleus volume and G.) Corpus callosum volume. All volumes are shown in cm^3^.

### Supplement I: Correlations between Manual and volBrain Segmentation

**Figure I1.**
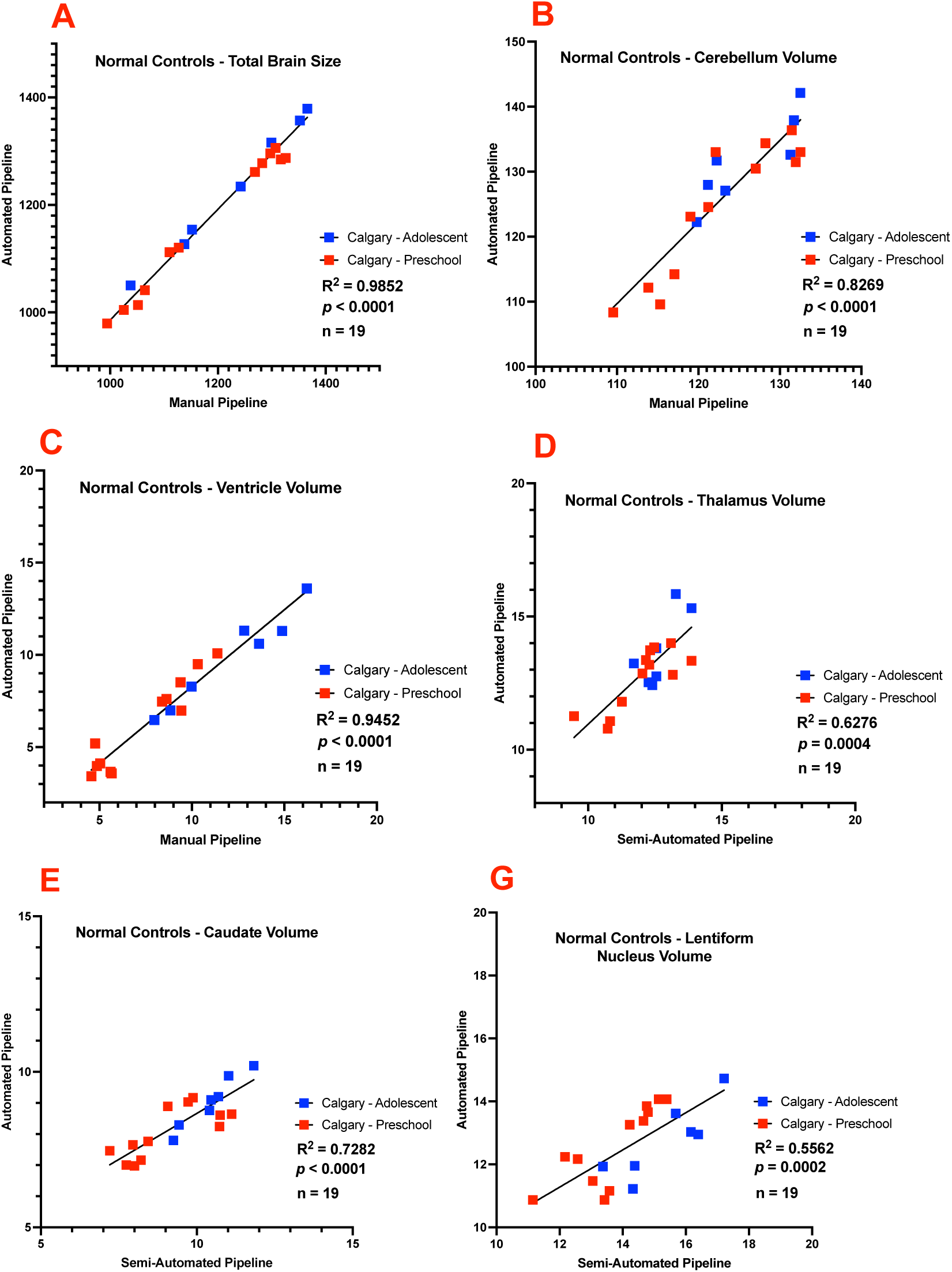
Correlations between volBrain’s automated and the manual approach in neurotypical controls for the A.) Total brain volume B.) Cerebellum volume C.) Ventricle volume D.) Thalamic volume E.) Caduate volume and F.) Lentiform nucleus volume. Participants from the Calgary preschool MRI data set are shown in red and participants from the adolescent Calgary dataset are shown in blue. All volumes are shown in cm^3^.

**Figure I2.**
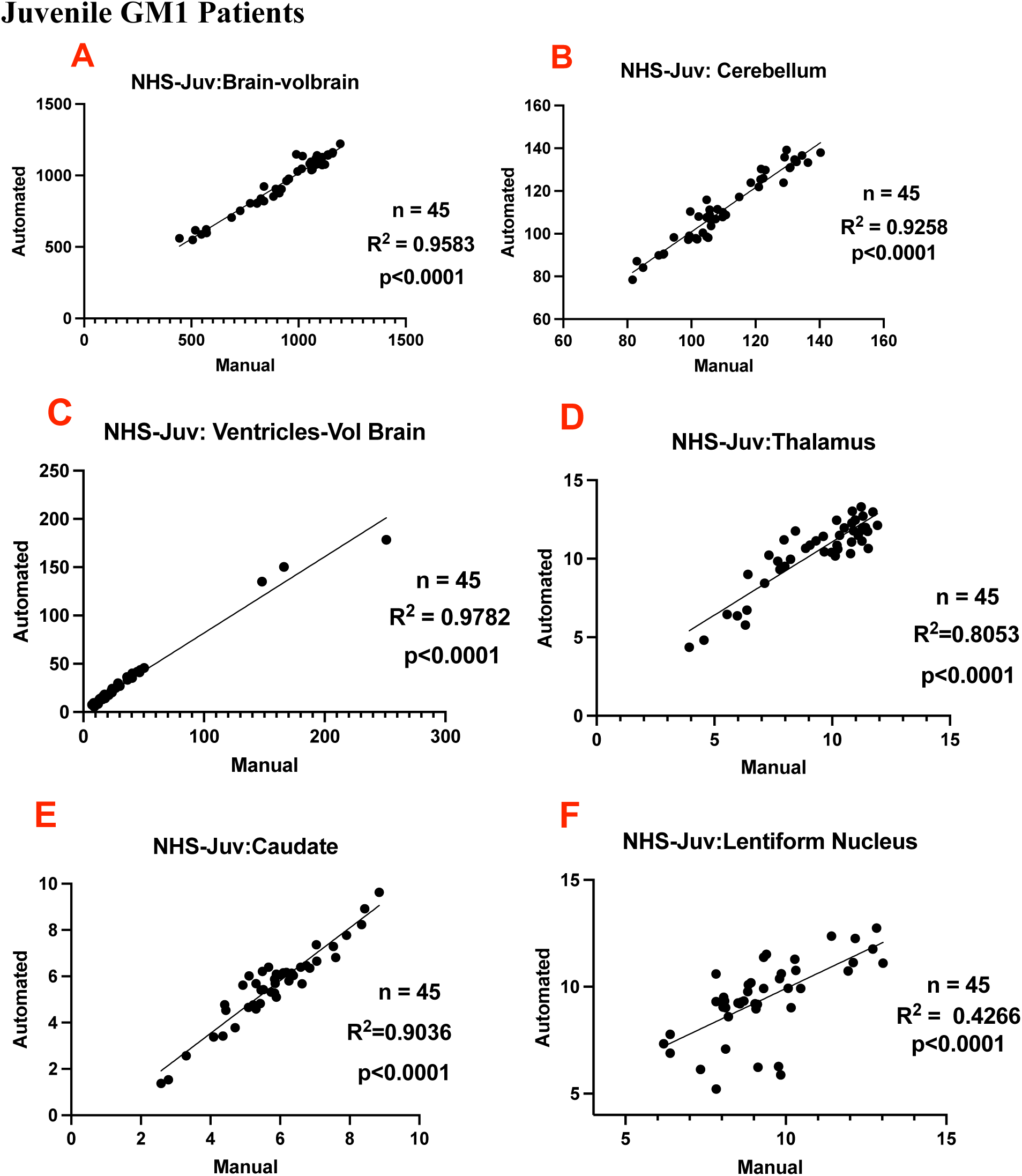
Correlations between volBrain’s automated and the manual approach in natural history study (NHS) juvenile GM1 gangliosidosis patients for the A.) Total brain volume B.) Cerebellum volume C.) Ventricle volume D.) Thalamic volume E.) Caduate volume and F.) Lentiform nucleus volume. All volumes are shown in cm^3^.

**Figure I3.**
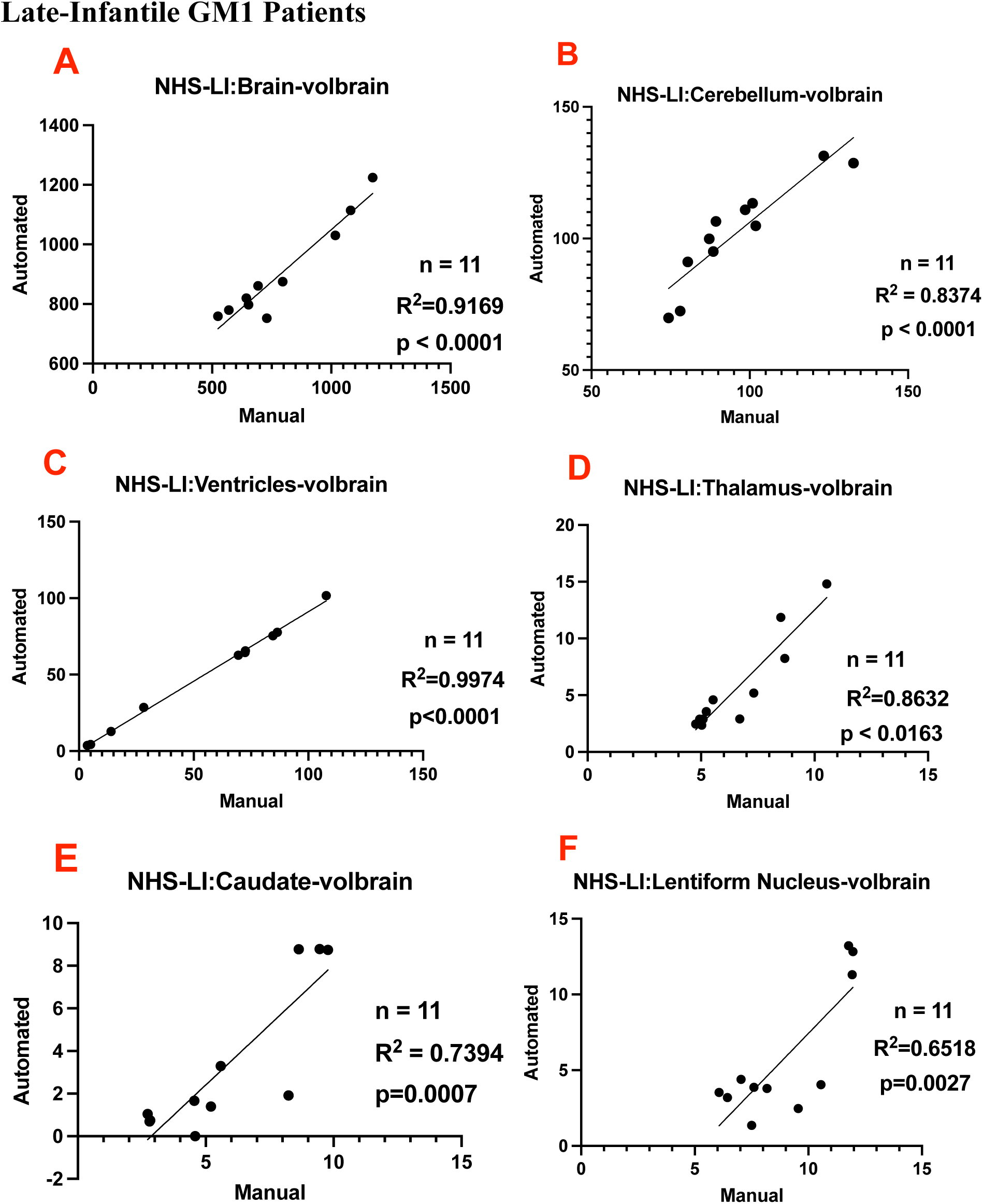
Correlations between volBrain’s automated and the manual approach in natural history study (NHS) late-infantile GM1 gangliosidosis patients for the A.) Total brain volume B.) Cerebellum volume C.) Ventricle volume D.) Thalamic volume E.) Caduate volume and F.) Lentiform nucleus volume. All volumes are shown in cm^3^.

### Supplement J: Correlations between Manual and FSL Segmentation

**Figure J1.**
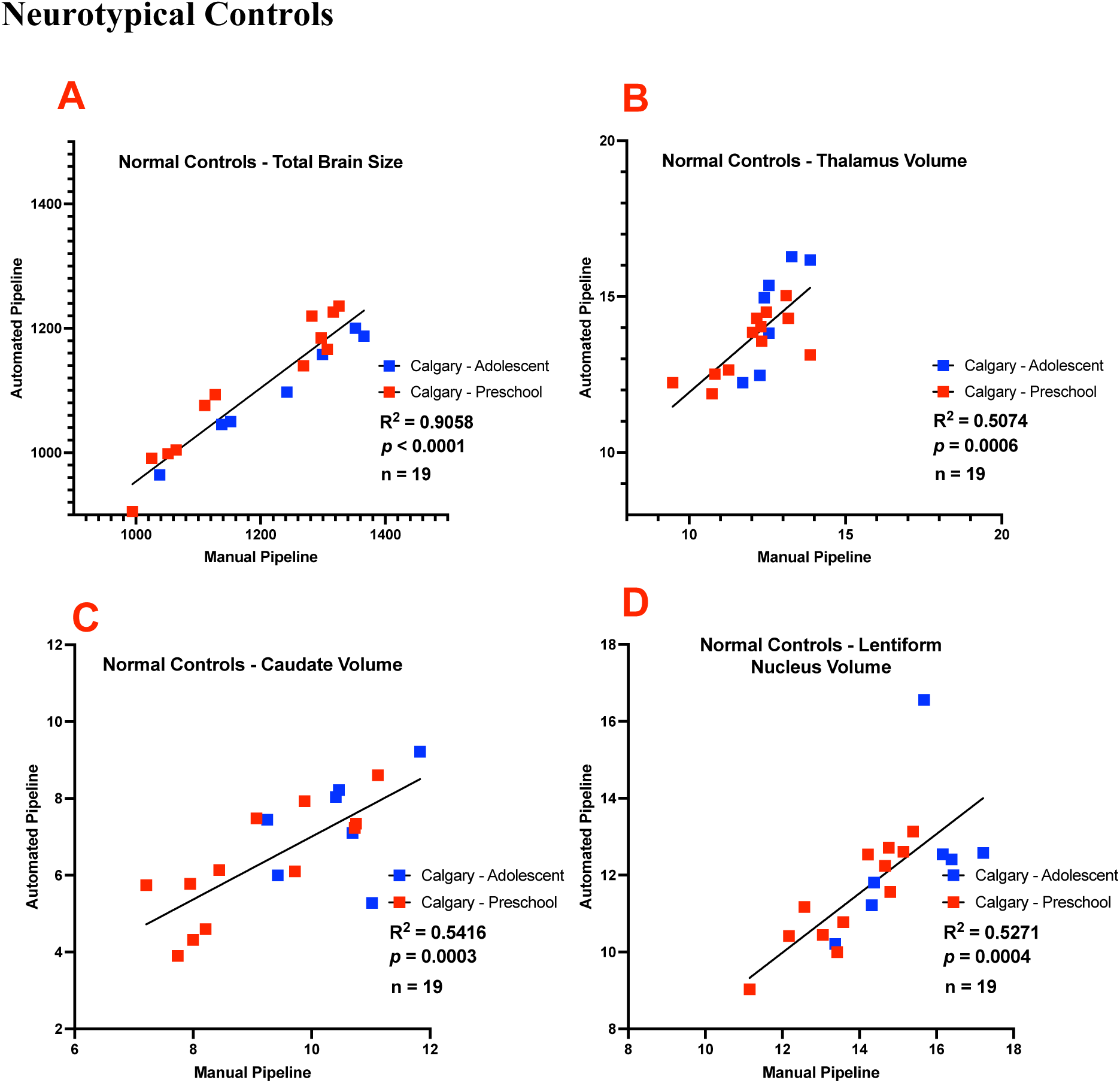
Correlations between FSL’s automated and the manual approach in neurotypical controls for the A.) Total brain volume B.) Thalamic volume C.) Caduate volume and D.) Lentiform nucleus volume. Participants from the Calgary preschool MRI data set are shown in red and participants from the adolescent Calgary dataset are shown in blue. All volumes are shown in cm^3^.

**Figure J2.**
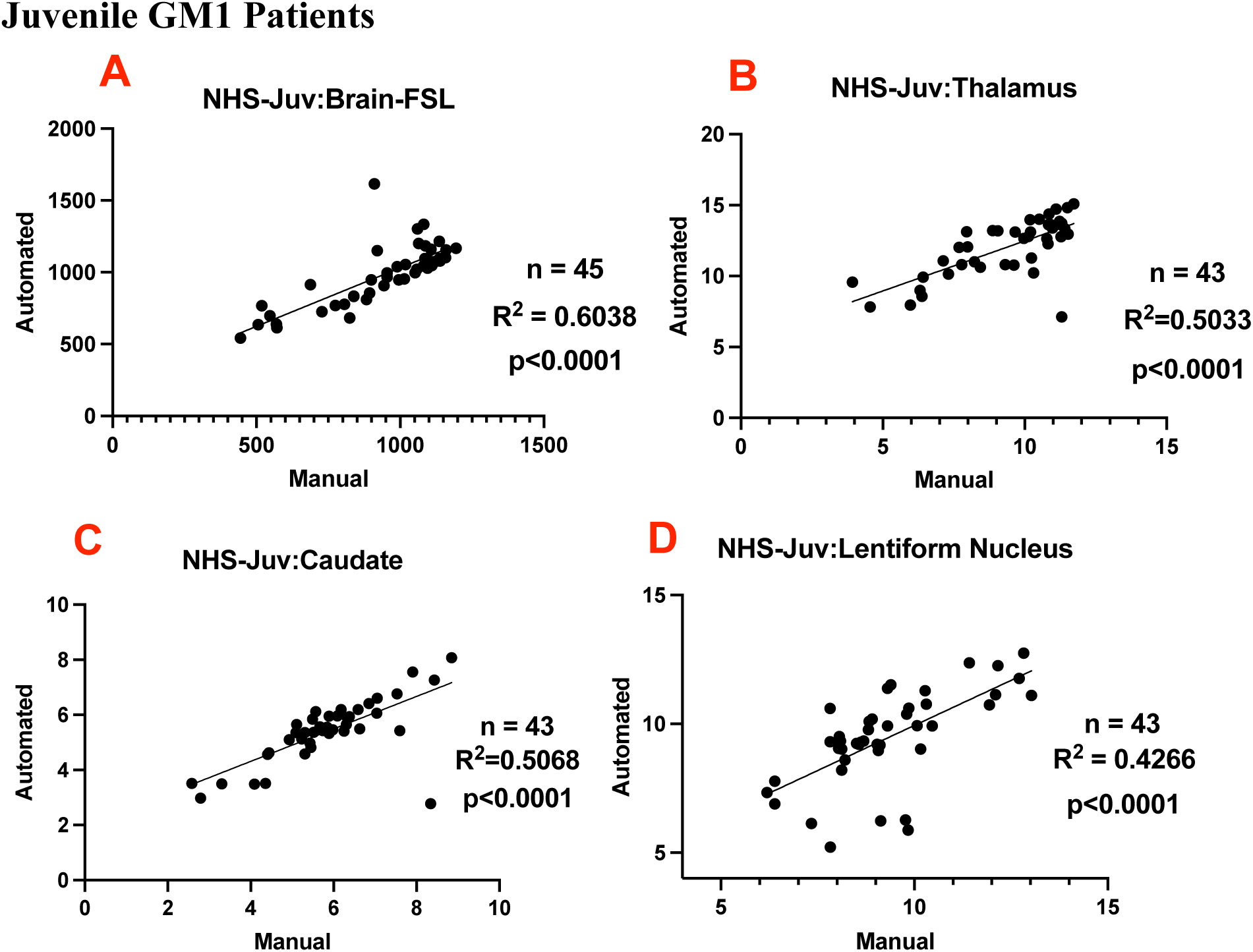
Correlations between FSL’s automated and the manual approach in natural history study (NHS) juvenile GM1 gangliosidosis patients for the A.) Total brain volume B.) Thalamic volume C.) Caduate volume and D.) Lentiform nucleus volume. All volumes are shown in cm^3^.

**Figure J3.**
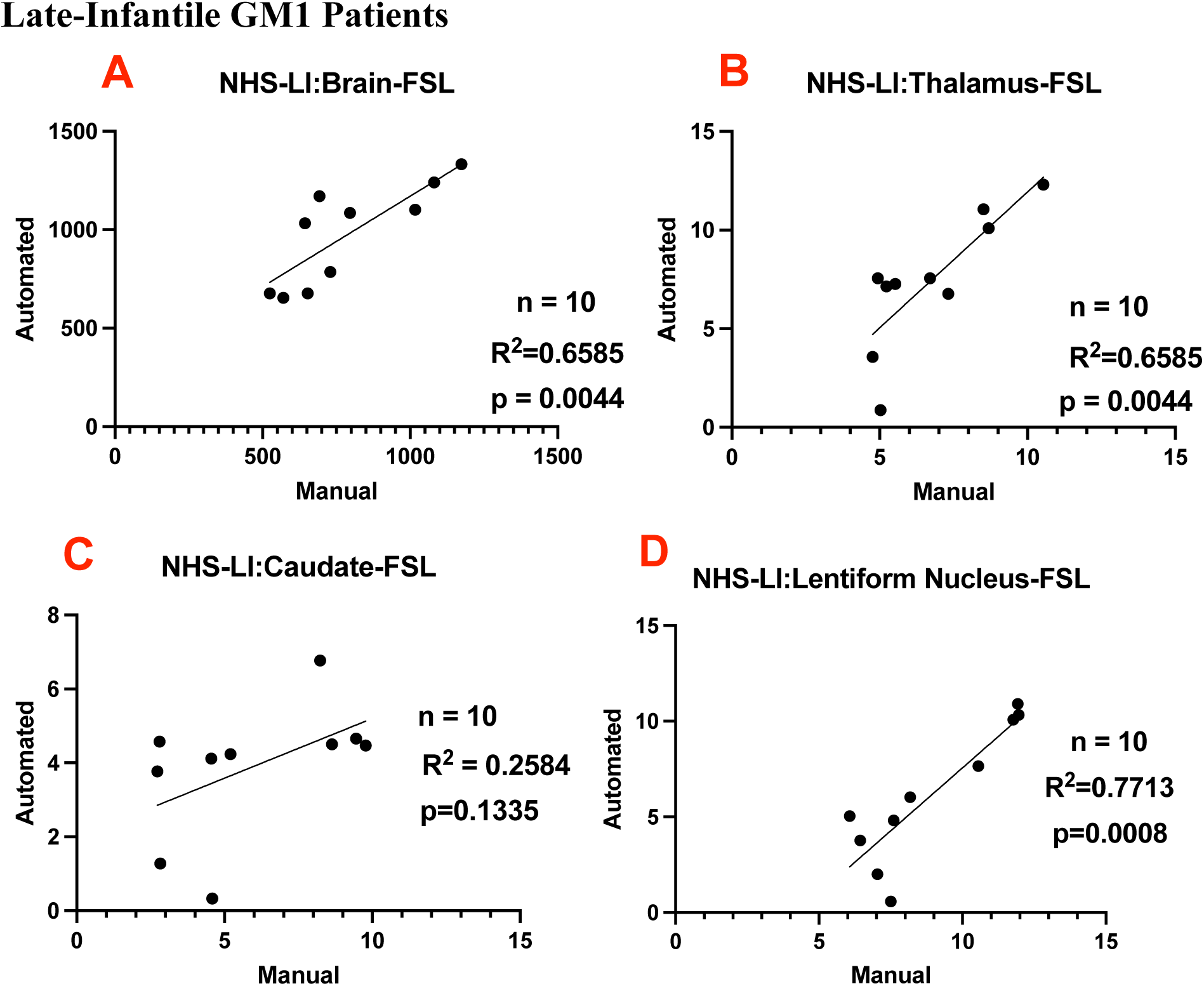
Correlations between FSL’s automated and the manual approach in natural history study (NHS) juvenile GM1 gangliosidosis patients for the A.) Total brain volume B.) Thalamic volume C.) Caduate volume and D.) Lentiform nucleus volume. All volumes are shown in cm^3^.

### Supplement K: Correlations between Manual and SPM Segmentation

**Figure K1.**
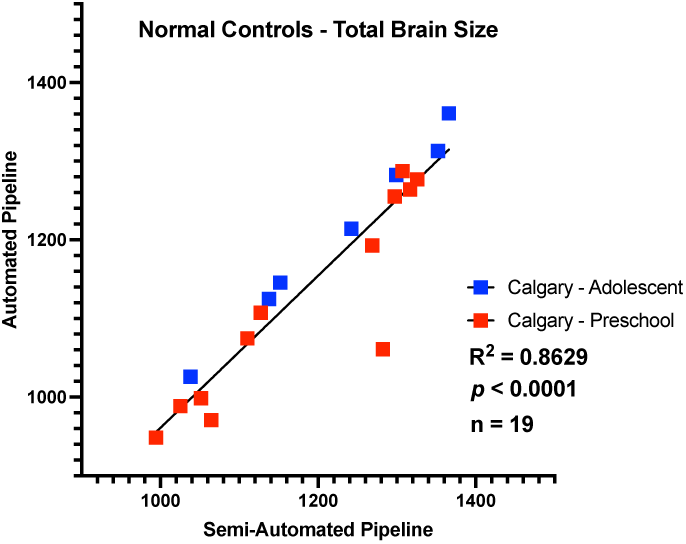
Correlations between SPM’s automated and the manual approach in neurotypical controls for the total brain volume. Participants from the Calgary preschool MRI data set are shown in red and participants from the adolescent Calgary dataset are shown in blue. All volumes are shown in cm^3^.

**Figure K2.**
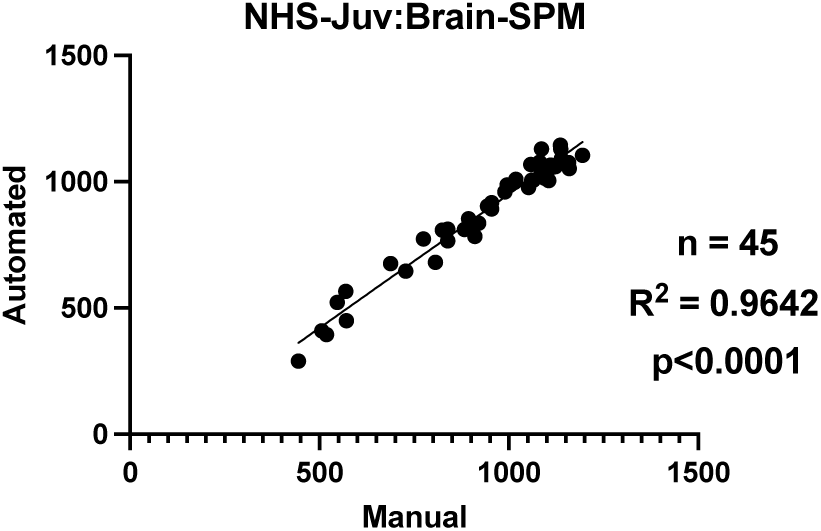
Correlations between SPM’s automated and the manual approach in natural history study (NHS) juvenile GM1 gangliosidosis patients for total brain volume. All volumes are shown in cm^3^.

**Figure K3.**
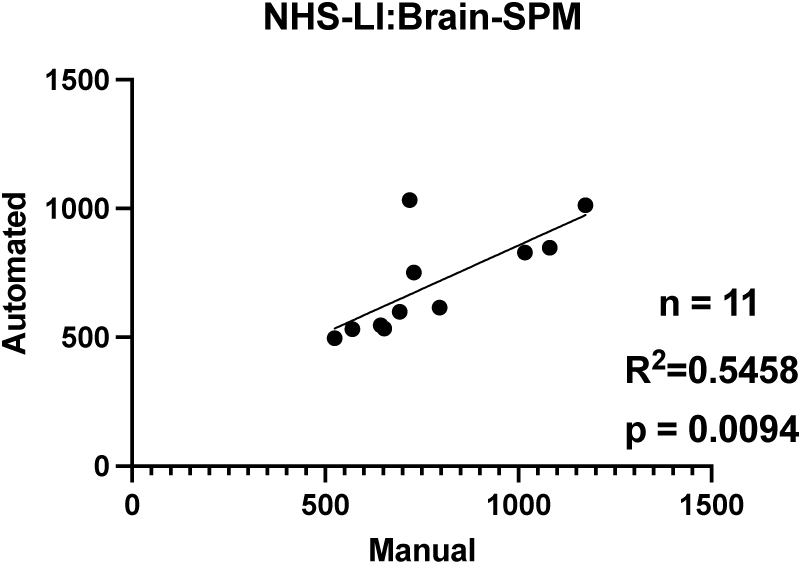
Correlations between SPM’s automated and the manual approach in natural history study (NHS) late-infantile GM1 gangliosidosis patients for total brain volume. All volumes are shown in cm^3^.

### Supplement L: Correlations between Manual and Headreco Segmentation

**Figure L1.**
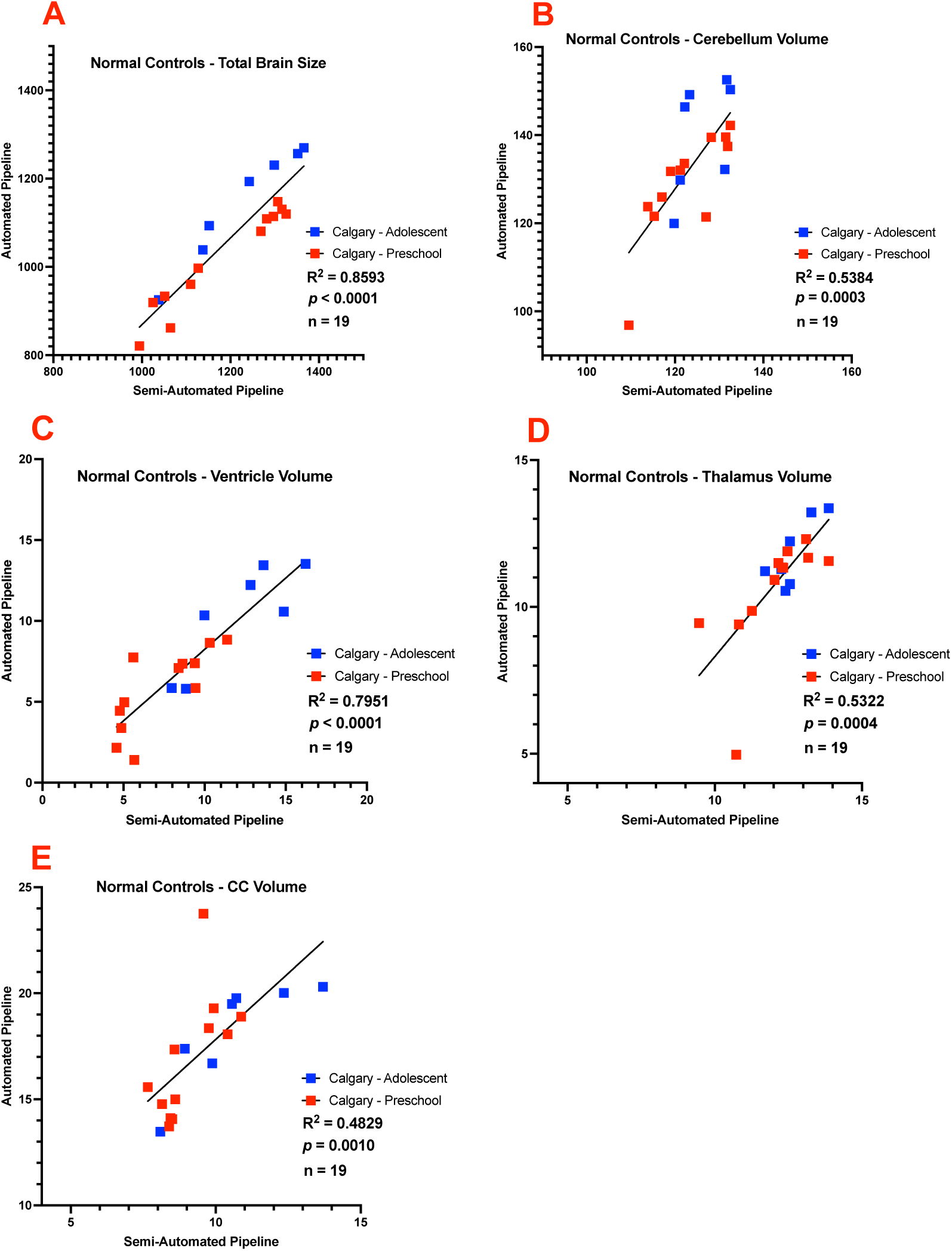
Correlations between Headreco’s automated and the manual approach in neurotypical controls for the A.) Total brain volume B.) Cerebellum volume C.) Ventricle volume D.) Thalamic volume and E.) Corpus callosum volume. Participants from the Calgary preschool MRI data set are shown in red and participants from the adolescent Calgary dataset are shown in blue. All volumes are shown in cm^3^.

**Figure L2.**
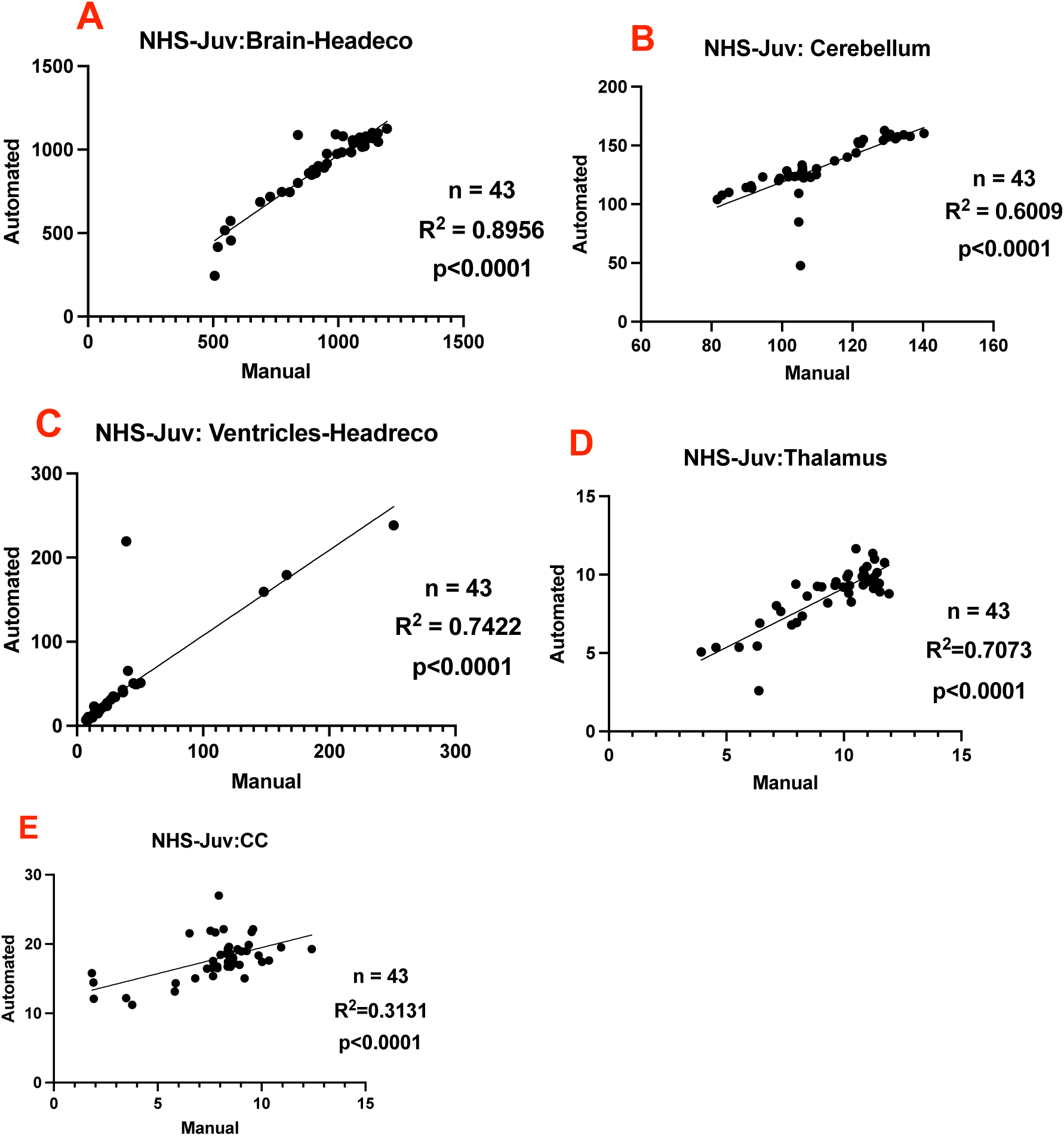
Correlations between Headreco’s automated and the manual approach in natural history study (NHS) juvenile GM1 gangliosidosis patietns for the A.) Total brain volume B.) Cerebellum volume C.) Ventricle volume D.) Thalamic volume and E.) Corpus callosum volume. All volumes are shown in cm^3^.

**Figure L3.**
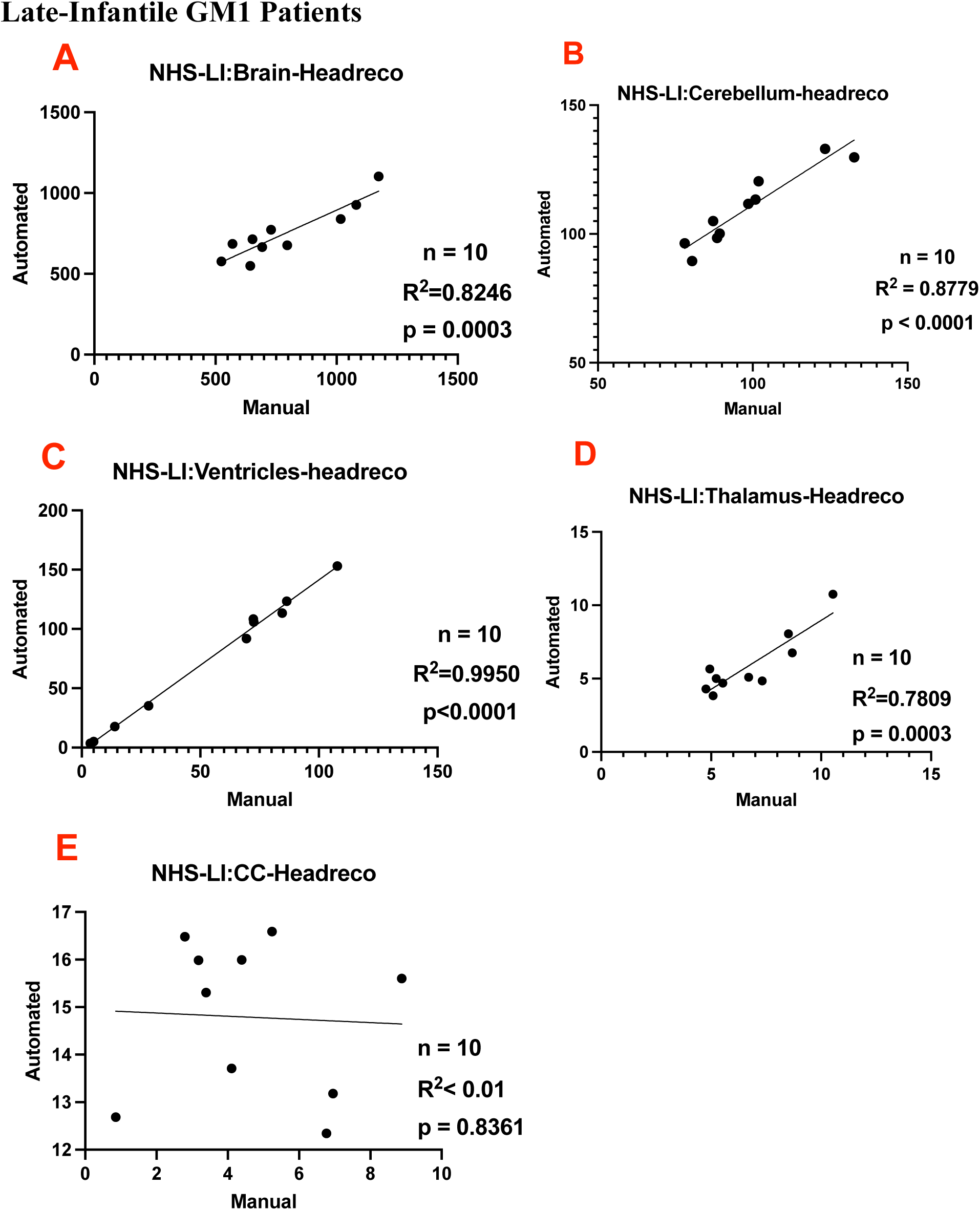
Correlations between Headreco’s automated and the manual approach in natural history study (NHS) late-infantile GM1 gangliosidosis patietns for the A.) Total brain volume B.) Cerebellum volume C.) Ventricle volume D.) Thalamic volume and E.) Corpus callosum volume. All volumes are shown in cm^3^.

### Supplement M: Slopes and Intercepts between Manual and Automated Segmentation

**Table MI.** Comparison of slopes and intercepts of the linear regression modeling between the 5 fully automated segmentation pipelines and the manual pipeline in the 7 different brain regions for the neurotypical controls. N/A are designations where the region was not calculated using the specified segmentation algorithm. The slope of the linear regression line is given in the top row, and the intercept is given in the bottom row for each cell. The 95% confidence interval for the slope and intercept shown in the brackets. For all linear regression modeling the automated volumetric calculations were along the y-axis, and the manual volumetric calculations were along the x-axis.

| Structure | Freesurfer | FSL | volBrain | SPM | SimNIBS (SPM+CAT) |
| --- | --- | --- | --- | --- | --- |
| **Whole Brain** | 0.99 [0.76, 1.23]<br>-99.32 [-380.9, 182.3] | 0.75 [0.63, 0.88]<br>202.1 [52.83, 351.5] | 1.03 [0.97, 1.10]<br>-44.99 [-122.8, 32.82] | 0.97 [0.77, 1.16]<br>-6.21 [-243.7, 231.2] | 0.98 [0.78, 1.19]<br>112.6 [-357.4, 132.2] |
| **Ventricles** | 0.68 [0.40, 0.95]<br>2.77 [0.13, 5.41] | N/A | 0.83 [0.73, 0.94]<br>-0.05 [-1.04, 0.95] | N/A | 0.88 [0.65, 1.11]<br>-0.58 [-2.81, 1.64] |
| **Cerebellum** | 1.27 [0.84, 1.71]<br>25.61 [-80.32, 27.75] | N/A | 1.26 [0.96, 1.55]<br>-28.53 [-65.00, 7.93] | N/A | 1.38 [0.73, 2.04]<br>-38.04 [-119.2, 43.11] |
| **Thalamus** | 1.15 [0.60, 1.70]<br>0.34 [-6.39, 7.08] | 0.87 [0.43, 1.30]<br>3.26 [-2.11, 8.62] | 0.94 [0.57, 1.31]<br>1.55 [-3.00, 6.10] | N/A | 1.21 [0.63, 1.78]<br>-3.75 [-10.85, 3.35] |
| **Caudate** | 0.89 [0.67, 1.10]<br>-0.39 [-2.47, 1.68] | 0.82 [0.43, 1.20]<br>-1.18 [-4.90, 2.55] | 0.59 [0.41, 0.78]<br>2.75 [0.96, 4.53] | N/A | N/A |
| **Lentiform Nucleus** | 0.80 [0.54, 1.05]<br>2.55 [-1.06, 6.16] | 0.77 [0.40, 1.15]<br>0.72 [-4.67, 6.12] | 0.59 [0.32, 0.86]<br>2.13 [0.28, 8.08] | N/A | N/A |
| **Corpus Callosum** | 0.09 [-0.08, 0.27]<br>2.04 [0.33, 3.74] | N/A | N/A | N/A | 1.25 [0.59, 1.90]<br>5.38 [-1.05, 11.81] |

**Table MII.** Comparison of slopes and intercepts of the linear regression modeling between the 5 fully automated segmentation pipelines and the manual pipeline in the 7 different brain regions for the juvenile GM1 patients. N/A are designations where the region was not calculated using the specified segmentation algorithm. The slope of the linear regression line is given in the top row, and the intercept is given in the bottom row for each cell. The 95% confidence interval for the slope and intercept shown in the brackets. For all linear regression modeling the automated volumetric calculations were along the y-axis, and the manual volumetric calculations were along the x-axis.

| Structure | Freesurfer | FSL | volBrain | SPM | SimNIBS (SPM+CAT) |
| --- | --- | --- | --- | --- | --- |
| **Whole Brain** | 0.85 [0.77, 0.95]<br>74.26 [-10.34, 158.9] | 0.82 [0.63, 1.02]<br>215.7 [20.80, 410.5] | 0.92 [0.86, 0.98]<br>92.83 [36.75, 148.9] | 1.06 [1.00, 1.12]<br>-109.9 [-170.1, -46.67] | 1.04 [0.92, 1.15]<br>-68.93 [-177.2, 39.38] |
| **Ventricles** | 0.97 [0.92, 1.02]<br>0.80 [-0.40, 2.00] | N/A | 0.79 [0.75, 0.83]<br>2.89 [0.88, 4.91] | N/A | 0.91 [0.87, 0.95]<br>0.22 [-2.24, 2.68] |
| **Cerebellum** | 1.00 [0.90, 1.11]<br>5.48 [-6.27, 17.24] | N/A | 1.04 [0.95, 1.12]<br>-2.67 [-12.42, 7.09] | N/A | 1.15 [0.86, 1.45]<br>3.55 [-29.67, 36.78] |
| **Thalamus** | 0.63 [0.39, 0.88]<br>6.53 [4.09, 8.98] | 0.71 [0.48, 0.93]<br>5.42 [3.30, 7.55] | 0.94 [0.79, 1.08]<br>1.74 [0.38, 3.11] | N/A | 0.76 [0.60, 0.91]<br>1.58 [0.09, 3.07] |
| **Caudate** | 0.83 [0.70, 0.96]<br>0.54 [-0.29, 1.37] | 0.59 [0.41, 0.77]<br>1.96 [0.86, 3.06] | 1.14 [1.02, 1.26]<br>-1.02 [-1.72, -0.33] | N/A | N/A |
| **Lentiform Nucleus** | 0.97 [0.69, 1.25]<br>0.39 [-2.28, 3.06] | 0.70 [0.44, 0.96]<br>2.94 [0.51, 5.36] | 0.70 [0.44, 0.96]<br>2.99 [0.51, 5.46] | N/A | N/A |
| **Corpus Callosum** | 0.23 [0.14, 0.32]<br>0.79 [0.04, 1.54] | N/A | N/A | N/A | 0.75 [0.40, 1.10]<br>11.96 [9.11, 14.81] |

**Table MIII.** Comparison of slopes and intercepts of the linear regression modeling between the 5 fully automated segmentation pipelines and the manual pipeline in the 7 different brain regions for the late-infantile GM1 patients. N/A are designations where the region was not calculated using the specified segmentation algorithm. The slope of the linear regression line is given in the top row, and the intercept is given in the bottom row for each cell. The 95% confidence interval for the slope and intercept shown in the brackets. For all linear regression modeling the automated volumetric calculations were along the y-axis, and the manual volumetric calculations were along the x-axis.

| Structure | Freesurfer | FSL | volBrain | SPM | SimNIBS<br>(SPM+CAT) |
| --- | --- | --- | --- | --- | --- |
| **Whole Brain** | 0.32 [-0.04, 00.69]<br>657.2 [361.0, 953.4] | 0.68 [-1.21, 2.57]<br>593.0 [-934.2, 2120] | 0.70 [0.53, 0.87]<br>349.7 [209.4, 490.0] | 0.68 [0.21, 1.15]<br>178.6 [-198.5, 555.8] | 0.68 [0.42, 0.93]<br>217.8 [10.00, 425.6] |
| **Ventricles** | 0.68 [0.35, 1.00]<br>7.70 [-13.27, 28.62] | N/A | 0.91 [0.87, 0.95]<br>0.22 [-2.24, 2.68] | N/A | 1.44 [1.36, 1.52]<br>-2.55 [-7.98, 2.87] |
| **Cerebellum** | 0.63 [0.27, 0.99]<br>38.87 [3.25, 74.49] | N/A | 0.98 [0.66, 1.31]<br>-8.23 [-23.71, 39.85] | N/A | 0.85 [0.61, 1.09]<br>25.10 [1.70, 48.49] |
| **Thalamus** | 0.97 [0.23, 1.71]<br>3.51 [-1.65, 8.66] | 1.38 [0.57, 2.19]<br>-1.85 [-7.50, 3.80] | 2.02 [1.41, 2.62]<br>-7.63 [-11.77, -3.50] | N/A | 0.92 [0.55, 1.28]<br>-0.23 [-2.73, 2.28] |
| **Caudate** | 0.70 [0.47, 0.94]<br>0.73 [-2.47, 2.27] | 0.32 [-0.12, 0.77]<br>1.97 [-0.92, 4.86] | 1.13 [0.62, 1.63]<br>-3.24 [-6.47, 0.00] | N/A | N/A |
| **Lentiform Nucleus** | 1.22 [0.51, 1.94]<br>-3.26 [-9.94, 3.43] | 1.32 [0.73, 1.90]<br>-5.59 [-10.95, -0.22] | 1.55 [0.70, 2.41]<br>-8.08 [-15.96, -0.20] | N/A | N/A |
| **Corpus Callosum** | 0.06 [-0.14, 0.26]<br>1.64 [0.60, 2.69] | N/A | N/A | N/A | -0.05 [-0.55, 0.45]<br>15.03 [12.56, 17.51] |

## References

1. O’Brien LM, Ziegler DA, Deutsch CK, Frazier JA, Herbert MR, Locascio JJ. Statistical adjustments for brain size in volumetric neuroimaging studies: some practical implications in methods. Psychiatry Res. 2011 Aug 30;193(2):113–22. doi: 10.1016/j.pscychresns.2011.01.007. PMID: 21684724; PMCID: PMC3510982.

2. Bottino, C. M., Castro, C. C., Gomes, R. L., Buchpiguel, C. A., Marchetti, R. L., & Neto, M. R. (2002). Volumetric MRI measurements can differentiate Alzheimer’s disease, mild cognitive impairment, and normal aging. International psychogeriatrics, 14(1), 59–72. 10.1017/s1041610202008281.

3. Pomponio, R., Erus, G., Habes, M., Doshi, J., Srinivasan, D., Mamourian, E., Bashyam, V., Nasrallah, I. M., Satterthwaite, T. D., Fan, Y., Launer, L. J., Masters, C. L., Maruff, P., Zhuo, C., Völzke, H., Johnson, S. C., Fripp, J., Koutsouleris, N., Wolf, D. H., Gur, R.,…Davatzikos, C. (2020). Harmonization of large MRI datasets for the analysis of brain imaging patterns throughout the lifespan. NeuroImage, 208, 116450. 10.1016/j.neuroimage.2019.116450.

4. Morey, R. A., Petty, C. M., Xu, Y., Hayes, J. P., Wagner, H. R., 2nd, Lewis, D. V., LaBar, K. S., Styner, M., & McCarthy, G. (2009). A comparison of automated segmentation and manual tracing for quantifying hippocampal and amygdala volumes. NeuroImage, 45(3), 855–866. 10.1016/j.neuroimage.2008.12.033.

5. Antonopoulos, G., More, S., Raimondo, F., Eickhoff, S. B., Hoffstaedter, F., & Patil, K. R. (2023). A systematic comparison of VBM pipelines and their application to age prediction. NeuroImage, 279, 120292. 10.1016/j.neuroimage.2023.120292.

6. Lenchik, L., Heacock, L., Weaver, A. A., Boutin, R. D., Cook, T. S., Itri, J., Filippi, C. G., Gullapalli, R. P., Lee, J., Zagurovskaya, M., Retson, T., Godwin, K., Nicholson, J., & Narayana, P. A. (2019). Automated Segmentation of Tissues Using CT and MRI: A Systematic Review. Academic radiology, 26(12), 1695–1706. 10.1016/j.acra.2019.07.006.

7. Giorgio, A. and De Stefano, N. (2013), Clinical use of brain volumetry. J. Magn. Reson. Imaging, 37: 1–14. 10.1002/jmri.23671.

8. Zoppo C, Kolstad J, Johnston J, D’Souza P, Kühn AL, Vardar Z, Peker A, Lindsay C, Rentiya ZS, King R, Gray-Edwards H, Vachha B, Acosta MT, Tifft CJ, Shazeeb MS. Quantitative reliability assessment of brain MRI volumetric measurements in type II GM1 gangliosidosis patients. Front Neuroimaging. 2024 Sep 13;3:1410848. doi: 10.3389/fnimg.2024.1410848. PMID: 39350771; PMCID: PMC11440193.

9. Singh, M. K., & Singh, K. K. (2021). A Review of Publicly Available Automatic Brain Segmentation Methodologies, Machine Learning Models, Recent Advancements, and Their Comparison. Annals of neurosciences, 28(1-2), 82–93. 10.1177/0972753121990175.

10. Sharma, N., & Aggarwal, L. M. (2010). Automated medical image segmentation techniques. Journal of medical physics, 35(1), 3–14. 10.4103/0971-6203.58777.

11. Bach Cuadra, M., Duay, V., Thiran, JP. (2015). Atlas-based Segmentation. In: Paragios, N., Duncan, J., Ayache, N. (eds) Handbook of Biomedical Imaging. Springer, Boston, MA. 10.1007/978-0-387-09749-7_12.

12. Pinto-Coelho L. (2023). How Artificial Intelligence Is Shaping Medical Imaging Technology: A Survey of Innovations and Applications. Bioengineering (Basel, Switzerland), 10(12), 1435. 10.3390/bioengineering10121435.

13. Wang, S., Pang, X., de Keyzer, F. et al. AI-based MRI auto-segmentation of brain tumor in rodents, a multicenter study. acta neuropathol commun 11, 11 (2023). 10.1186/s40478-023-01509-w.

14. Kuwabara, M., Ikawa, F., Nakazawa, S. et al. Artificial intelligence for volumetric measurement of cerebral white matter hyperintensities on thick-slice fluid-attenuated inversion recovery (FLAIR) magnetic resonance images from multiple centers. Sci Rep 14, 10104 (2024). 10.1038/s41598-024-60789-x.

15. Heinen, R., Steenwijk, M.D., Barkhof, F. et al. Performance of five automated white matter hyperintensity segmentation methods in a multicenter dataset. Sci Rep 9, 16742 (2019). 10.1038/s41598-019-52966-0.

16. Schell, M., Foltyn-Dumitru, M., Bendszus, M. et al. Automated hippocampal segmentation algorithms evaluated in stroke patients. Sci Rep 13, 11712 (2023). 10.1038/s41598-023-38833-z.

17. Opfer, R., Krüger, J., Spies, L., Ostwaldt, A. C., Kitzler, H. H., Schippling, S., & Buchert, R. (2023). Automatic segmentation of the thalamus using a massively trained 3D convolutional neural network: higher sensitivity for the detection of reduced thalamus volume by improved inter-scanner stability. European radiology, 33(3), 1852–1861. 10.1007/s00330-022-09170-y.

18. Yamanakkanavar, N., Choi, J. Y., & Lee, B. (2020). MRI Segmentation and Classification of Human Brain Using Deep Learning for Diagnosis of Alzheimer’s Disease: A Survey. Sensors (Basel, Switzerland), 20(11), 3243. 10.3390/s20113243.

19. Desikan, R. S., Ségonne, F., Fischl, B., Quinn, B. T., Dickerson, B. C., Blacker, D., Buckner, R. L., Dale, A. M., Maguire, R. P., Hyman, B. T., Albert, M. S., & Killiany, R. J. (2006). An automated labeling system for subdividing the human cerebral cortex on MRI scans into gyral based regions of interest. NeuroImage, 31(3), 968–980. 10.1016/j.neuroimage.2006.01.021.

20. Kim, MJ., Hong, E., Yum, MS. et al. Deep learning-based, fully automated, pediatric brain segmentation. Sci Rep 14, 4344 (2024). 10.1038/s41598-024-54663-z.

21. Billot, B., Bocchetta, M., Todd, E., Dalca, A. V., Rohrer, J. D., & Iglesias, J. E. (2020). Automated segmentation of the hypothalamus and associated subunits in brain MRI. NeuroImage, 223, 117287. 10.1016/j.neuroimage.2020.117287.

22. Nigro, S., Cerasa, A., Zito, G., Perrotta, P., Chiaravalloti, F., Donzuso, G., Fera, F., Bilotta, E., Pantano, P., Quattrone, A., & Alzheimer’s Disease Neuroimaging Initiative (2014). Fully automated segmentation of the pons and midbrain using human T1 MR brain images. PloS one, 9(1), e85618. 10.1371/journal.pone.0085618.

23. Choi, U. S., Sung, Y. W., & Ogawa, S. (2024). deepPGSegNet: MRI-based pituitary gland segmentation using deep learning. Frontiers in endocrinology, 15, 1338743. 10.3389/fendo.2024.1338743.

24. van Elst, S., de Bloeme, C. M., Noteboom, S., de Jong, M. C., Moll, A. C., Göricke, S., de Graaf, P., & Caan, M. W. A. (2023). Automatic segmentation and quantification of the optic nerve on MRI using a 3D U-Net. Journal of medical imaging (Bellingham, Wash.), 10(3), 034501. 10.1117/1.JMI.10.3.034501.

25. D’Souza, P., Farmer, C., Johnston, J. M., Han, S. T., Adams, D., Hartman, A. L., Zein, W., Huryn, L. A., Solomon, B., King, K., Jordan, C. P., Myles, J., Nicoli, E. R., Rothermel, C. E., Mojica Algarin, Y., Huang, R., Quimby, R., Zainab, M., Bowden, S., Crowell, A.,…Tifft, C. J. (2024). GM1 gangliosidosis type II: Results of a 10-year prospective study. Genetics in medicine: official journal of the American College of Medical Genetics, 26(7), 101144. 10.1016/j.gim.2024.101144.

26. Laur D, Pichard S, Bekri S, et al. Natural history of GM1 gangliosidosis—Retrospective cohort study of 61 French patients from 1998 to 2019. J Inherit Metab Dis. 2023; 46(5): 972–981. doi:10.1002/jimd.12646.

27. Lang FM, Korner P, Harnett M, Karunakara A, Tifft CJ. The natural history of Type 1 infantile GM1 gangliosidosis: A literature-based meta-analysis. Mol Genet Metab. 2020 Mar;129(3):228–235. doi: 10.1016/j.ymgme.2019.12.012. Epub 2019 Dec 30. PMID: 31937438; PMCID: PMC7093236.

28. Nicoli E-R, Annunziata I, d’Azzo A, Platt FM, Tifft CJ and Stepien KM (2021) GM1 Gangliosidosis—A Mini-Review. Front. Genet. 12:734878. doi: 10.3389/fgene.2021.734878.

29. Liu, S., Ma, W., Feng, Y., Zhang, Y., Jia, X., Tang, C., Tang, F., Wu, X., & Huang, Y. (2022). AAV9-coGLB1 Improves Lysosomal Storage and Rescues Central Nervous System Inflammation in a Mutant Mouse Model of GM1 Gangliosidosis. Current gene therapy, 22(4), 352–365.

30. Gray-Edwards HL, Maguire AS, Salibi N, Ellis LE, Voss TL, Diffie EB, Koehler J, Randle AN, Taylor AR, Brunson BL, Denney TS, Beyers RJ, Gentry AS, Gross AL, Batista AR, Sena-Esteves M, Martin DR. 7T MRI Predicts Amelioration of Neurodegeneration in the Brain after AAV Gene Therapy. Mol Ther Methods Clin Dev. 2019 Dec 24;17:258–270. doi: 10.1016/j.omtm.2019.11.023. PMID: 31970203; PMCID: PMC6962699.

31. Regier, D. S., Kwon, H. J., Johnston, J., Golas, G., Yang, S., Wiggs, E., Latour, Y., Thomas, S., Portner, C., Adams, D., Vezina, G., Baker, E. H., & Tifft, C. J. (2016). MRI/MRS as a surrogate marker for clinical progression in GM1 gangliosidosis. American journal of medical genetics. Part A, 170(3), 634–644. 10.1002/ajmg.a.37468.

32. Nestrasil I, Ahmed A, Utz JM, Rudser K, Whitley CB, Jarnes-Utz JR. Distinct progression patterns of brain disease in infantile and juvenile gangliosidoses: Volumetric quantitative MRI study. Mol Genet Metab. 2018 Feb;123(2):97–104. doi: 10.1016/j.ymgme.2017.12.432. Epub 2017 Dec 20. PMID: 29352662; PMCID: PMC5832355.

33. National Human Genome Research Institute. Natural History of Glycosphingolipid Storage Disorders and Glycoprotein Disorders ClinicalTrials.gov identifier: NCT00029965. Updated August 7, 2024. Accessed August 12, 2024. https://clinicaltrials.gov/study/NCT00029965.

34. Reynolds JE, Long X, Paniukov D, Bagshawe M, Lebel C. Calgary Preschool magnetic resonance imaging (MRI) dataset. Data Brief. 2020 Jan 31;29:105224. doi: 10.1016/j.dib.2020.105224. PMID: 32071993; PMCID: PMC7016255.

35. Mah A, Geeraert B, Lebel C (2017) Detailing neuroanatomical development in late childhood and early adolescence using NODDI. PLoS ONE 12(8): e0182340. 10.1371/journal.pone.0182340.

36. Li X, Morgan PS, Ashburner J, Smith J, Rorden C. The first step for neuroimaging data analysis: DICOM to NIfTI conversion. J Neurosci Methods. 2016 May 1;264:47–56. doi: 10.1016/j.neumeth.2016.03.001. Epub 2016 Mar 2. PMID: 26945974.

37. D. L. Collins, P. Neelin, T. M. Peters, and A. C. Evans, “Automatic 3d intersubject registration of mr volumetric data in standardized talairach space.,” Journal of computer assisted tomography, vol. 18, no. 2, pp. 192–205, 1994.

38. A. M. Dale, B. Fischl, and M. I. Sereno, “Cortical surface-based analysis: I. segmentation and surface reconstruction,” Neuroimage, vol. 9, no. 2, pp. 179–194, 1999.

39. B. Fischl, M. I. Sereno, and A. M. Dale, “Cortical surface-based analysis: Ii: inflation, flattening, and a surface-based coordinate system,” Neuroimage, vol. 9, no. 2, pp. 195– 207, 1999. 3

40. B. Fischl, M. I. Sereno, R. B. Tootell, and A. M. Dale, “High-resolution intersubject averaging and a coordinate system for the cortical surface,” Human brain mapping, vol. 8, no. 4, pp. 272–284, 1999.

41. B. Fischl and A. M. Dale, “Measuring the thickness of the human cerebral cortex from magnetic resonance images,” Proceedings of the National Academy of Sciences, vol. 97, no. 20, pp. 11050–11055, 2000.

42. B. Fischl, A. Liu, and A. M. Dale, “Automated manifold surgery: constructing geometrically accurate and topologically correct models of the human cerebral cortex,” IEEE transactions on medical imaging, vol. 20, no. 1, pp. 70–80, 2001.

43. “Non-uniform intensity correction,” http://www.bic.mni.mcgill.ca/software/N3/node6.html.

44. B. Fischl, D. H. Salat, E. Busa, M. Albert, M. Dieterich, C. Haselgrove, A. Van Der Kouwe, R. Killiany, D. Kennedy, S. Klaveness, et al., “Whole brain segmentation: automated labeling of neuroanatomical structures in the human brain,” Neuron, vol. 33, no. 3, pp. 341–355, 2002.

45. B. Fischl, A. Van Der Kouwe, C. Destrieux, E. Halgren, F. Śegonne, D. H. Salat, E. Busa, L. J. Seidman, J. Goldstein, D. Kennedy, et al., “Automatically parcellating the human cerebral cortex,” Cerebral cortex, vol. 14, no. 1, pp. 11–22, 2004.

46. Woolrich, M. W., Jbabdi, S., Patenaude, B., Chappell, M., Makni, S., Behrens, T., Beckmann, C., Jenkinson, M., & Smith, S. M. (2009). Bayesian analysis of neuroimaging data in FSL. NeuroImage, 45(1 Suppl), S173–S186. 10.1016/j.neuroimage.2008.10.055.

47. Smith, S. M., Jenkinson, M., Woolrich, M. W., Beckmann, C. F., Behrens, T. E., Johansen-Berg, H., Bannister, P. R., De Luca, M., Drobnjak, I., Flitney, D. E., Niazy, R. K., Saunders, J., Vickers, J., Zhang, Y., De Stefano, N., Brady, J. M., & Matthews, P. M. (2004). Advances in functional and structural MR image analysis and implementation as FSL. NeuroImage, 23 Suppl 1, S208–S219. 10.1016/j.neuroimage.2004.07.051.

48. Jenkinson, M., Beckmann, C. F., Behrens, T. E., Woolrich, M. W., & Smith, S. M. (2012). FSL. NeuroImage, 62(2), 782–790. 10.1016/j.neuroimage.2011.09.015.

49. Manjón, J. V., & Coupé, P. (2016). volBrain: An Online MRI Brain Volumetry System. Frontiers in neuroinformatics, 10, 30. 10.3389/fninf.2016.00030.

50. Manjón, J. V., Romero, J. E., Vivo-Hernando, R., Rubio, G., Aparici, F., de la Iglesia-Vaya, M., & Coupé, P. (2022). vol2Brain: A New Online Pipeline for Whole Brain MRI Analysis. Frontiers in neuroinformatics, 16, 862805. 10.3389/fninf.2022.862805.

51. Penny, W. D., Friston, K. J., Ashburner, J. T., Kiebel, S. J., & Nichols, T. E. (Eds.). (2011). Statistical parametric mapping: the analysis of functional brain images. Elsevier.

52. Nielsen, J. D., Madsen, K. H., Puonti, O., Siebner, H. R., Bauer, C., Madsen, C. G., Saturnino, G. B., & Thielscher, A. (2018). Automatic skull segmentation from MR images for realistic volume conductor models of the head: Assessment of the state-of-the-art. NeuroImage, 174, 587–598. 10.1016/j.neuroimage.2018.03.001.

53. Dahnke, R., Yotter, R. A., & Gaser, C. (2013). Cortical thickness and central surface estimation. NeuroImage, 65, 336–348. 10.1016/j.neuroimage.2012.09.050.

54. Stolte SE, Indahlastari A, Chen J, Albizu A, Dunn A, Pedersen S, See KB, Woods AJ, Fang R. Precise and Rapid Whole-Head Segmentation from Magnetic Resonance Images of Older Adults using Deep Learning. Imaging Neurosci (Camb). 2024 Mar;2:10.1162/imag_a_00090. doi: 10.1162/imag_a_00090. Epub 2024 Feb 13. PMID: 38465203; PMCID: PMC10922731.

55. Tavares, V., Prata, D., & Ferreira, H. A. (2019). Comparing SPM12 and CAT12 segmentation pipelines: a brain tissue volume-based age and Alzheimer’s disease study. Journal of neuroscience methods, 334, 108565. Advance online publication. 10.1016/j.jneumeth.2019.108565.

56. Hammers, A., Allom, R., Koepp, M. J., Free, S. L., Myers, R., Lemieux, L., Mitchell, T. N., Brooks, D. J., & Duncan, J. S. (2003). Three-dimensional maximum probability atlas of the human brain, with particular reference to the temporal lobe. Human brain mapping, 19(4), 224–247. 10.1002/hbm.10123.

57. Platten, M., Brusini, I., Andersson, O., Ouellette, R., Piehl, F., Wang, C., & Granberg, T. (2021). Deep Learning Corpus Callosum Segmentation as a Neurodegenerative Marker in Multiple Sclerosis. Journal of neuroimaging: official journal of the American Society of Neuroimaging, 31(3), 493–500. 10.1111/jon.12838.

58. Brusini, I., Platten, M., Ouellette, R., Piehl, F., Wang, C., & Granberg, T. (2022). Automatic deep learning multicontrast corpus callosum segmentation in multiple sclerosis. Journal of neuroimaging: official journal of the American Society of Neuroimaging, 32(3), 459–470. 10.1111/jon.12972.

59. Vachet, C., Yvernault, B., Bhatt, K., Smith, R. G., Gerig, G., Hazlett, H. C., & Styner, M. (2012). Automatic corpus callosum segmentation using a deformable active Fourier contour model. Proceedings of SPIE--the International Society for Optical Engineering, 8317, 831707–. 10.1117/12.911504.

60. Kolstad J, Zoppo C, Johnston JM, D’Souza P, Kühn AL, Vardar Z, Peker A, Hader A, Celik H, Lewis CJ, Lindsay C, Rentiya ZS, Lebel C, Vedantham S, Vachha B, Gray-Edwards HL, Acosta MT, Tifft CJ, Shazeeb MS. Natural history progression of MRI brain volumetrics in type II late-infantile and juvenile GM1 gangliosidosis patients. Mol Genet Metab. 2025 Jan 19;144(3):109025. doi: 10.1016/j.ymgme.2025.109025. Epub ahead of print. PMID: 39874851.

61. Zou, K. H., Warfield, S. K., Bharatha, A., Tempany, C. M., Kaus, M. R., Haker, S. J., Wells, W. M., 3rd, Jolesz, F. A., & Kikinis, R. (2004). Statistical validation of image segmentation quality based on a spatial overlap index. Academic radiology, 11(2), 178–189. 10.1016/s1076-6332(03)00671-8.

62. Deeley, M. A., Chen, A., Datteri, R., Noble, J. H., Cmelak, A. J., Donnelly, E. F., Malcolm, A. W., Moretti, L., Jaboin, J., Niermann, K., Yang, E. S., Yu, D. S., Yei, F., Koyama, T., Ding, G. X., & Dawant, B. M. (2011). Comparison of manual and automatic segmentation methods for brain structures in the presence of space-occupying lesions: a multi-expert study. Physics in medicine and biology, 56(14), 4557–4577. 10.1088/0031-9155/56/14/021.

63. Aydin, O. U., Taha, A. A., Hilbert, A., Khalil, A. A., Galinovic, I., Fiebach, J. B., Frey, D., & Madai, V. I. (2021). On the usage of average Hausdorff distance for segmentation performance assessment: hidden error when used for ranking. European radiology experimental, 5(1), 4. 10.1186/s41747-020-00200-2.

## Supplementary References

64. National Human Genome Research Institute. Natural History of Glycosphingolipid Storage Disorders and Glycoprotein Disorders ClinicalTrials.gov identifier: NCT00029965. Updated August 7, 2024. Accessed August 12, 2024. https://clinicaltrials.gov/study/NCT00029965.

65. Reynolds JE, Long X, Paniukov D, Bagshawe M, Lebel C. Calgary Preschool magnetic resonance imaging (MRI) dataset. Data Brief. 2020 Jan 31;29:105224. doi: 10.1016/j.dib.2020.105224. PMID: 32071993; PMCID: PMC7016255.

66. Mah A, Geeraert B, Lebel C (2017) Detailing neuroanatomical development in late childhood and early adolescence using NODDI. PLoS ONE 12(8): e0182340. 10.1371/journal.pone.0182340.

